# Principled distillation of multidimensional UK Biobank data reveals insights into the correlated human phenome

**DOI:** 10.1101/2022.09.02.22279546

**Authors:** Caitlin E. Carey, Rebecca Shafee, Amanda Elliott, Duncan S. Palmer, John Compitello, Masahiro Kanai, Liam Abbott, Patrick Schultz, Konrad J. Karczewski, Samuel C. Bryant, Caroline M. Cusick, Claire Churchhouse, Daniel P. Howrigan, Daniel King, George Davey Smith, Robbee Wedow, Benjamin M. Neale, Raymond K. Walters, Elise B. Robinson

**Author notes:** These authors contributed equally (Benjamin M. Neale, Raymond K. Walters, Elise B. Robinson). Correspondence to Caitlin E. Carey, and Raymond K. Walters.

## Abstract

Broad yet detailed data collected in biobanks captures variation reflective of human health and behavior, but insights are hard to extract given their complexity and scale. In the largest factor analysis to date, we distill hundreds of medical record codes, physical assays, and survey items from UK Biobank into 35 understandable latent constructs. The identified factors recapitulate known disease classifications, highlight the relevance of psychiatric constructs, improve measurement of health-related behavior, and disentangle elements of socioeconomic status. We demonstrate the power of this principled data reduction approach to clarify genetic signal, enhance discovery, and identify associations between underlying phenotypic structure and health outcomes such as mortality. We emphasize the importance of considering the interwoven nature of the human phenome when evaluating large-scale patterns relevant to public health.

## Introduction

The modern era is one of vast data generation, affording unprecedented opportunities to characterize the world around us. Decades of public and private investment in large-scale data collection and aggregation has in recent years yielded data repositories called biobanks linking health outcomes to biological samples for hundreds of thousands of individuals (e.g., (1–4)). These massive resources now power discovery in human health and disease (5–7).

Biobank-scale data are multidimensional and include diverse data types, often including thousands of variables drawn from electronic health records (EHR), self-report survey measures, laboratory assays, and physical and cognitive assessments (4). The tremendous breadth and depth of data can obfuscate larger patterns present across a biobank. For example, indicators of a particular health construct might be reflected across a range of variables nested within different data modalities, in ways that are both anticipated and unexpected.

To more completely consider the correlated human health landscape, approaches are needed that can reduce thousands of variables into a smaller number of constructs. In refining data in a way that is digestible and scalable for human interpretation, such approaches should ideally move beyond a “black box” and allow us to look “under the hood” at relationships between variables. Though a host of data reduction methods have been well established and used at scale(8–10), factor analysis (FA; (11–14)), a model-based statistical technique developed in the social sciences, is optimized for these purposes.

Here, we perform the largest factor analysis to date to explore the cross-sectional phenome of the UK Biobank (UKB) (4). We integrate linked phenotypic and genetic data to demonstrate that this principled distillation can power new insights into human health. This approach takes us closer to the next frontier of biobank-driven discovery, which will be to better characterize the complex and multifaceted human phenome.

### Distilling the phenotypic landscape

We use a multi-stage factor analytic approach to distill UKB’s phenotypic landscape, first considering 2772 phenotypes and all unrelated individuals with predominantly estimated European genetic ancestry (N=361,144; **Fig S1**; **Supplementary Text**). FA treats observed variables (items) as measures of a small number of unobserved (latent) factors *F*, with corresponding effect sizes, or “loadings,” *⩘*, and item-specific residuals *ϵ*, allowing the observed covariance between items *Σ* to be fit as *Σ* = *⩘Cov*(*F*)*⩘*′ + *Cov*(*ϵ*) (13). We deconvolve the latent factors and loadings by prioritizing sparsity criteria for the loadings while requiring the factors to be statistically independent (orthogonal). The content of each fitted factor can therefore be interpreted as reflecting the correlation between sets of items conditional on the relationships captured by other factors (**Supplementary Text**). To reduce the impact of differential availability of certain assessments across participants, we begin by fitting a factor model in a core subsample of 42,325 individuals with high assessment-level completion and 898 phenotypes with low (i.e., mean=9.1%, s.d.=10.7%) missingness and prevalence above 1% (**Supplementary Text**).

The final factor model consists of 35 orthogonal latent factors drawn from 505 observed items in UKB (**Fig 1**; **Fig S2**). An initial exploratory model explains 18.5% of overall variance across input phenotypes in a modeling sample (N=33,860). While this is low for a traditional factor analysis with a much smaller set of individual items taken from a single survey, the aim was to extract major axes of variation across a biobank with a broad and diverse set of items. A reduced and refined confirmatory FA demonstrates acceptable fit in a holdout sample of 8465 UKB participants (RMSEA=0.028; see **Supplementary Text**). On average, each factor influences 32.49 items (s.d.=20.48; range 3-84; **Table S1**), with items known to be correlates of numerous health outcomes, such as “Health satisfaction” and “Overall health rating” (15–19), loading on as many as 10 factors.

**Fig. 1.**
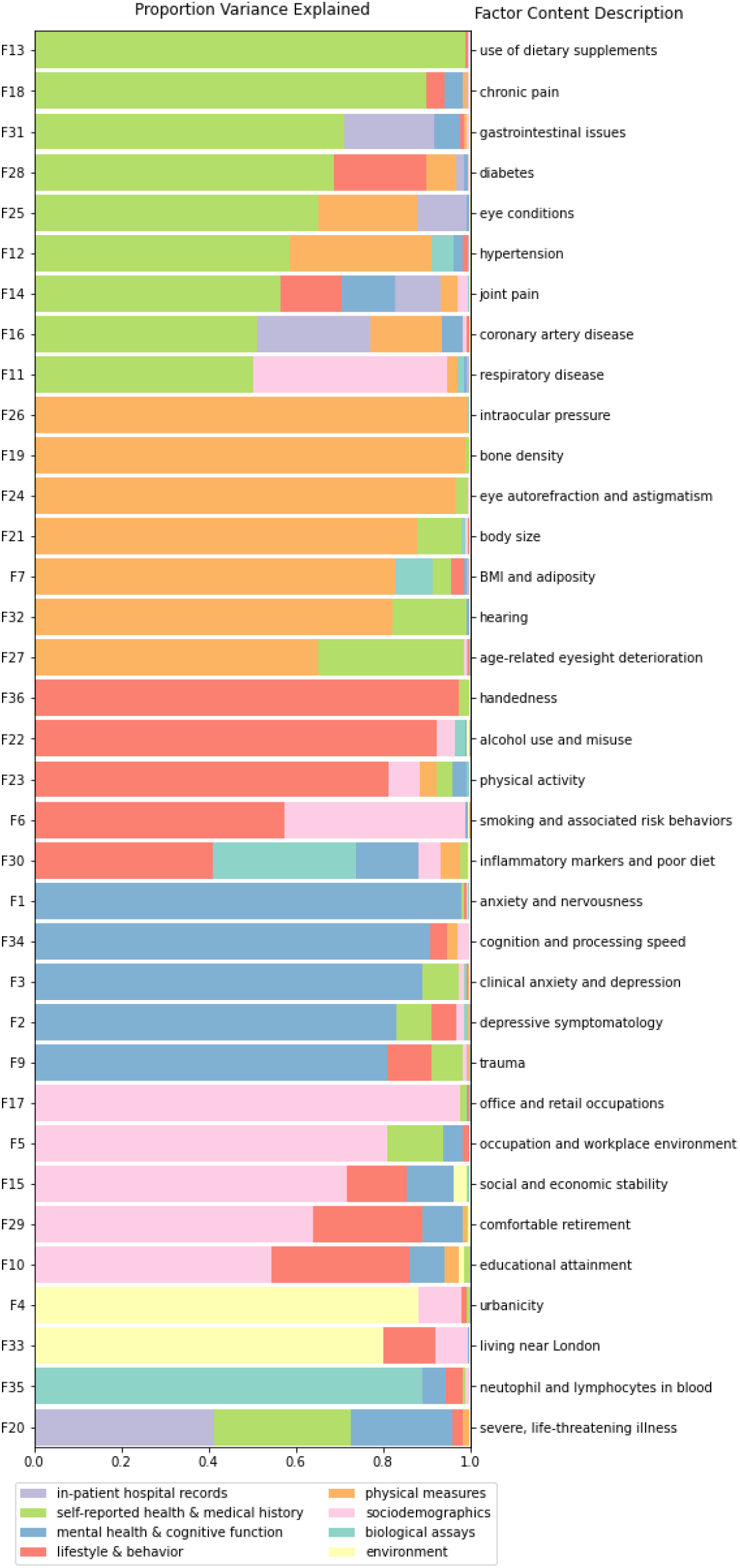
Makeup of factors in the final model. Horizontal bars represent proportion variance explained in a given factor score by each of 8 major categories of assessment in UKB, estimated using hierarchical partitioning. To the left, factors are numbered in order of variance extraction in the exploratory factor analysis. To the right are listed brief descriptions of the items contained within a factor.

Because individual assessments in UKB were designed to capture specific phenotypic domains, many factors draw heavily from individual categories within the 72 UKB questionnaires and assessments included in the factor model (**Table S2**), with nine deriving most of their items from a single source. Most factors, however, draw items from multiple sources (mean=11.11; s.d.=6.26; range 2-26), highlighting the utility of a factor analytic approach in identifying phenotypic structure *across* questionnaires. Factor 12, for example, captures correlates of hypertension (HTN) across the phenome including diagnostic items (e.g., self-reported HTN and measured blood pressure), risk factors (e.g., family history, BMI, and waist circumference; (20)), comorbidity (e.g., self-reported high cholesterol and diabetes; (21,22)), and relevant medications (e.g., diuretics and calcium channel blockers). Beyond these more expected associations, factors capture less established links between variables such as BMI and frequency of computer gaming (i.e., Factor 7), diet and education (i.e., Factor 10), and IT occupations and performance on computerized cognitive tests (i.e., Factor 34).

The loadings of a factor’s component items reflect their relative importance within the construct captured (11–13). Using these loadings, we categorize the factors into five broad domains: 1) medical disease (e.g., respiratory disease [F11], hypertension [F12], and coronary artery disease [F16]); 2) physical characteristics (e.g., BMI and adiposity [F7], bone mineral density [F19], and body size [F21]), 3) health behaviors (e.g., smoking [F6], alcohol intake [F22], and physical activity [F23]); 4) psychiatric and cognitive outcomes (e.g., clinical anxiety and depression [F3], trauma [F9], and processing speed [F34]); and 5) sociodemographics (e.g., occupation [F5], educational attainment [F10], and social and economic stability [F15]).

### Characterizing factors with linked biobank data

To further characterize the factors and their relation to public health outcomes, we generate weighted sum scores indexing individuals’ values for each factor in the full sample of individuals with predominately European estimated genetic ancestry (N=361,099, due to subsequent participant withdrawals from UKB). Scores are computed based on the fitted factor model using weighted linear combinations of observed items while accounting for missingness (**Supplementary Text**). Using these scores, we conduct a series of follow-up analyses including: correlating factors with 403 top-level medical diagnostic codes (or “phecodes”; **Table S3**; **Fig S3**) and 28 biomarkers (**Fig S4**); predicting all-cause mortality from survey completion to the last date at which participant death records are available (**Fig 2a**); and performing a series of genetic analyses including GWAS of the factors (**Table S4**), heritability estimation and enrichment (**Fig 2b**; **Fig S5**), and genetic correlations (**Fig S6**).

**Fig. 2.**
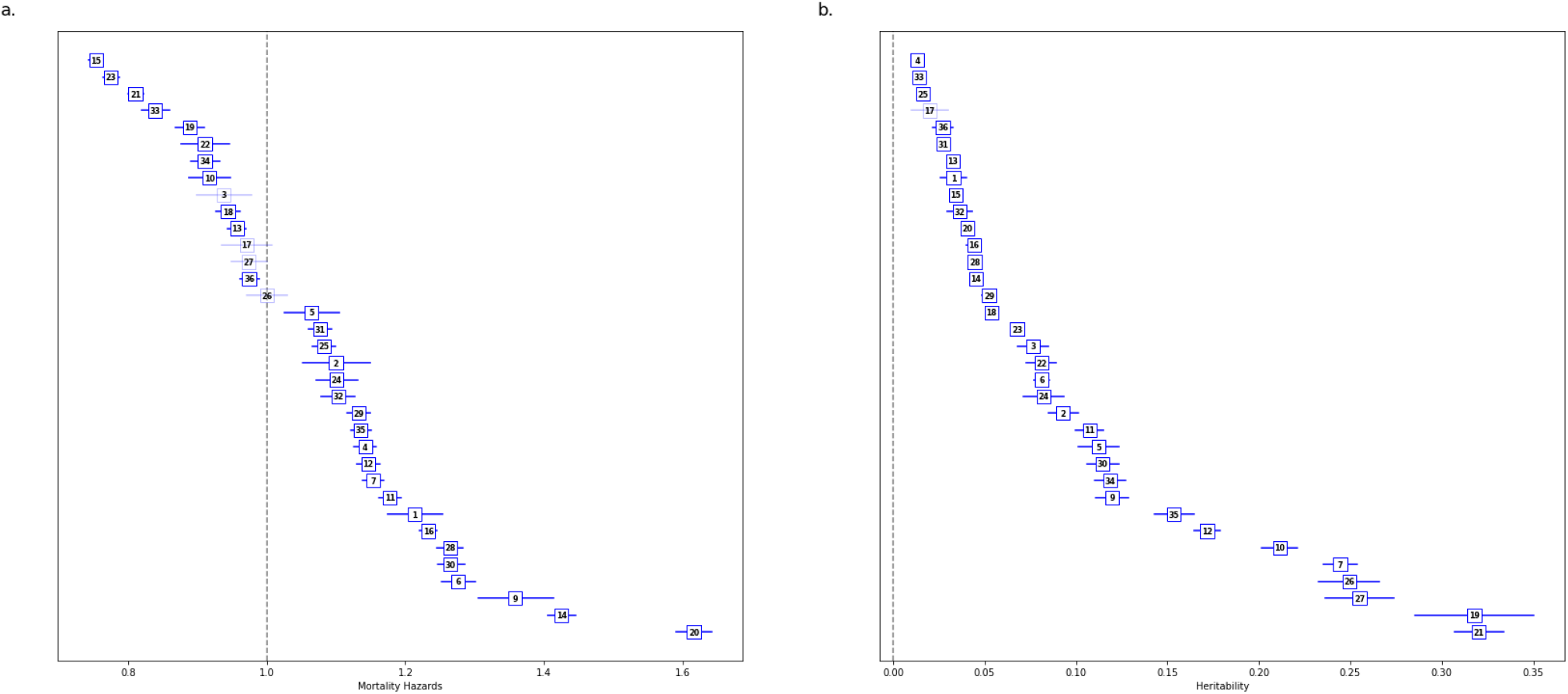
Prospective mortality hazard ratios and heritability estimates across all 35 factors. (**A**) Mortality hazards per factor. Factors are ordered from most protective to most predictive of mortality from time of last survey completion to the last date at which death records were available for analysis. (**B**) Heritability estimates for each factor. Factors are ordered by heritability point estimates. Error bars in Panel **A** reflect 95% Cis, while error bars in Panel **B** reflect standard errors. For both panels, darker blue boxes remain significant after adjustment for multiple comparisons.

All but three factors are associated with prospective mortality after correction for multiple testing (i.e., p<0.0014 for 35 factors), reflecting the significance of these axes of variation across individuals (**Fig 2a**). Factor 20, which includes items like number of surgeries, cancer diagnosis, and “diagnosis with a life-threatening illness,” has the highest mortality prediction across all 35 factors (Hazard Ratio [HR]=1.62[1.59-1.64]). Other factors among the strongest predictors of mortality capture constructs such as joint pain and disability (F14), trauma (F9), social and economic stability (F15), and physical activity (F23). These results provide a benchmark for assessing which constructs are most central to health outcomes and highlight the utility of using factor scores prospectively as additional longitudinal data is incorporated into the biobank.

All but one factor are significantly heritable after multiple testing correction, though SNP heritability estimates vary widely based on factor content (mean factor h_g_^2^=0.10[0.09]; **Fig 2b**). Factors that capture measured physical characteristics have higher heritabilities on average (mean=0.22, s.d.=0.10), with Factor 21, which captures body size, having the highest overall SNP heritability (h_g_^2^=0.32[0.01]), and Factor 33, which captures geographic and cultural indicators of living near London, having the lowest point estimate (h_g_^2^=0.01[0.003]). Heritability enrichment analyses reveal associations consistent with known underlying biology (**Fig S5**). For example, Factor 21 is enriched for regions of the genome associated with musculoskeletal and connective cell types (p=3.75e-07). CNS tissues are the most widely enriched cell-type group across factors (i.e., significant at bonf. p<0.05 in 8 factors).

Factor analysis can also aid in genetic discovery by combining shared information across items, which decreases measurement error of the underlying construct. Indeed, the observed SNP heritability of the 35 factors is on the whole higher than for the 505 component items (mean item h_g_^2^=0.05[0.07]; 2-sample t-test p=0.002; **Fig 3a**). Within factors, 20 of 35 have higher SNP heritability point estimates than all 5 of their top-loading items (e.g., **Fig 3b**), with the largest gains observed for factors containing mostly dichotomous or ordinal self-report items; these items are likely to have higher measurement error than empirically measured continuous items (Supplementary Note of (23); **Fig S7**). Across all 35 factors this increase in power from modelling covariance across items yields 548 loci, of 2329 total, that are not genome-wide significant in GWAS of their top 5 component items (**Fig 3c**). This capture of shared information across component items is further reflected in factor GWAS identifying nearly all (i.e., 91.5%) loci significant in at least 3 of 5 top-loading items, but much fewer (i.e., 20.7%) of loci significant in only 1 of the top items, which are thus likely to be item-specific (**Fig 3c**; e.g., **Fig 3d**). In other words, loci that are common to multiple component items are more likely to capture shared covariance across these items and thus be picked up in the factor GWAS.

**Fig. 3.**
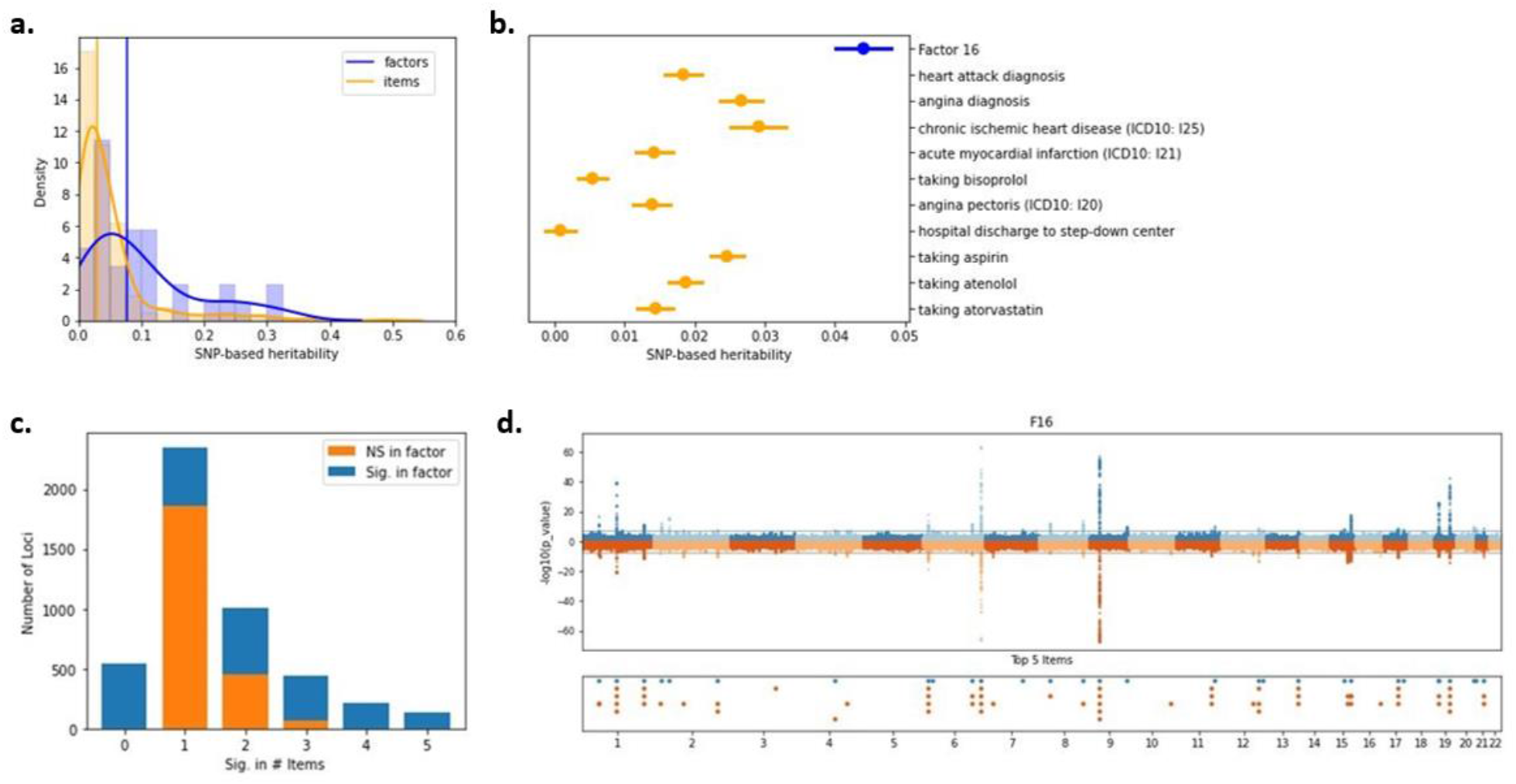
Genetic properties of factors vs. items. (**A**) Distributions of SNP-based heritability point estimates for items and factors, with density curves overlaid. Vertical lines represent the median point estimate for each category. (**B**) SNP heritability point estimates, with standard error bars shown, for an example factor, Factor 16, and its top 10 component items by loading. (**C**) Number of GWAS significant loci (p<5×10^−8^) across all 35 factors and their top 5 component items by loading. Loci shown in orange are significant only in GWAS of one or more top items. Loci shown in blue are significant in GWAS of the factors. For example, there are 2350 loci that are significant in GWAS of only one of a factor’s top 5 items (second bar in the graph). Of these loci, 486 are also significant in GWAS of the corresponding factor (shown in blue). (**D**) Comparison of loci identified in Factor 16 (top of Miami plot) versus its top 5 items by loading (bottom of Miami plot). Below the Miami plot are all loci across the factor (in blue) and top items (in orange), demonstrating the patterns presented in (C) at the single-factor level.

Combining factor definitions with individual-level data and linked biobank phenotypes allows us to further characterize the factors and discover new associations relevant for health. Beyond the general trends described above, in the next sections we highlight key findings in vignettes of individual factors from the medical, psychiatric/cognitive, behavioral, and sociodemographic domains.

### Recapitulating known disease nosology and biology

Factors in the medical domain, for which nosology is well-defined, recapitulate prior clinical, epidemiological, and biological knowledge, without any expert or manual curation. Factor 11, for example, captures items related to respiratory disease, including self-report diagnoses (e.g., asthma, hayfever, and COPD); medications (e.g., bronchodilators and antihistamines); symptoms (e.g., wheezing, cough, and sputum production); and laboratory findings (e.g., eosinophil counts and forced expiratory volume [FEV1]; **Table S1**). Scores on the factor are associated with severe disease such as respiratory failure (OR=1.53[1.48-1.58]) and pulmonary fibrosis (OR=1.55[1.48-1.64]) in inpatient hospital records (**Table S3**), as well as with increased mortality risk (HR: 1.18[1.16-1.19]; **Fig 2a**). It is also associated with serum inflammatory biomarker C-reactive protein levels (β=0.082[0.002], z=39.041; **Fig S4**), and its heritability is enriched in regions of the genome associated with blood and immune cell types (p=1.61×10^−6^; **Fig S5**), highlighting a known key role for the immune system in the pathogenesis of chronic respiratory disease. Genetic correlation with a prior GWAS of asthma is 0.89(0.01; (24)).

One of the clearest alignments between the factor analysis and a known disease is observed in Factor 16, which captures commonly used diagnostic indicators of coronary artery disease. Items in the factor include self-report and hospital inpatient diagnoses (e.g., chronic ischemic heart disease and myocardial infarction; (25)); symptoms (e.g., angina, pain in the chest or throat, and shortness of breath; (26)); and medications (e.g., aspirin, beta-blockers, and statins; (27); **Table S1**). Factor 16 has a strong genetic correlation with a prior CAD GWAS (r_g_=0.87[0.02]; (28)), and GWAS of Factor 16 captures known lipid biology, with many of the 33 significant loci mapped to core lipid metabolism genes such as *LPA, LPL, LDLR, SORT1, APOE*, and *PCSK9* (**Fig 3d**). These genes have been implicated in the development of CAD and cardiometabolic disease more broadly (28–30). These results suggest that Factor 16 provides a strong data-driven approximation of CAD, consistent with previous efforts to define CAD status based on medical records either algorithmically or with clinical curation (28,31), but with a possible twist towards other correlates of cardiovascular disease, and in the absence of angiographic measures.

When studying individual diseases there is of course no substitute for rigorously obtained clinical assessments, repeated over time, though factors return strong approximations. While factors should not be interpreted as diagnostic tools, the results show that factor analysis at biobank scale pushes beyond individual health measures to paint a broader picture of the medical phenome, more explicitly modeling the correlated nature of comorbidities often unmeasured, yet captured implicitly, in case-control designs.

### Discovering pervasive associations between trauma and public health outcomes

Unlike somatic diseases with well characterized, directly measurable criteria, psychiatric constructs are not biologically defined, and diagnostic boundaries are thus imprecise.

Experiences of trauma, for example, represent one of the most critical but understudied public health concerns globally, with significant associations to downstream chronic health problems and mortality (32–35). Factor 9 places trauma within the larger behavioral and medical landscape without the need to decide which trauma-related measures to include or how to combine and weight them. The factor’s component items include exposure to traumas like feeling hated as a child or being physically abused by a family member, as well as related mental health outcomes like PTSD, major depressive disorder, mania, psychosis, addiction, and self-harm (**Table S1**).

Of the 35 factors, Factor 9 has one of the highest prospective all-cause mortality hazards (HR=1.36[1.31-1.41]; **Fig 2a**), reinforcing the critical public health importance of the construct. Its associations with psychiatric and medical diagnoses are uniquely broad (**Fig 4**), with strong phenotypic correlations observed across all diagnostic categories, including common circulatory (e.g., hypertension, OR=1.29[1.27-1.32]), digestive (e.g., acid reflux, OR=1.34[1.30-1.38]), respiratory (e.g., asthma, OR=1.39[1.36-1.43]), and endocrine (e.g., type 2 diabetes OR=1.62[1.57-1.68]) outcomes (**Table S3**). The biomarker most associated with this factor is C-reactive protein (β=0.11[0.004], z=31.47), a blood-based indicator of inflammation and inflammatory disorders, providing further evidence for its relevance as a clinical flag for suites of trauma-related exposures and outcomes (36–41).

**Fig. 4.**
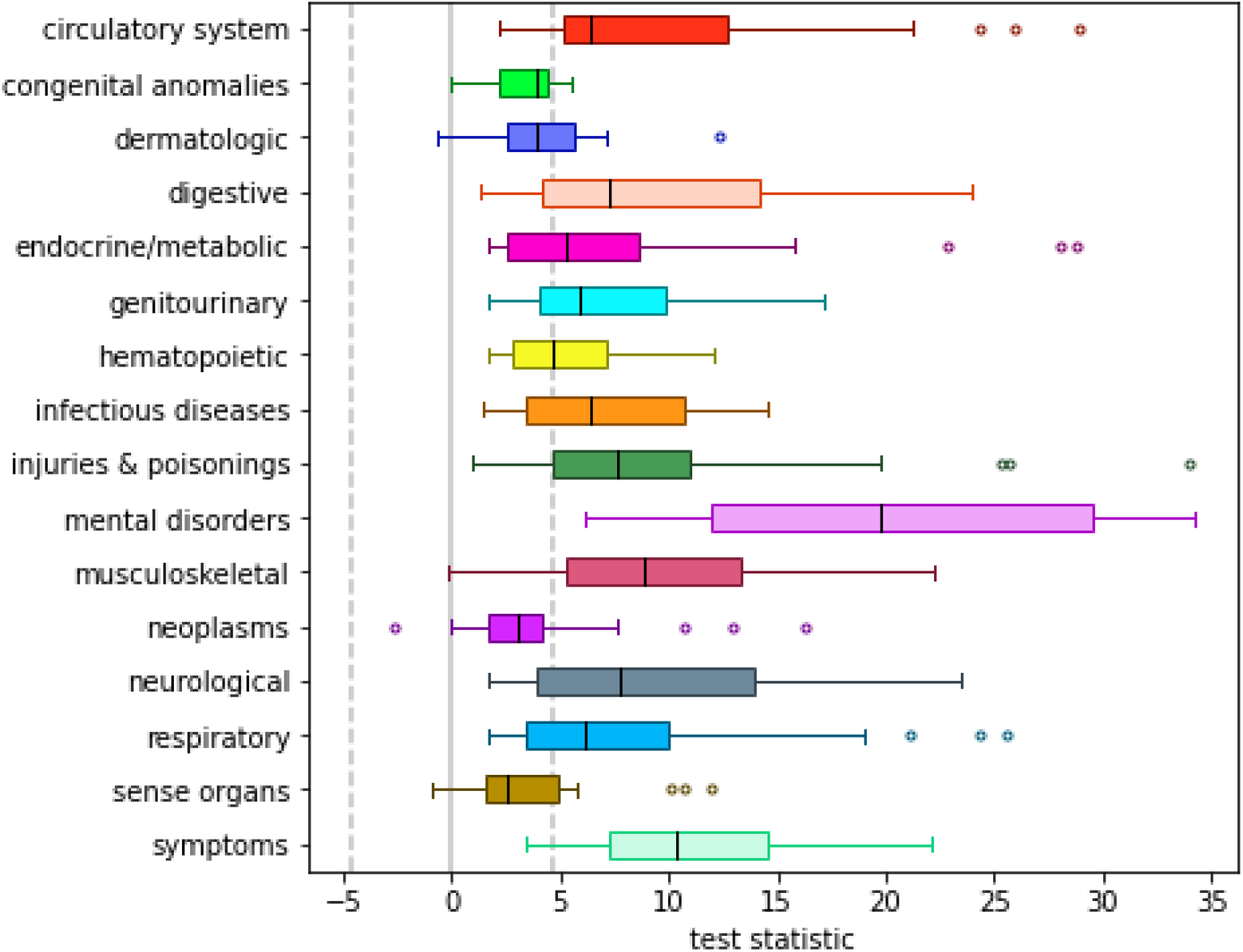
Factor 9 associations across top-level inpatient diagnostic phecodes. Box-and-whisker plots are shown for associations with 403 derived medical phecodes grouped by category. Boxes represent the middle quartiles, with whiskers extending to maximum and minimum observed values, excluding outliers >1.5x the interquartile range away from the middle quartiles while are plotted individually. Median values per category are indicated by individual black lines inside the boxes. The dotted grey lines represents the critical test statistics for significance at p<0.05 once correcting for comparisons across all 403 phecodes.

Factor 9 shows strong genetic correlations with prior GWAS of trauma exposure (r_g_=0.93[0.02]; (42)), childhood maltreatment (r_g_=0.81[0.02]; (43)), and PTSD (r_g_=0.75[0.07]; (44)). It has moderate genetic correlations with external GWAS of psychiatric (e.g., schizophrenia r_g_=0.35[0.03]; (45)) and substance use (e.g., cannabis use disorder r_g_=0.45[0.05]; (46)) outcomes (**Fig S6**). Of the 8 genome-wide significant loci, 4 have been previously identified in GWAS of trauma phenotypes (42,43). The remaining 4 loci novel to trauma associations have been identified in prior GWAS of psychiatric (47,48), behavioral (49,50), neural (51,52), and medical (53,54) outcomes (**Table S4**).

Overall, Factor 9 draws on the strength of relationships between multiple trauma indicators across the phenome, combining them in a data-driven way to demonstrate robust and ubiquitous associations with morbidity and mortality. FA thus facilitates a more global understanding of the importance of trauma for public health that would not be possible with individual indicators alone.

### Bundling correlated health behaviors boosts association power

Health behaviors represent a strong candidate for factor analysis, since multiple observed indicators likely reflect a generalized underlying tendency capturing individual differences. Factor 23, for example, includes traditional self-reported measures of exercise frequency and duration, as well as other physical and social activity measures like spending time outdoors, playing sports, and walking for pleasure (**Table S1**). It also incorporates dietary items such as fruit, vegetable, and oily fish intake, likely reflecting broader correlations across pro-health behaviors. Modifiable health behaviors such as these have been shown to mitigate adverse medical outcomes, with inactivity accounting for 1% of disability-adjusted life years lost globally (55). Indeed, Factor 23 has strong correlations across all categories of medical phecodes and a substantial association with prospective survival (HR=0.77[0.76-0.79]; **Fig 2a**).

Analyses linking Factor 23 to other biobank data identify clearer associations than comparable analyses using any individual top component item. Prospective analyses of all-cause mortality, for example, show weaker effects for each of Factor 23’s 10 top-loading items (incremental pseudo-R^2^=1.27 × 10^−3^ for factor vs. 6.75 × 10^−10^ to 1.12 × 10^−3^ for individual items). An unweighted sum of z-scores across these top items is also more weakly associated with survival (incremental pseudo-R^2^=1.00 × 10^−3^) than the weighted sum used by the factor scores. Even with optimal weighting, at least 7 items are necessary to capture 80% of total factor variance, suggesting that signal in Factor 23 relies on the ability to summarize information across related items.

This stronger signal enables GWAS of Factor 23 to identify the largest number of significant loci to date for a self-reported measure of physical activity. Of 34 loci, 24 are not significant in GWAS of the top 5 items, and 25 have not previously been identified in GWAS of self-reported physical activity, although 14 were significant at a less stringent threshold (i.e., p<5×10^−5^; (56)). Similarly, consistent with the trend across the factors, the SNP heritability of Factor 23, 0.07(0.003), is higher than most of its component items and sum of top-item z-scores (h_g_^2^=0.05 [0.004]). This increase in power likely comes as a result of leveraging the correlated structure across items and moving toward a more continuous construct measure. Furthermore, heritability for Factor 23 is enriched for regions of the genome associated with central nervous system cell types (p=2.52×10^−9^; **Fig S5**), indicating that physical activity is primarily influenced by brain and behavior, rather than traits like muscle tone or cardiovascular health.

These results suggest that pro-health behaviors such as physical activity can be more powerfully studied by considering many indicators. Factor 23 successfully leverages the correlated structure of its component items in a principled yet hypothesis-free manner to enable novel discoveries that can begin to differentiate individuals based on their engagement in physical activity.

### Parsing subdomains of socioeconomic status

Socioeconomic status (SES) is one of largest single predictors of health and mortality (57) and, for research purposes, is traditionally estimated with indicators of education, occupation, and income (58). Three of the factors predominantly include SES-related variables, and reflect correlates of occupation (Factor 5), educational attainment (EA; Factor 10), and social and economic stability (Factor 15), respectively, across the phenome. These factors pull from a range of questionnaires, from diet to employment history to social support, capturing items both traditionally and non-traditionally considered SES indicators (**Table S1**). Factor 10 includes classic SES items like educational attainment and job codes, as well as apparent cohort-specific correlates like intake of ground coffee/espresso, muesli, and wine. Factor 5 captures jobs such as low-ranking military, physical labor, and factory occupations, as well as work environments full of fumes or noise. Finally, Factor 15 reflects social and economic stability, including social support networks, loneliness, home ownership, household income, and having never been divorced.

The SES factors are pervasively, yet differentially, associated with health outcomes in UKB (**Table S3**; **Fig S3**). Factors 5 and 10 are much more similar to each other in their patterns of association to linked hospital inpatient phecodes (Pearson correlation of regression betas |r|=0.61) than they are to Factor 15 (|r|=0.24-0.33). Factors 5 and 10 are most distinguished by differential associations with respiratory and cardiometabolic diseases, with Factor 5 being more associated with respiratory diseases and Factor 10 with cardiometabolic. Factor 15, in contrast, is much more protective against hospitalization for mental health and substance use disorders than the other two factors. Factor 15 therefore suggests a distinct domain of SES that is protective against the “diseases and deaths of despair” that have been shown to be the most significant drivers of decreasing life expectancy in recent years (59,60). In fact, Factor 15 is the most prospectively predictive of survival of all 35 factors (HR=0.75[0.74-0.76]; **Fig 2a**).

Genetic data provides additional insight into how the factors separate multiple axes of SES. Genetic associations with SES are often correlated with environmental influences, and thus cannot be interpreted as purely causal. However, we can use genetic information as a tool to inform the understanding of these domains. For example, genetic correlations across the SES factors are low to moderate, suggesting partially overlapping yet distinct domains of SES (**Fig 5**). Factor 10 shares substantial genetic overlap with prior GWAS of EA (r_g_=0.93[0.01]; (61)) and its correlates, including household income (62) and cognitive performance (63). Notably the SNP heritability of Factor 10 (h_g_^2^=0.21[0.01]) is greater than that of this most recent EA GWAS (h_g_^2^=0.12, (23)), reflecting a potential benefit of measures that include correlated measures of traditional and nontraditional SES. Factor 5, in contrast, reflects strong and roughly equal genetic overlap with EA, region-based social deprivation (64), and household income. Factor 15 is the most distinct SES factor, with moderate genetic correlations with social deprivation and household income, but low and nonsignificant associations with EA and cognitive performance, respectively. Building on recent work (64), Factors 5 and 15 are associated only with genetic effects that operate through the inherited family environment, while Factor 10 is also associated with direct genetic effects (65).

**Fig. 5.**
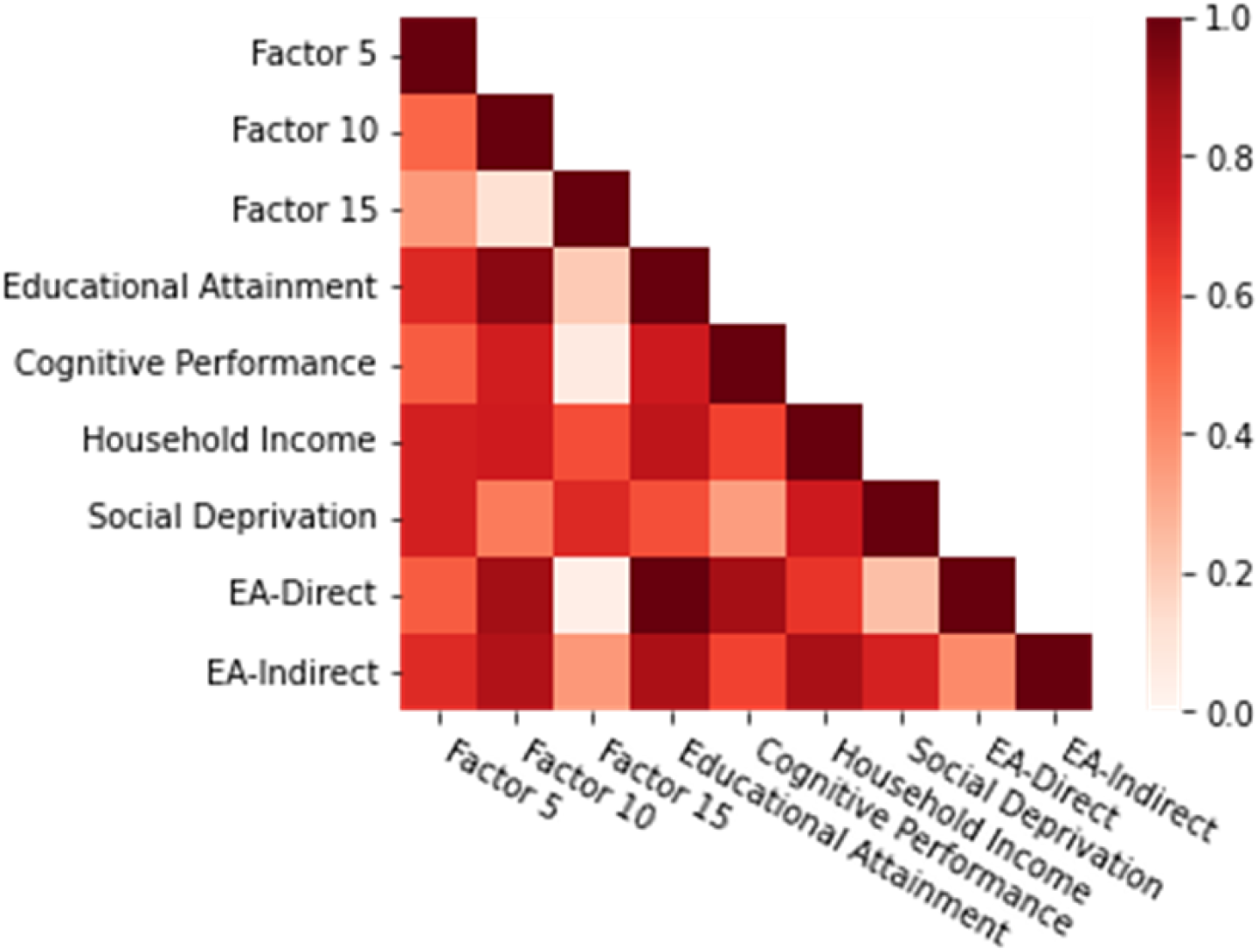
Genetic overlap across factors in the SES domain and prior GWAS of SES indicators. All genetic associations are flipped to be in the direction reflecting greater SES for consistency. Color of each box within the heatmap indicates the strength of genetic overlap across the two corresponding phenotypes. EA: Educational Attainment.

While the allocation of items among factors reflects the variables measured in UKB, the UKB environmental context, and the modeling approach, these highly powered results confirm that SES is a multidimensional construct. Factors 5, 10, and 15 offer one possible set of axes for this space, and demonstrate the value of disentangling aspects of SES to parse their complex associations across health and disease.

## Discussion

The results of this study demonstrate that by applying a principled, model-based data reduction technique, large-scale biobank data can be distilled in a way that is sensible and digestible. The factors that we extract from UKB link causes, correlates, and consequences of health, behavior, and disease at the cohort level, revealing sweeping patterns that are otherwise obscured by the sheer volume of data. Apart from increases in heritability and power that can aid in genetic discovery, we use the factors to recapitulate known disease nosology, to highlight the importance of psychiatric constructs for public health outcomes, to combine indicators to better stratify individuals across major axes of behavioral variation, and to parse subdomains of socioeconomic status. As the first factor analysis attempted across multimodal data at biobank scale, the results provide a proof-of-concept that this approach can return both sensible and insightful relationships.

For researchers interested in large-scale patterns across the human phenome, these results provide a critical advance. Without sacrificing depth or interpretability, FA allows researchers to better analyze the fully correlated human phenome. Since FA is dataset-specific, analyses of this nature in other biobanks can highlight the patterns of variation that matter for health in different sociopolitical, cultural, and diagnostic contexts.

The results are subject to several important caveats (see **Supplementary Text** for additional discussion). First, latent constructs identified via factor analysis are not “real”; they are simply statistical relationships, or the weighted linear combination of items that capture the most variance-covariance from the overall dataset. Factors are therefore non-trivially dependent upon the nature of the dataset used, including the variables measured, the characteristics of the participants, and the sociodemographic context in which the data were collected (66). In these analyses, we utilize UKB participants of predominately estimated European genetic ancestry, thus limiting the generalizability of the results outside of that population. Second, the structure and complexity of data at this scale forced us to require orthogonality across the factors. As such, the variance captured by each factor is independent. Caution must therefore be exercised in interpreting relationships within and between factors, since this independence is an artifact of the modeling approach. Nevertheless, with proper care these orthogonal factors can allow insight into the complex relationships among the set of items spanned by the factor analysis. While factors themselves should not be thought of as risk metrics as they conflate causes and consequences at a particular cross-section in time, they can inform development of future measures by identifying items central to a construct.

In sum, we perform the largest phenotypic factor analysis to date, distilling the full range of traits in a single biobank into a tractable number of interpretable constructs. This approach provides an important first step toward better embracing the full and complex measured phenome to power discovery for human health and wellbeing.

## Materials and methods

### UK Biobank cohort

#### Sample selection and quality control

Starting from the 487,409 genotyped participants in the UK Biobank second round release, we first subset to unrelated individuals with low autosomal missingness rates used for principal components analysis (PCA) by Bycroft et al. (4). We adopt this selection to aid consistency with other UK Biobank applications and analyses. We then restrict the cohort to individuals of predominantly estimated European genetic ancestry based on analysis of the top six principal components (PCs). This selection was intended to be more broadly inclusive than the “white British” criteria used by the UK Biobank genetics team (4) while still restricting to a sufficiently homogenous and unrelated set of individuals to permit GWAS with conventional linear regression. After ancestry selection, we make final exclusions for individuals who withdrew from UK Biobank participation prior to the GWAS analysis and individuals who were omitted from imputation phasing (e.g. individuals with sex chromosome aneuploidies). After all sample QC, there are 361,194 QC positive individuals. Between initial QC and the start of analyses for the current study, an additional 50 participants withdrew from UKB, resulting in a final N of 361,144.

#### Phenotype curation

A core challenge to the analysis of such a wide range of phenotypes as those available in the UK Biobank is the curation and harmonization of the large number of variable scalings, categorizations, and follow-up responses. To automate this process, we used a modified version of the PHEnome Scan ANalysis Tool (PHESANT (67)). Unlike standard PHESANT, the modified version does not perform association analyses, but simply generates a collection of re-coded phenotypes.

The incorporation of new phenotypes requires careful examination of raw data codings and, in the case of binary phenotypes, consideration of control definition. Re-codings of variables, and inherent orderings of ordinal categorical variables, are defined in the data-coding file, available in the GitHub repository https://github.com/astheeggeggs/PHESANT. We restrict the phenotype data to those that belong to individuals in the unrelated GWAS subset, and run the modified version of PHESANT on the phenotypes in the UK Biobank application. For continuous phenotypes, we retain the raw version of the continuous phenotype, with no transformation applied to the data. We processed 3,011 unique phenotypes using PHESANT. For all binary phenotypes, we require a minimum case count of 100.

Along with these PHESANT-curated phenotypes, we also processed 633 ICD10 disease codes, treating all individuals with a specific ICD10 code as cases, and the remaining UK Biobank sample as controls. Curation of the ICD10 codes was carried out separately for computational efficiency. For the ICD10 phenotypes, individuals are assigned a vector of ICD10 primary diagnoses. We truncated these codes to the three-digit category level, and assigned each individual to either case or control status for that ICD10 code in turn by checking if their vector of primary diagnoses contains that code. Throughout, we assume the data contains no missingness, so the sum of cases and controls is the number of individuals in the European ancestry subset of the UK Biobank data. Consistent with our treatment of binary phenotypes, ICD10 code case/control phenotypes are removed if less than 100 individuals in the European ancestry subset had a given phenotype as a primary diagnosis.

### Factor analysis modelling

The factor analysis model treats observed variables *X* as measures of a smaller number of unobserved latent factors *F*, with corresponding effect sizes, or “loadings,” Λ, and item-specific residuals *ϵ* (11–14). Specifically, let

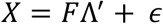

where *X* is the *n* × *p* matrix of *p* centered and standardized observed variables for *n* individuals, *F* is the *n* × *t* matrix of *n* individuals’ values for *t* latent variables, and Λ is the *p* × *t* matrix of the effects of the *t* latent variables on the *p* observed variables. This model is easily extendable to the inclusion of “nuisance” covariates, either by explicitly adding terms for covariates or residualizing them out of *X*. For the purposes of our analyses, we chose the latter approach, which is described in later sections.

We are able to fit this model by considering the observed covariance across items.

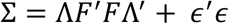

Assuming that the observed covariance between the items is fully explained by the latent factors, we denote Ψ = *E*[*ϵ*^′^*ϵ*] as the *p* × *p* diagonal matrix of residual variances per item (i.e., item uniquenesses). We additionally choose to model the latent factors as independent and fix their scale to have unit variance, allowing us to model the observed covariance between the items with the matrix decomposition

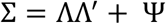

Multiple methods exist for the extraction of latent factors which generally yield similar results; we consider multiple such methods below. Once factors are extracted, however, there are an infinite number of equivalent Λs up to a particular rotation; this is referred to as “rotational indeterminacy.” To uniquely fit the model additional optimization criteria must be specified for Λ. For the current analysis we rely on the “varimax” rotation, one of a number of standard rotations that encourages sparsity, or a “simple structure,” on the model by penalizing factors or items with multiple large loadings. This rotational restriction facilitates interpretability, as the majority of the signal associated with a particular factor can be identified based on a limited number of its top-loading items.

In the current study, we undertook a multi-stage approach to best meet the assumptions of the factor analysis methodology, while adapting it for such large-scale data (**Fig S1**). We first identified a core data group consisting of 42,325 individuals and 898 items from which a stable pairwise correlation matrix could be estimated across a range of questionnaires and assessments. We split this group into modeling (N=33,860) and holdout (N=8,465) subgroups based on an 80:20 split. After systematically removing colinear items from the dataset (730 items remaining), we performed an exploratory factor analysis (EFA) within the modelling subgroup in order to determine the factor structure (i.e. the number of latent factors *t* and which elements of Λ are non-zero). We then further refined the model suggested by the EFA utilizing a structural equation model for confirmatory factor analysis (CFA) in the same modelling subgroup. We tested the fit of this final model, with constrained parameters, in the holdout sample. These steps are described in more detail below, as well as in the **Supplementary Text**.

#### Core data group

To enable the estimation of reasonably unbiased pairwise correlations between variables, we began with all individuals of European ancestry (N=361,144) and all phenotypes analyzed in both sexes in the initial release of the Neale Lab UKB Round 2 mega-GWAS (2,772 phenotypes; https://www.nealelab.is/uk-biobank/ukbround2announcement). We first identified a core group of individuals with a high rate of assessment completion (N=42,325; see **Supplementary Text: Selection of individuals for core data group**). We then identified items with low missingness in this core group, sufficient prevalence (>1%), and non-structured and non-item-dependent missingness, from which pairwise correlations could be successfully estimated (898 items; see **Supplementary Text: Item selection for core data group**). The overall missingness rate in this final core data group was 9.1%, with missingness on each item of up to 28.6% (SD: 10.7%), and for each individual up to 33.3% (SD: 7.9%). See **Supplementary Text: Characteristics of core data group** for more information about demographic and item composition.

From this core data group, we systematically removed collinear items to improve the stability of factor analysis estimation. Starting with a Pearson correlation matrix residualized for our chosen “nuisance” covariates (i.e., first 20 genetic PCs, age, chromosomal sex, age^2^, age-x-chromosomal sex, and age^2^-x-chromosomal sex; see **Supplementary Text: Selection of “nuisance” covariates**), we removed items that were redundant between observed and derived items (67 items), highly correlated with missingness (6 items), had pairwise correlations r>0.95 (43 items), squared multiple correlation (SMC) >0.98 (43 items), or had correlation induced by “None of the above” response categories (6 items). After these exclusions, 730 items remained for factor analysis.

Finally, this core group was further divided into modelling (N=33,860) and holdout (N=8,465) groups based on an 80:20 split to avoid bias from overfitting in evaluation of model fit.

#### Exploratory factor analysis

We performed an exploratory factor analysis (EFA) using the “psych” package (68) in R (version 4.0.2) on a partial pairwise Pearson correlation matrix--residualized for the first 20 genetic PCs, age, chromosomal sex, age^2^, age-x-chromosomal sex, and age^2^-x-chromosomal sex--within the modelling subgroup (N=33,860) in order to determine the factor structure.

Conventional methods for selecting the number of latent factors provided inconsistent results for the data: the scree plot suggested 30 – 50 factors (**Fig S8**), parallel analysis suggested 177 factors, and 253 eigenvalues of the correlation matrix were >1. We explored factor solutions with an increasing number of factors using WLS (weighted least square), GLS (generalized weighted least square), MINRES (minimum residual) and ULS (unweighted least square), all with “varimax” rotation to extract orthogonal factors (**Fig S8, S9**). As an upper bound, ultra-Heywood cases were observed when fitting more than 169 factors with GLS, 186 actors with WLS, or 38 factors with ULS or MINRES. Inspection of these models found that WLS and GLS yielded many factors with strong loadings (>0.3) for at most 1 item, while the ULS or MINRES solutions provided more interpretable results with factors incorporating variation from multiple items.

Based on stability and interpretability, we selected a 36-factor MINRES solution (MINRES-36; RMSR = 0.02, variance explained = 18.5%; **Fig S9d**) as our preferred EFA model for subsequent refinement.

#### Confirmatory factor analysis

We further refined the model suggested by the EFA utilizing a structural equation model in the same modelling subgroup. This confirmatory factor analysis (CFA) allows us to test the fit of a more parsimonious model (i.e., omitting loadings with small estimates in the EFA) while more appropriately modelling the covariance structure of the diverse variable types (i.e., binary, ordinal, continuous) and with more robust handling of missingness.

For the 564 variables from the EFA with loadings >0.1 on at least one factor (see **Selection of minimum loading for factor inclusion**), missing data was imputed using classification and regression trees (CART) within the Multivariate Imputation by Chained Equations (MICE) package (69) in R, with all covariates as well as 20 additional auxiliary variables (e.g., previously excluded “gatekeeper” items and assessment center) included as predictors. Items whose missingness pattern depended on the target item’s missingness were omitted as predictors for that target item. Evaluation of this approach using synthetic missingness at random (MAR) and completely at random (MCAR) showed good convergence and minimal systematic bias.

Confirmatory factor analysis models were fit to the imputed data for the modelling group (now N=33,854 due to participant withdrawals during the course of the study) using structural equation modeling using an extensively modified version of the lavaan package ((70); version 0.6-3) in R (see **Supplementary Text: Computing aspects of structural equation modeling**; adapted code is available via github [LINK TO BE ADDED UPON ACCEPTANCE]). Correlations between variables were estimated as appropriate for their measurement scale (e.g., polychoric for pairs of ordered variables, Pearson for pairs of continuous variables, and polyserial for pairs containing one of each), assuming an underlying normal distribution. Continuous variables were standardized prior to modeling, and all variables were modelled conditional on exogenous “nuisance” covariates (i.e., first 20 genetic PCs, age, chromosomal sex, age^2^, age-x-chromosomal sex, and age^2^-x-chromosomal sex). Model parameters were estimated using diagonal weighted least squares (DWLS).

Fitting the EFA-derived model using CFA yielded a number of initial errors due to a lack of estimable pairwise correlation (e.g., due to collinearity) and cell sizes of 0 for ordinal variables. After removing 9 items to address these errors, 23 Heywood cases remained, indicative of overfitting and near collinearity. These new instances of collinearity were observed in part due to the lenient pairwise r>0.95 threshold used for the initial EFA and the addition of latent modelling of ordinal and categorical variables in the CFA. Items were iteratively removed until no negative residual variances remained (505 final items). Additionally, once these items were removed, one factor (Factor 8) overlapped completely with another (Factor 4) and was removed to facilitate model fitting (see **Supplementary Text: Differences between EFA and CFA**). **Table S6** documents the reason for each variable’s exclusion from the EFA to the final factor model.

Finally, we noticed that misfit in certain parts of the model was being driven by the presence of extreme outliers (see **Supplementary Text: Extreme outliers of continuous variables**). Therefore, we removed from analysis all individuals in the core group with values greater than 20 standard deviations from the mean on any continuous variable (N in modelling group = 52; N in holdout group = 13). This resulted in a final N of 33,802 in the modelling subgroup.

To evaluate the applicability of the factor model beyond the modelling subgroup, we obtained fit metrics in the validation holdout subgroup (initially N=8,465; N=8,452 after removing continuous-variable outliers) while constraining the model parameters (i.e., factor loadings) to those estimated in the training subgroup. Finally, fit of the model was assessed by the root mean square error of the approximation (RMSEA; values 0.01, 0.05, and 0.08 indicate excellent, good, and acceptable fit, respectively), standardized root mean squared residual (SRMR; values <0.08 indicate good fit), and comparative fit index (CFI; values >0.90 indicate good fit).

### Factor scores

Based on the final factor model, we then generated latent factor scores for each individual from the values of their observed indicator items (see **Supplementary Text: Factor score generation** for full details). For each individual *i* the estimated factor score for factor *t* is a weighted sum of the items *x*_*j*_

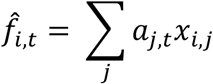

If we take the factor model as true, then the resulting estimates are of an individual’s “true” score for the underlying latent construct, otherwise they simply estimate the value that best approximates the observed data for each individual with the low rank approximation of the complete data modelled by the CFA. The current analysis uses two sets of factor scores, corresponding to two different estimation methods to compute the factor scoring coefficients *a*_*j,t*_, following previous recommendations to avoid biased results in factor score regression (71,72).

#### Dependent variable factor scoring coefficients

Where factor scores are used as the dependent variable in an analysis (e.g. GWAS), we 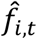 calculate using factor scoring coefficients computed with Bartlett’s method (73,74):

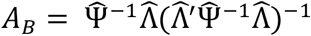

where: *X* is the *n* × *p* matrix of *p* residualized and standardized observed variables for *n* individuals, 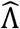 is the *p* × *t* matrix of estimated factor loadings; and (4) 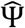 is the *p* × *p* diagonal matrix of estimated residual variances (i.e., item uniquenesses). Bartlett’s estimator is a weighted least squares solution that minimizes the residual variance of the items given the factor scores, weighting by the fitted item uniquenesses from the model.

#### Independent variable factor scoring coefficients

Where factor scores are used as the independent variable in the analysis (e.g. mortality), we use factor scoring coefficients computed with the Thomson-Thurstone (Regression) method (14,75),

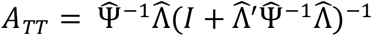

where *I* is an identity matrix. These factor scoring coefficients give the best linear prediction of the factor score, minimizing the sum of the expected mean squared error across factors.

#### Adjustments for categorical items

The above framework for the factor score estimators assumes that all of the item data *X* is observed and residualized for exogenous “nuisance” covariates. Although this is true for observed continuous items in the model, it is not true for the CFA model where the observed data is categorical and modelled through a link function.

To address the different measurement scale for categorical variables, we estimate the expected value of each individual’s latent continuous variable given the observed categorical item and the fitted probit regression with exogenous “nuisance” covariates. The residual between this expected latent value and the value predicted by the covariates is then substituted for the observed categorical variable in the factor scoring calculations.

The categorical variables will also have weaker covariances with the factor than the unobserved latent values modelled for the CFA loadings. To account for this attenuation, we transform the loadings and residual variances corresponding to categorical items for use in factor scoring (see **Supplementary Text: Modifications for categorical and missing data**). Specifically, we use

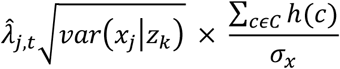

as loadings, where 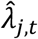 is the loading for item *j* on trait *t* estimated in the CFA, *h*(*c*) is the density of the standard normal distribution at the fitted probit threshold for each category *c* of the categorical variable, the variance of the categorical item conditional on the covariates *var*(*x*_*j*_|*z*_*k*_) is estimated empirically from the residual expected latent values described above, and the standard deviation of the categorical item *σ*_*x*_ is estimated from the class probabilities. Similarly, as the residual variance for the categorical items we substitute

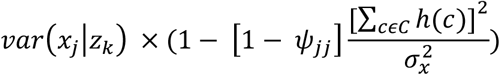

where *ψ*_*jj*_ is the estimated residual variance in the CFA and the remaining terms are defined as in the transformation of the loadings.

#### Adjustments for missingness

Factor scores estimates that sum across all items as described above cannot be computed for individuals with missing data. Instead, for each individual with a set of missing items *M* we compute factor scoring coefficients optimized for the subset of items that are observed, i.e.

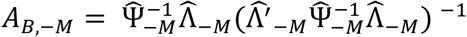

for dependent variable (Bartlett) factor scores and

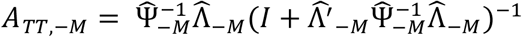

for independent variable (Thomson-Thurstone) factor scores.

The resulting factor score estimates maintain their desired relationship to other variables conditional on each missingness pattern, consistent with the bias-avoiding method of factor score regression, but the different amounts of information about the factor available for each individual leads to heteroskedasticity in factor scores across the missingness patterns. For analyses with the factor score as the dependent variable, we address this heteroskedasticity using weighted least squares (WLS) regression with estimated inverse-variance weights

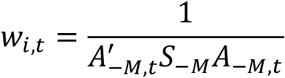

where S is the sample covariance matrix of pairwise complete observations after residualization for exogenous “nuisance” covariates. For linear regression analyses with factor scores as the independent variable we use Huber-White sandwich standard errors (76). Detail on motivation for each of these adjustments is provided in the **Supplementary Text**.

To further limit potential artifacts from heteroskedasticity related to structured missingness and ensure consistent interpretation of scores across UKB participants, individuals were included in factor score regressions if the factor score from their observed items was expected to correlate *r*^2^ ≥ 0.8 with what their estimated factor score would be with if all items were observed (see **Supplementary Text: Minimum correlation with complete data scores**; **Fig S10**). Because we observed this threshold to be more universally liberal in the independent than dependent factor scores, to allow for better concordance in samples across phenotypic and genetic analyses, we further restricted phenotypic analyses with the independent variable factor score to only those individuals included in the genetic analyses, resulting in sample sizes ranging from 75,226 (Factor 24) to 360,656 (Factor 20) (mean N = 252,219.571[121,829.646]).

#### Validation of factor scoring adjustments

To validate our factor-score-generating methodologies, we compared scores from our methods to those generated using a maximum-likelihood-based (ML) method in lavaan (70) for the core data group. Factor scoring was performed using the ML option in lavaan for all 10 multiple imputations of the core dataset. To test for phenotypic concordance across methods, we obtained Pearson correlation coefficients between factor scores generated using our method and the mean of those obtained in lavaan across all 10 imputations. GWAS for each factor for the scores generated with lavaan and with our dependent-variable factor scores. GWAS of the lavaan-generated scores were similarly conducted with WLS and estimated inverse-variance weights based on the observed variance in scores across individuals and across imputation replicates. Heritability and genetic correlation between the GWAS results from the two methods were compared using LD score regression (77).

### Phenotypic association analyses

To further characterize the latent factors and also reveal potentially interesting associations, we tested the independent-variable factor scores for associations with 403 top-level phecodes and 28 biomarkers in the UK Biobank, as well as with prospective mortality. Covariates for these phenotypic analyses included the first 20 genetic PCs, age, chromosomal sex, age^2^, age-x-chromosomal sex, age^2^-x-chromosomal sex, and dummy variables representing the assessment centers of origin.

#### Phecodes

Phecodes (1685 items) were taken from the Pan-UK Biobank pan-ancestry GWAS project (https://pan.ukbb.broadinstitute.org/) and were derived from ICD-9 and ICD-10 codes across a patient’s inpatient hospitalization records and, if applicable, death registry data. These diagnostic codes were mapped to descriptive phecodes using scripts from the University of Michigan (available at https://github.com/umich-cphds/createUKBphenome), which derived their mappings from PheWAS Catalog ((78–80); https://phewascatalog.org/). Phecodes were filtered to have a minimum case count of 250 in the full EUR sample, with a minimum of 25 cases per chromosomal sex, leaving 940 for analysis. Given the nested nature of the phecodes, such that top-level codes contain all diagnoses listed in subcodes, we restricted analyses to the 403 remaining top-level phecodes only. Associations were performed using a generalized linear model with a binomial link function and Huber-White (“HC0”) robust standard errors (76,81) using the statsmodels package ((82); version 0.13.1) in Python.

#### Biomarkers

Serum biomarkers were obtained from the UK Biobank (28 items, after excluding rheumatoid factor and oestradiol for known QC issues (83)). Associations between each independent-variable factor score and biomarker were performed using ordinary least squares regression and Huber-White (“HC0”) robust standard errors (76,81) using the statsmodels package ((82); version 0.13.1) in Python. Due to known issues with sample dilution (83), an additional covariate was included representing the estimated serum sample dilution factor.

#### Mortality analyses

Given that the factors from our analyses represent major axes of measured phenotypic variation in the UK Biobank, it is plausible that they would be differentially associated with downstream mortality. We therefore performed cox proportional hazards regression (84) to assess relative risk of mortality across individuals based on independent-variable factor scores.

Since surveys and assessments were administered at different times, to avoid issues of immortal time bias, *T*_0_ was defined as the last contact an individual had with the UK Biobank study, within the context of the items included in the factors. Of the items included in the final factor model, the differently-timed assessments included: baseline, a maximum of five 24-hour diet follow-up questionnaires, a work environment questionnaire, and a mental health questionnaire. For example, if an individual completed the baseline assessment, mental health questionnaire, and a 24-hour diet follow-up questionnaires, their *T*_0_ would be their most recent questionnaire completion date. We included several “continuously-updating” items within the factors (e.g., primary ICD-10 codes and items relating to hospital stays, which are updated based on linked inpatient hospital records). Therefore, for the mortality analyses, we recoded each individual’s factor scores on factors containing these “continuously-updating” items with their values at *T*_0_. If a person’s first instance of a primary ICD-10 code was dated after their *T*_0_, they would be recoded and rescored as *not* having that code.

Date of death was obtained with linked death registries, and analyses were right-censored to the earliest recommended censoring date across the death registries specified by UKB for England & Wales and Scotland (i.e., 30 Sept 2021 at the time of analyses). In addition to the standard phenotypic analysis covariates, a covariate was added representing days from baseline assessment to *T*_0_. Analyses were performed using the lifelines package ((85); version 0.26.4) in Python.

### Genome-wide association analysis

#### Variant QC

Over 92 million imputed autosomal and X chromosome variant dosages were available in the UK Biobank release. The variant QC process focused on using widely adopted GWAS QC parameters to retain high quality variants. After restricting to the 361,194 QC positive individuals, we retained SNPs with minor allele frequency (MAF) > 0.001, Hardy-Weinberg Equilibrium (HWE) *p*-value > 1e-10, and imputation INFO score > 0.8. INFO scores were taken directly from the UK Biobank SNP manifest file. The only exception involved SNPs annotated as having protein-truncating or missense consequences (from Ensembl VEP consequence annotation), where we relaxed the cutoff to MAF > 1e-6. After variant QC, 13,364,303 autosomal variants were retained for association analysis.

#### GWAS model and implementation

GWAS of the dependent-variable factor scores were performed in Hail (https://hail.is/) using weighted least squares regression. Variance weights were calculated as described in the **Factor score generation** section above. To limit the impact of structured missingness and ensure consistent interpretation of scores across individuals, individuals were included only if their score based on their missingness pattern explained 80% of variance in a hypothetical observed factor score for which no items were missing (**Fig S10**). Covariates included the first 20 genetic PCs, age, chromosomal sex, age^2^, age-x-chromosomal sex, age^2^-x-chromosomal sex, and dummy variables representing the assessment centers of origin. Post-association test statistics were corrected for LDSC intercept to reduce potential impacts of stratification. Effective sample size for genetic analyses, taking into account missingness, was calculated as the sum of each person’s inverse variance weight divided by the regression weight assigned to a hypothetical person with 0 missingness, and ranged from 74,782 to 359,419; (mean=236,980.029[112899.969]).

#### Identification of significant independent loci

We used the FUMA (86) pipeline to identify independent genomic loci. We considered an independent locus as the region including all SNPs in pairwise linkage disequilibrium (LD; r^2^ > 0.6), with the lead SNPs in a range of 250 kb from each other and independent from other loci at r^2^ < 0.1. We used the 1000 Genomes Phase 3 Europeans reference panel (87) to determine LD.

#### Comparison of factor vs. item GWAS

To investigate the genetic properties of the factors, we compared factor GWAS to GWAS of component items. Summary statistics for all 505 items included in the final factor model were publicly available via the Neale Lab UKB Round 2 mega-GWAS (https://www.nealelab.is/uk-biobank/ukbround2announcement), and h_g_^2^ estimates were downloaded from the corresponding Heritability Browser (https://nealelab.github.io/UKBB_ldsc/index.html)

Independent loci for the 5 top-loading items per factor were identified using a local version of FUMA (86) and identical parameters to those used to define the factor GWAS loci. Given that the intention for these analyses was to compare loci across top items and corresponding factors, loci were henceforth defined by their basepair intervals, and loci across GWAS (e.g., for a factor and its top item) were combined if their basepair intervals overlapped.

#### LD Score Regression analyses

LD score regression analyses of heritability, enrichment (88), and genetic correlation (77) were performed using LDSC ((77); available at https://github.com/bulik/ldsc) with LD scores computed in individuals of European genetic ancestry from the 1000 Genomes Project. All analyses were performed with default settings except where otherwise indicated.

#### SNP Heritability

SNP-based heritability was estimated on the observed scale for each factor using stratified LD score regression (S-LDSR) (88) and version 1.1 of the baseline-LD model ((89); available at https://alkesgroup.broadinstitute.org/LDSCORE/). We use this stratified model to more robustly fit variation in genetic signal across the genome, estimating per-SNP heritability conditional on 75 annotations, including functional categories, evolutionary constraint, histone marks, and LD- and allele frequency-related annotations. The default filter in ldsc for maximum chi square was omitted to avoid truncating top hits at our large sample size.

#### Cell type enrichment analyses

To gain insights into the underlying biology of the factors, we evaluated heritability enrichment of regions of the associated with cell-type specific chromatin marks using S-LDSC (88) and annotations derived from the Roadmap Epigenomics Consortium (90), as described in Finucane et al. (88). To reduce multiple testing burden and give a broader summary of systems-level biology, we grouped the updated cell type–specific annotations described in (91) into 9 tissue groups (Adipose, Blood/Immune, Cardiovascular, Central Nervous System, Digestive, Liver, Musculoskeletal/Connective, Pancreas, and Other) following the same procedure described in (88), taking the union of annotations belonging to each group. Consistent with recommendations from Finucane et al. (88), only factors with strongly significant heritability estimates (z>7) were included in these analyses.

## Data Availability

All data, code, and summary statistics produced in the present study are available upon reasonable request to the authors. Access to individual level data from the UK Biobank can be obtained by bona fide scientists through application to UK Biobank (https://www.ukbiobank.ac.uk/enable-your-research). Summary statistics for item-level GWAS are available as part of the Neale Lab UKB Round 2 Mega-GWAS (http://www.nealelab.is/uk-biobank/ukbround2announcement).

https://www.ukbiobank.ac.uk/enable-your-research

## Acknowledgements

CEC and EBR are supported by National Institute of Health (NIH) grant R01MH124851 and the Stanley Center for Psychiatric Research. RW received funding and support as an AnalytiXIN scholar from AnalytiXI Indiana. GDS works for the MRC Integrative Epidemiology Unit at the University of Bristol, which is supported by the Medical Research Council (MC_UU_00011/1). BMN is supported by NIH grant 5R37MH107649.

We thank S. Hyman, R. Hosking, B. Domingue, M. Nivard, and T. Ulrich for carefully reading and commenting on the manuscript. We also wish to thank members of the Neale and Robinson Labs, the Stanley Center for Psychiatric Research at the Broad Institute of MIT and Harvard, and the Analytic and Translational Genetics Unit at Massachusetts General Hospital for helpful discussions and feedback. This research was conducted by using the UK Biobank Resource under application 31063. We thank all cohort participants for making this study possible.

## Author Contributions

Conception: CEC, RS, BMN, RKW, RBR

Design: CEC, RS, AE, MK, DH, DK, BMN, RKW, EBR

Acquisition: CEC, DSP, MK, LA, KJK, SCB, CMC, CC, DPH, BMN, RKW, EBR

Analysis: CEC, RS, DP, MK, LA

Interpretation: CEC, RS, RW, AE, GDS, BMN, RKW, EBR

Software: CEC, DSP, LA, PS, DK

Manuscript: CEC, RW, BMN, RKW, EBR

