## Supplementary Text for "Principled distillation of multidimensional UK Biobank data reveals insights into the correlated human phenome"

### Contents


Selection and composition of core data group ... 3
  Selection of individuals ... 3
  Selection of items ... 5
  Characteristics of final core data group ... 6
Selection of items to omit for collinearity ... 8
Exploratory factor analysis ... 10
Confirmatory factor analysis ... 11
  Selection of minimum loading for factor inclusion ... 12
  Multiple imputation of core data group ... 13
  Computational implementation of structural equation modeling ... 15
  Iterative model refinement ... 15
  Constrained confirmatory factor analysis ... 17
Differences between EFA and CFA ... 17
Factor score generation ... 20
  Bartlett estimator ... 21
  Thomson-Thurstone estimator ... 23
  Modifications for categorical and missing data ... 25
    Measurement scale for categorical variables ... 25
    Transformed loadings ... 27
    Transformed residual variances ... 29
    Estimating factor scores with missingness ... 30
    Heteroskedasticity of Bartlett factor scores by missingness pattern ... 32
    Missingness-based heteroskedasticity and bias for Thomson-Thurstone scores ... 35
    Minimum correlation with complete data scores ... 37
  Validation of factor scoring methods in the core data group ... 39
Notes on modeling and limitations ... 41
  Impact of input items on factor structure ... 42
  Impact of questionnaire structure ... 43
  Extreme outliers on continuous variables ... 45
  Modeling of non-continuous items ... 46
  Forced orthogonality between factors ... 47
  Selection of “nuisance” covariates ... 48
  Assumptions of factor invariance ... 49
  Impact of structured missingness ... 49
References ... 52
Tables S2, S5, and S6 ... 55
Figures S1 to S15 ... 62

### Selection and composition of core data group

#### Selection of individuals

Factor analysis relies on the formation of a stable correlation matrix as input. However, the UK Biobank was not designed as a single survey measure. It instead consists of numerous self-report surveys, interviews, and assessments given across multiple timepoints (1). Missingness is introduced at the assessment level through selective response behaviors on the part of participants (e.g., electing to respond to a voluntary follow-up questionnaire), selective ascertainment (e.g., questions only asked of self-identified females or smokers), and later introduction into the UKB battery. The structured missingness introduced by this differential completeness is problematic for factor analysis, which relies upon the correlation matrix being consistent across individuals in the study population. This assumption is less likely to hold when pairwise elements of the correlation matrix are estimated from substantively different sets of individuals. To ensure sufficient pairwise overlap between individuals responding to different surveys and assessments in our core data group, we first considered inter- and intra-survey missingness, identifying a group of individuals who had usable phenotypic data across a wide range of assessments.

We first identified common patterns of structured missingness across individuals in UKB. Starting with the 2,772 phenotypes for which GWAS were performed across both sexes in the Neale Lab UKB Round 2 mega-GWAS (https://www.nealelab.is/uk-biobank/ukbround2announcement), those outcomes derived from a single question were collapsed into one item for the purposes of evaluating missingness patterns (e.g., in the case of categorical-multiple items, in which a participant could choose any number of responses from a list). We then assigned items to their most specific category based on the UKB tree structure (https://biobank.ctsu.ox.ac.uk/crystal/browse.cgi). To examine patterns of missingness, we then generated a null correlation matrix for all items. Within each category, all items with missingness correlations r>0.95 were merged. These collections of items with highly similar missingness patterns within each predefined category are called "questionnaires" in our analyses.

Of all the questionnaires, those that were asked only of a specific demographic subgroup of individuals (e.g., male- or female-specific questionnaires) or for which inclusion was conditional on a specific index event (e.g., cancer and death registries or the maternity questionnaire) were dropped. We then considered questionnaires with sample sizes of between 75,000 and 250,000; those with sample sizes of less than 75,000 would severely restrict the size of our core data group, while those with sample sizes greater than 250,000 were unlikely to require special consideration for missingness. Next, we dropped questionnaires with less than 5 items. This left us with 6 questionnaires: 1) the claudication and peripheral artery disease questionnaire from the touchscreen survey; 2) the mental health questionnaire from the online follow-up; 3) the work environment questionnaire from the online follow-up; 4) cognitive function measures; 5) physical measures (mostly: blood pressure measures, heel measures, and a hearing test); and 6) eye and vision measures.

Finally, we selected individuals who were missing at most 1 of the 6 questionnaires (N=42,325). This is done to ensure adequate overlap of individuals across questionnaires and also to help better ensure that questionnaire-level missingness was reasonably occurring at random.

#### Selection of items

After identifying a group of 42,325 individuals with sufficient completion of less commonly answered questionnaires, we then identified items for which pairwise correlations could be reasonably estimated, and for which missingness was reasonably occurring at random (e.g., was not dependent on an individual's would-be response value). In doing so, we eliminated items for which a response was dependent upon a response to a previous item, items with low prevalence, and items with relatively high missingness, even within the analytic core group of individuals.

Starting with the 2,772 items, we first removed those that were asked only of a specific demographic subgroup of individuals (e.g., male- or female-specific questionnaires) or for which inclusion was conditional on a specific index event (e.g., cancer and death registries; 113 items). We next removed all items with N<30,000 in our core data group (566 items), and, in the case of binary items, those with a prevalence of <1% in either the core data group or full EUR dataset (1,182 items).

We then formed a cross-item Pearson correlation matrix and identified pairs of items for which a correlation could not be estimated, likely indicating dependencies in missingness across the items. We sorted items by the number of "NaN" values they shared with other items, and we excluded the less prevalent item for any "gatekeeper" items or items that were dependent on other items. For example, since the vast majority of the sample reported having drank alcohol (core group: 97.6%; full group: 96.9%), we excluded the "alcohol drinker status" item for which only a small proportion of individuals reported not drinking, in favor of keeping the downstream questions on weekly alcohol intake. Conversely, since the vast majority of the sample was not adopted (core group: 98.8%; full group: 98.6%), we included the “adopted as a child” item and did not include the downstream questions about adoption. Resolution of these dependencies (removed 13 items) resulted in a final item count of 898. Below we provide more information about demographic characteristics and item composition for our core data group.

#### Characteristics of final core data group

The core data group was comprised of 42,325 individuals with a high rate of questionnaire completion, along with (1) 898 items with a high rate of completion among those individuals; (2) sufficient prevalence for binary variables (>1%) among both the core and full EUR data groups; and (3) no completely dependent inter-item missingness. The overall missingness rate was 9.1%, with missingness on each item up to 28.6% (SD: 10.7%), and for each individual up to 33.3% (SD: 7.9%). For individuals not within the core data group, item- and individual-level missingness was substantially higher, at 33.4% (SDs: 37.5% and 9.9%, respectively). Individuals in the core data group represented 10 of the 22 UKB assessment centers: Sheffield (N=8,155), Croydon (N=7,919), Hounslow (N=7,618), Birmingham (N=6,590), Liverpool (N=5,229), Middlesborough (N=3,071), Bristol (N=2,338), Nottingham (N=915), Swansea (N=398), and Wrexham (N=92).

Completion rates within the core data group for the 6 questionnaires on which the individuals were selected were as follows: 99.7% (N=42,205) for the claudication and peripheral artery disease questionnaire from the touchscreen survey; 89.8% (N=38,001) for the mental health questionnaire from the online follow-up; 74.7% (N=31,620) for the work environment questionnaire from the online follow-up; 100% (N=42,318) for the cognitive function measures; 100% (N=42,325) for the physical measures; and 79.8% (N=33,787) for eye and vision measures. A total of 18,631 individuals (44.0% of the core data group) completed all 6 questionnaires.

The causes of structured missingness for these different questionnaires differed: both the mental health and work environment questionnaires were online follow-ups to the original assessment, and participation was contingent on both providing a valid email address and opting in to these later assessments. In contrast, the 4 other questionnaires were added to the initial assessment later during recruitment, such that certain centers that completed recruitment earlier did not administer these questionnaires to any or all of their participants. Within the core data group, this is evident in the eye and vision measures questionnaire, which was not completed by anyone at 4 of the 10 assessment centers (i.e., Bristol, Nottingham, Middlesborough, and Wrexham), and by only 26.9% of individuals at the Swansea assessment center.

A plurality of overall items were taken from the initial touchscreen questionnaire (343 items), with a substantial number taken from the online follow-up as well (239 items, though these contained 159 employment history questions that included both raw and derived job codes). See **Table S5** for a full breakdown of items by category.

Individuals within the core data group were equivalent in age to those not included in the core data group (M=56.8 for both; t=0.556, p=0.578), but more were female (55.6% core group, 53.5% non-core group; $\chi^2$=66.412, p<0.001) and substantially more likely to report having completed college or university (45.7% core group, 30.7% non-core group; $\chi^2$=3816.0, p<0.001). These demographic shifts are consistent with prior literature demonstrating higher response and completion rates for females and individuals with higher educational attainment (2–4).

For our GWAS analyses, because of the differences between the core and full groups and because the factor analysis was modeled and tested exclusively on the core data group, we performed GWAS in both groups to allow for potential comparisons in genetic architecture between them. Though the main results presented in the paper are from the full EUR group, heritability estimates across the groups were generally concordant (range absolute difference: 0.001-0.051, mean=0.013(0.012)), and genetic correlations ranged from all high to very high (range: 0.864-1.101, mean=0.990(0.057); **Fig S11**).

### Selection of items to omit for collinearity

Though we identified a group of individuals with low *structured* missingness at the survey level, and a corresponding group of items with low missingness among this group, sufficient prevalence (>1%), and lack of cross-item missingness dependencies, additional adjustments needed to be made to facilitate the factor analysis algorithm. Specifically, issues of pairwise collinearity and multicollinearity needed to be resolved.

To address multicollinearity (e.g., perfect prediction of a variable by a combination of other variables), we first made sure that no item clusters existed for which both derived and component items were included. For example, items from a neuroticism questionnaire were originally included alongside a summed composite score; in this case, we removed the score in favor of keeping the items. Conversely, raw job codes were included alongside derived job codes, which combined multiple raw codes into larger “umbrella” categories. In this case, we kept the derived job codes, as prevalences for these items were increased relative to raw codes. A total of 67 items were removed from such item clusters.

We next removed items for which >50% of variance, once residualizing for covariates, was dependent upon the missingness of one or more items, likely reflective of structured missingness (6 items). This is similar to the above removals for inestimable correlations due to dependent missingness. For example, an item indicating whether or not a hearing test was performed was removed, as its variance was heavily dependent upon the missingness patterns of hearing test outcomes.

Next, starting with a Pearson correlation matrix residualized for our chosen “nuisance” covariates (i.e., first 20 genetic PCs, age, chromosomal sex, $age^2$, age-x-chromosomal sex, and $age^2$-x-chromosomal sex) in the modelling subset of the core data group (N=33,860), we identified all pairwise correlations r>0.95. For each of these pairs, one variable was removed (43 items); we prioritized keeping the variable with the least missingness in the core dataset.

We next identified items for which the squared multiple correlation (SMC) was >0.98. Most of these items had obvious reasons for near perfect prediction; for example, some phenotypes were derived from a single question in which a participant could select only one answer. One phenotype from each cluster of items was removed; in most cases we selected the phenotype with the lowest prevalence or least amount of variance (10 items). Three multicollinear item clusters remained for which survey response patterns were obviously not the cause: eye measures, body fat measures, and blood assays. A clinician was consulted in these cases, and items were removed until perfect prediction (SMC>0.98) stopped (33 items).

Finally, we identified 6 additional items that were “None of the above” responses to questions about otherwise unrelated illnesses of family members (e.g., Parkinson’s disease and depression). Given that such items could introduce multicollinearity and were not otherwise informative/meaningful, these items were removed from analyses. Resolution of all collinearity issues resulted in a final item count of 730 to be carried forward into the exploratory factor analysis.

### Exploratory factor analysis

We then performed an exploratory factor analysis (EFA) in the modelling subset of the core data group (N=33,860) using these remaining 730 items in order to determine the structure of the data. EFA was performed using the “psych” package (5) in R (version 4.0.2), using the partial Pearson correlation matrix of the 730 items as input. The number of factors in EFA is usually determined using scree plots, parallel analysis or by counting the number of eigenvalues of the correlation matrix that are greater than one. These three approaches provided inconsistent results for the data: the scree plot suggested 30 – 50 factors (**Fig S8**). Parallel analysis suggested 177 factors and 253 eigenvalues of the correlation matrix were >1.

To solve this problem, we devised a custom approach, with the goal to find a stable solution with interpretable factors. We explored factor solutions using the following factor extraction methods: WLS (weighted least square), GLS (generalized weighted least square), MINRES (minimum residual) and ULS (unweighted least square). In all steps the "varimax" rotation was used to find solutions with orthogonal factors. For each method the number of factors extracted was increased until mathematical error (ultra-Heywood cases, UH) indicated that the number of factors was possibly too large. This provided an upper limit for the maximum number of factors for each method: 169 for GLS, 186 for WLS, 38 for ULS and 38 for MINRES. The MINRES and ULS methods produced almost identical results and so we only used GLS, WLS and MINRES for further analyses.

For each factor extraction method, increasing the number of factors increased the variance explained by the model but at the same time produced "small" factors with one or no items having significant loading (magnitude > 0.3). **Fig S9** shows the distribution of the number of significant items in the factors for the different models. Because the 169-factor GLS solution and the 186-factor WLS solution produced many factors with only one significant item, we decided to pursue the more interpretable 38-factor MINRES model.

The 38-factor MINRES solution contained one "empty" factor, i.e., one factor with no item loading with magnitude > 0.3. A 37-factor MINRES solution also contained an empty factor. Reducing the number of factors again resulted in a 36-factor MINRES solution (MINRES-36, RMSR = 0.02, variance explained = 18.5%) with no empty factors (**Fig S9d**).

### Confirmatory factor analysis

Model refinement was carried out on an imputed version of the core data group (see **Multiple imputation of core data group**) using a confirmatory factor analysis framework in structural equation modeling using lavaan ((6); version 0.6-3) in R as a template, with extensive modifications (see **Computing aspects of structural equation modeling**). A confirmatory factor analysis tests the fit of a predefined model to the data. In contrast to an exploratory factor analysis, paths are constrained such that certain factors influence only certain prespecified observed variables. In the case, we wanted to test the fit of the model as defined by the EFA in the same modeling subset (now N=33,854 due to participant withdrawals during the course of the study) when modeling the covariance structure of all items appropriately and restricting the paths being estimated.

As in the EFA, we modelled all latent factors as orthogonal. Observed variables were assigned to latent factors if their loadings were >0.10 in the EFA (see **Selection of minimum loading for factor inclusion**). Continuous variables were standardized, and all variables were residualized for exogenous nuisance covariates (i.e., first 20 genetic PCs, age, chromosomal sex, $age^2$, age-x-chromosomal sex, and $age^2$-x-chromosomal sex). These "nuisance" covariates were treated as fixed parameters in the model. Variances of all observed and latent variables were fixed to 1. Correlations between variables were estimated as appropriate (e.g., polychoric for pairs of ordered variables, Pearson for pairs of continuous variables, and polyserial for pairs containing one of each), assuming an underlying normal distribution. Model parameters were estimated using diagonal weighted least squares (DWLS).

#### Selection of minimum loading for factor inclusion

To understand the range of meaningful factor loadings, we generated random data and compared the factor loadings for these random items to the loadings of the original 730 items. Three different kinds of random data were used: 1) Random normal ("rnorm" function in R, mean = 0, sd = 1), 100 repetitions; 2) Random binary traits ("rbinom" function in R, 100 repetitions each, p = 0.01, 0.05, 0.1, 0.2, 0.3,0.4, 0.5); 3) Random exponential traits ("rexp" function in R, rate =1, 100 repetitions). These data were corrected for covariates in the same way as the original 730 items, and combined correlation matrices were calculated with the main dataset. Once again 36 factors were extracted from these merged matrices using the MINRES method. In all cases the absolute value of the factor loadings of the random items were less than 0.03 (**Fig S12**). Based on these analyses we used a conservative cutoff of loading magnitude > 0.1 when considering items for inclusion for confirmatory factor analysis.

#### Multiple imputation of core data group

The core group dataset (N=42,325) was imputed using the Multivariate Imputation by Chained Equations (MICE) package (7) in R. Though the overall missingness of the core data group was low for items included in the CFA (9.3%), structural equation modeling of the dataset in lavaan required complete data, and we employed multiple imputation as a principled way to accomplish this complete data requirement.

We imputed all variables carried forward into the model refinement step (564 items; i.e., loadings >0.1 on a factor in the EFA). Of these 564 variables, 531 had missing values in the core dataset. To aid in imputation, we included all covariates, as well as 20 additional auxiliary variables (e.g., previously excluded "gatekeeper" items and assessment center) that could help explain patterns of missingness.

We chose classification and regression trees (CART) as an imputation method after testing this method and three other MICE methods appropriate for use with both ordered and continuous input variables; these included predictive mean matching, linear or logistic regression (as appropriate), and random forest on 104 variables purposefully selected to represent a range of data types and prevalence rates in the modeling subset of the core group. We used 10 iterations each to yield 5 imputations per method. Of the methods tested, CART demonstrated consistent (and relatively efficient) convergence, yielded values within the expected bounds of the observed values, and yielded mean values across imputations that were moderately correlated to the true, masked values (mean $r_{MAR}$=0.518[0.222]; mean $r_{MCAR}$=0.524[0.222]) in versions of our dataset with an additional 10% artificially induced missingness in either a missing at random (MAR) or missing completely at random (MCAR) pattern. There was little evidence of systematic bias using this method (mean standardized difference, MAR=-0.001[0.018]; mean standardized difference, MCAR=-0.001[0.017]). Importantly, across all pairs of variables, the correlations generated using datasets with artificially induced missingness imputed via CART were almost identical to those without such induced missingess for both MAR (e.g., for one of the 5 imputations: mean absolute difference = 0.009[0.011]) and MCAR (mean absolute difference = 0.010[0.012]).

Using the CART algorithm, we further restricted predictors for each item to those with a pairwise r>0.1 for either the target value or missingness pattern of the target item (for items with >50 individuals missing). These correlation matrices were constructed using the psych package's (5) "mixedCor" function in R, with functionality modeled after MICE's "quickpred" function. We additionally restricted predictors to those for which the missingness pattern was not dependent on that of the target item, with such dependence defined as items for which >50% of individuals missing the target item were also missing the predictor item. We generated 10 imputations of the core data group, with 10 iterations per imputation. Visual inspection of mean values across imputations revealed good convergence for all variables with any missingness.

#### Computational implementation of structural equation modeling

Structural equation modeling was carried out using lavaan ((6); version 0.6-3) as a template, with significant modifications made to achieve computational efficiency. Initially, portions of the method scaled quartically based on the number of items (in our case, 564). Leveraging matrix sparsity (e.g., using R's Matrix package (8), and R's bigstatsr package (9)), an optimized BLAS (i.e., Intel MKL BLAS), explicit parallelization (e.g., using R's doparallel package (10)), and reduction of linear algebra computations based on our specific use case, we were able to greatly reduce the computational and time complexity of the analysis. Adapted code is available via github [LINK TO BE ADDED UPON ACCEPTANCE].

#### Iterative model refinement

Fitting the EFA-derived model using CFA yielded a number of initial errors due to a lack of estimable pairwise correlation (e.g., due to collinearity) and cell sizes of 0 for ordinal variables. One of each pair of collinear items was removed (4 items), and a minimum cell size threshold of 25 within the core data set further resulted in the removal of 5 ordinal variables.

Following these changes, the EFA-derived model (minus the aforementioned variables) yielded 23 Heywood cases (out of 555), characterized by negative residual variance and indicative of overfitting. For the most part, these Heywood cases were related to one of two major issues: 1) pairwise collinearity (r>0.99) newly identified due to correct modeling of binary and ordinal variables; 2) multicollinearity due to the inclusion of variables indicating a response of “None of the above” to a certain question. For purposes of consistency and model stability, we removed all remaining “None of the above” variables (13 items; see **Impact of questionnaire structure**), as well as one of each pair of inestimable or r>0.99 correlations present in either the training or testing subgroup, regardless of whether those items were linked to a Heywood case (12 items).

Additionally, in looking at the remaining Heywood cases, it appeared that some were likely due to pairwise correlations just below the r>0.99 threshold, or unresolved multicollinearity across item clusters (i.e., smoking and traffic items). We removed one of each pair of items with a pairwise correlation just <0.99 that was likely causing a Heywood case (2 items), and removed 9 additional items due to multicollinearity. Finally, we observed that low cell counts across pairs of variables and/or covariates were further causing problems with estimating correlations; we thus removed 3 additional items.

After systematically removing these items (516 items remaining), 7 Heywood cases remained; the majority of these were due to pairwise correlations just below the r>0.99 threshold and item-cluster multicollinearity. Items were iteratively removed until no negative residual variances remained (11 items), leaving 505 items in the final model. Additionally, once these items were removed, one factor (Factor 8) overlapped completely with Factor 4 and was removed to facilitate model fitting (see **Differences between EFA and CFA**). **Table S6** documents the reason for each variable’s exclusion from the EFA to the final factor model. Finally, we noticed that misfit in certain parts of the model was being driven by the presence of extreme outliers (see **Extreme outliers of continuous**

**variables**). Therefore, we removed from analysis all individuals in the core group with values greater than 20 standard deviations from the mean on any continuous variable (N in modelling group = 52; N in holdout group = 13). This resulted in a final N of 33,802 in the modelling subgroup.

Finally, fit of the model was assessed by the root mean square error of the approximation (RMSEA; values 0.01, 0.05, and 0.08 indicate excellent, good, and acceptable fit, respectively), standardized root mean squared residual (SRMR; values <0.08 indicate good fit), and comparative fit index (CFI; values >0.90 indicate good fit).

#### Constrained confirmatory factor analysis

To evaluate the applicability of the factor model beyond the training subgroup, we obtained fit metrics in the validation holdout subgroup (N=8,465; N=8,452 after removing continuous-variable outliers) while constraining the model parameters (i.e., factor loadings) to those estimated in the training subgroup. Fit was assessed by the root mean square error of the approximation (RMSEA), standardized root mean squared residual (SRMR), and comparative fit index (CFI).

### Differences between EFA and CFA

Our exploratory and confirmatory factor analyses differed in several important ways. First, the exploratory factor analysis was performed on a partial pairwise Pearson correlation matrix, whereas the confirmatory factor analysis was performed on an imputed version of the core data set modeling correlations as appropriate (e.g., polychoric for pairs of ordered variables, Pearson for pairs of continuous variables, and polyserial for pairs containing one of each). Second, in an EFA, all paths and cross loadings are modeled;

however, in a CFA, paths are pre-specified in accordance with a hypothesized structure (e.g., in our case, factors were said to include all items with a loading of >0.1 in the EFA). These changes in modeling resulted in dropping 59 items and one factor when moving from the EFA to CFA (see **Table S6** for reasons for dropping each item) in order to facilitate model fitting.

Factor 8, the factor that we dropped from analysis, previously contained items relating to air and noise pollution, as well as road traffic. As we began investigating Heywood cases in the initial model within the CFA framework, it became evident that Factor 8 was primarily serving to separate the covariance due to the traffic items from the rest of the items contained in Factor 4, which encompasses items related to population density, pollution, and transportation. Once items within the traffic cluster were removed in the CFA due to collinearity, the remaining items were almost entirely contained within Factor 4. We therefore removed Factor 8 due to redundancy.

Apart from removing Factor 8, structure and interpretation within each factor was generally consistent. Of the remaining 35 factors, correlation between loadings for the EFA vs. CFA were very high (i.e., r>0.9) for 9 factors (i.e., Factors 1, 6, 17, 21, 22, 23, 25, 26, and 36), high (i.e., r=0.7-0.9) for 15 factors (i.e., Factors 2, 3, 4, 5, 7, 9, 11, 15, 16, 19, 28, 27, 29, 31, and 32), moderate (i.e., r=0.5-0.7) for 6 factors (i.e., Factors 10, 12, 18, 20, 33, and 34), low (i.e., r=0.3-0.5) for 2 factors (i.e., Factors 13 and 24), and very low (i.e., r<0.3) for 3 factors (i.e., Factors 14, 30, and 35).

Factors with low and very low loading correlations were primarily affected by removal of top items or highly correlated clusters of items. For example, the top-loading items in

Factor 14 in the EFA were a cluster relating to disability assistance (i.e., “Attendance/disability/mobility allowance: None of the above”, “Attendance/disability/mobility allowance: Disability living allowance”, “Attendance/disability/mobility allowance: Blue badge”, and “Current employment status: Unable to work because of sickness or disability”) that were removed due to multicollinearity in the CFA. The resulting factor relates to long-term disability, but loadings within the factor we reorganized such that items relating to pain medication and joint and bone disease were much more prominent. Similarly, a cluster of items within Factor 30 relating to diet (e.g., “Pork intake”, “Beef intake”, and “Never eat eggs, dairy, wheat, sugar: I eat all of the above”) was removed in the CFA due to low cell counts and multicollinearity issues. As a result, a factor that was previously related to poorer dietary habits became related a mix of poor dietary habits (e.g., “Bread type: White” and decreased “Fresh fruit intake”), blood inflammation/infection markers (e.g., “White blood cell (leukocyte) count” and “Platelet count”), and poor health behaviors (e.g., decreased “Leisure/social activities: Sports club or gym” and “Water intake”).

Our multi-stage approach, which was selected to help minimize computational burden and appropriately handle missingness, made it infeasible for us to return to the EFA and generate a new model of the data. Instead, we chose to move forward with the model suggested by the CFA, in spite of these changes to certain specific factors, due to overall acceptable fit to the data.

### Factor score generation

Once the factor model was determined, we then computed factor scores for each latent factor for each individual in the full EUR sample, taking into account differential missingness patterns across individuals.

Given the factor analysis model

$$X = F\Lambda' + \epsilon$$

and the resulting fitted parameter estimates from the CFA, it is possible to estimate each person's latent factor scores as a linear combination of the observed items. In other words, we can define a matrix of factor scoring coefficients $A$ such that

$$\widehat{F} = XA$$

And thus for each individual $i$ the estimated factor score for factor $t$ is a weighted sum of the items

$$\hat{f}_{i,t} = \sum_{j} a_{j,t} x_{i,j}$$

If we take the factor model as true then the resulting estimates are of an individual's "true" score for the underlying latent construct, otherwise they simply estimate the value that best approximates the observed data for each individual with the low rank approximation of the complete data modelled by the CFA. These estimated factor scores can then be used in subsequent analyses of how the factor score are related to other variables outside the CFA (e.g., genetics, mortality, other diagnoses).

$$\widehat{F_B} = XA_B$$

The current analysis uses two sets of factor scores, corresponding to two different estimation methods to compute the factor scoring coefficients $A$: (1) Bartlett's method (11,12), used when the estimated factor score is the dependent variable in an analysis (e.g. in the GWAS), and (2) the Thomson-Thurstone (Regression) method (13,14), used when the estimated factor score is the independent variable in an analysis (e.g., in the phenotypic associations).

This use of two different estimated factor scores, where the choice of factor score estimate to use in a given analysis depends on its placement in the model, follows previous recommendations to avoid biased results in factor score regression (15,16). Briefly, because these factor score estimation methods differ in how they prioritize correlation and covariance of the estimate with the "true" factor score they have different expected bias in the results of regressions that include the estimated factor score. Using factor scores estimated by Bartlett's method as dependent variables and factor scores estimated by the Thurstone-Thomson method as independent variables avoids this bias in both cases. In addition, both estimators have been extremely well studied in the factor analysis literature, and thus provide a familiar foundation for interpreting the factor scores in the current analysis. Below we introduce both the Bartlett and Thomson-Thurstone estimators for factor scores, followed by the modifications we make to each method to account for the presence of missing data and categorical variables in the current analysis.

#### Bartlett estimator

Bartlett's estimator (17) for individual factor scores is given by

$$\widehat{F_B} = XA_B$$

$$A_B = \Psi^{-1}\Lambda(\Lambda'\Psi^{-1}\Lambda)^{-1}$$

Where: (1) $X$ is the $n \times p$ vector of $p$ observed variables in the factor model for $n$ individuals, standardized and residualized for exogenous covariates, as in the fitted CFA; (2) $A_B$ is the $p \times t$ matrix of coefficients to estimate the $t$ factors from $p$ items (i.e., the factor scoring matrix); (3) $\Lambda$ is the $p \times t$ matrix of factor loadings from the fitted CFA; and (4) $\Psi$ is the $p \times p$ diagonal matrix of residual variances from the fitted CFA (i.e., item uniquenesses).

Bartlett's estimator can be motivated as a weighted least squares estimate that minimizes the residual variance of the items in the factor model after weighting for the expected variance due to the fitted item uniquenesses from the model. Specifically, recalling the factor model

$$X = F\Lambda' + \epsilon$$

(assuming all $X$ have been centered), the Bartlett estimator minimizes

$$\sum_j \frac{\epsilon_{ij}^2}{\Psi_j} = (X - F\Lambda')'\Psi^{-1}(X - F\Lambda')$$

For $F$, which is consistent with weighted least squares with weights $\Psi^{-1}$. Using the estimated values from the CFA and following standard weighted least squares this yields

$$\hat{F} = X\hat{\Psi}^{-1}\hat{\Lambda}(\hat{\Lambda}'\hat{\Psi}^{-1}\hat{\Lambda})^{-1} = XA$$

One key property of Bartlett estimator is that it produces unbiased estimates of the effect of variables on the (unobserved) true factor score when used as the dependent variable in factor score regression (as long as the model is correctly specified (16)). Letting $Z$ be a $n \times k$ matrix of of observed data on $k$ variables, regressing $\hat{F}$ on these variables by ordinary least squares will estimate regression coefficients

$$\hat{\beta} = (Z'Z)^{-1}Z'\hat{F}$$

$$= (Z'Z)^{-1}Z'XA$$

$$= (Z'Z)^{-1}Z'(F\Lambda' + \epsilon)\,\hat{\Psi}^{-1}\hat{\Lambda}(\hat{\Lambda}'\hat{\Psi}^{-1}\hat{\Lambda})^{-1}$$

$$= (Z'Z)^{-1}Z'F\Lambda'\hat{\Psi}^{-1}\hat{\Lambda}\left(\hat{\Lambda}'^{\hat{\Psi}^{-1}}\hat{\Lambda}\right)^{-1} + (Z'Z)^{-1}Z'\epsilon\hat{\Psi}^{-1}\hat{\Lambda}(\hat{\Lambda}'\hat{\Psi}^{-1}\hat{\Lambda})^{-1}$$

As $\hat{\Lambda}$ converges to $\Lambda$, as expected for a consistent estimator of $\Lambda$ in CFA, this reduces to

$$\hat{\beta} \approx (Z'Z)^{-1}Z'F + (Z'Z)^{-1}Z'\epsilon\hat{\Psi}^{-1}\Lambda(\Lambda'\hat{\Psi}^{-1}\Lambda)^{-1}$$

Then as long as the $k$ regression values are independent of the unique item residuals $\epsilon$, either directly ($E[Z'\epsilon] = 0$ or as a weighted average across the factor ($E[Z'\epsilon\Psi^{-1}A] = 0$, this leaves

$$\hat{\beta} \approx (Z'Z)^{-1}Z'F$$

i.e., the effect that would be estimated from regressing the “true” score $F$ on $Z$. It is for this reason that we use the Bartlett estimator for factor scores that are the dependent variable in an analysis. We discuss additional implications of this expectation as it relates to our decision to apply a threshold for minimum correlation with complete data factor scores below.

#### Thomson-Thurstone estimator

The Thomson-Thurstone estimator (13,14) for individual factor scores is given by

$$\widehat{F_{TT}} = XA_{TT}$$

$$A_{TT} = \Sigma^{-1}\Lambda = \Psi^{-1}\Lambda(I + \Lambda'\Psi^{-1}\Lambda)^{-1}$$

Where: (1) $X$ is the $n \times p$ vector of $p$ observed variables in the factor model for $n$ individuals, standardized and residualized for exogenous covariates, as in the fitted CFA; (2) $A$ is the $p \times t$ matrix of coefficients to estimate the $t$ factors from $p$ items (i.e., the factor scoring matrix); (3) $\Sigma^{-1}$ is the $p \times p$ covariance matrix for the observed items; (4) $\Lambda$ is the $p \times t$ matrix of factor loadings from the fitted CFA; (5) $\Psi$ is the $p \times p$ diagonal matrix of residual variances from the fitted CFA (i.e., item uniquenesses); and (6) $I$ is a $t$-dimensional identity matrix. The latter formulation of $A$ follows from a special case of the matrix inversion lemma (18–20) under the CFA model (i.e., $\Sigma = \Lambda\Lambda' + \Psi$) given the factors are uncorrelated with unit variance. We implement the Thomson-Thurstone estimator using this latter formulation.

The Thomson-Thurstone estimator, also sometimes known as the regression method for factor score estimation, is motivated by regression of the "true" factor scores $F$ on the items $X$. This can't be done directly since factor scores are unobserved, but by noting that the loadings $\Lambda$ are the expected covariance between $F$ and $X$ under the model it then follows that the desired regression estimate is given by

$$\hat{F} = X([X'X]^{-1}X'F) = X\Sigma^{-1}\Lambda = XA_{TT}$$

It can be shown (21) that the resulting factor score estimates are the best linear prediction, minimizing the trace and determinant of the expected mean squared error (MSE) matrix $E[(\hat{F}_{TT} - F)(\hat{F}_{TT} - F)']$.

Factor score regression with the Thomson-Thurstone estimator yields unbiased estimates only when the factor score is used as the independent variable in regression with an observed dependent variable (16). This is the converse of the Bartlett estimator, which provides unbiased factor score regression results when used as a dependent variable. We therefore use the bias-avoiding method of factor score regression (15,16), using the Bartlett estimator for factor score estimates used as dependent variables and the Thomson-Thurstone estimator for factor score estimates used as independent variables. To preserve the proper scaling of the factor scores with these properties we do not standardize the factor score estimates.

#### Modifications for categorical and missing data

The above framework for the factor score estimators assumes that all of the item data $X$ is observed. Although this is true for observed continuous items in the model, it is not true for the CFA model where either (a) the observed data is categorical and modelled through a link function or (b) the data is unobserved due to missingness. Ideally, these features could be addressed instead by estimating factor scores with maximum likelihood, but that is not currently computationally feasible. We describe here our solutions for addressing the problem of categorical and missing data, followed by a comparison of the performance of our approach to maximum likelihood estimation with imputed data.

##### Measurement scale for categorical variables

For categorical items, the CFA model assumes a (ordered) probit link function to connect the observed categorical values to a normally distributed continuous latent value. The estimated loadings $\widehat{\Lambda}$ and residual variances $\widehat{\Psi}^{-1}$ reflect effects for this continuous latent value. The factor scoring estimators therefore estimates the factor scoring coefficients $A$ that would be appropriate for estimating scores based on the unobserved latent variable. The observed categorical variable, however, will have different scaling (e.g. binary 0/1 rather than continuous scale relative to a standard normal distribution) and will have a weaker relationship with the factor due to the information lost when discretizing the underlying continuous variability (e.g., the same phenomenon often described for the impact of dichotomizing continuous variables and the distinction between observed and liability scale heritability for binary traits).

We first address the measurement scale by estimating the expected value of the latent continuous variable for each person for each categorical variable based on the fitted probit link. Specifically, let $x_j^*$ be the continuous latent variable corresponding to the observed categorical variable $x_j$. Then we estimate the expected value of $x_j^*$ according to a truncated normal distribution with mean equal to the individual's predicted value of the linear model from the probit regression with exogenous covariates $\boldsymbol{z}$, unit variance, and upper and lower truncation thresholds set according to the observed $x_{i,j}$ and the in-sample prevalences of the possible responses $\boldsymbol{p}$. We then use the difference between this $E[x_{i,j}^*|x_{i,j}, \boldsymbol{z}, \boldsymbol{p}]$ and the predicted value from the probit regression as our estimate of the desired latent continuous variable residualized for the exogenous covariates. We substitute this estimated residual in place of the observed $x_{i,j}$ for factor scoring, along with transforming the loadings and residual variances used in each estimator to reflect the attenuated signal in these observed items.

#### Transformed loadings

While we can approximate the latent continuous values for each categorical item, the expected values do not fully recover the variation of the unobserved latent value. To account for the attenuated signal present in these expected residualized values, we adjust the loadings using estimating factor scores to reflect the expected weaker covariance between the factor and the observed categorical variable. Again, we let

$$x_j^* = \sum_t \hat{\lambda}_{j,t} f_t + \sum_k \gamma_k z_k + e_j^*$$

be the model-implied fit with exogenous covariates $z_k$ for the continuous latent variable $x_j^*$ corresponding to the observed categorical variable $x_j$. For the CFA, the terms here are standardized such that $var(f_t) = 1$ for each factor, $var(x_j^* | z_k) = 1$, and each $f_t$ is independent of the covariates and all other factors. Thus each $\hat{\lambda}_{j,t}$ is the estimated partial correlation between the factor and the continuous latent $x_j^*$. For the purposes of factor score estimation, we want to transform this loading to a value that reflects the partial covariance of the factor with the observed residualized categorical item computed above.

To achieve this transformation, we first note that $\hat{\lambda}_{j,t}$ is effectively an estimate of the polyserial partial correlation between the factor and item (or biserial when the categorical variable is binary). Thus by standard expectations for the polyserial correlation (22) we approximate

$$\hat{\lambda}_{j,t} = cor(f_t, x_j^* | z_k)$$

$$\approx cor(f_t, x_j | z_k) \times \frac{\sigma_x}{\sum_{c \epsilon C} h(c)}$$

where $\sigma_x^2$ is the variance of the observed categorical variable $x_j$ and $h(c)$ is the density of the standard normal distribution at the threshold for each category $c$ of the categorical $x_j$ under the assumed probit link function. This expectation would hold more directly if we didn't transform the categorical data to residualize on covariates as described. Still, in most cases the impact of the covariates is small enough that the transformed residualized variables remain approximately categorical and thus should be well approximated by the expectations for a polyserial correlation; we thus adopt this approximation for convenience. The above expression can then be given in terms of the desired partial covariance as

$$\hat{\lambda}_{j,t} \approx \frac{cov(f_t, x_j|z_k)}{\sqrt{var(f_t)var(x_j|z_k)}} \times \frac{\sigma_x}{\sum_{c \epsilon C} h(c)}$$

Rearranging and noting $var(f_t) = 1$ results in

$$cov(f_t, x_j|z_k) \approx \hat{\lambda}_{j,t}\sqrt{var(x_j|z_k)} \times \frac{\sum_{c \epsilon C} h(c)}{\sigma_x}$$

where $var(x_j|z_k)$ and $\sigma_x$ do not cancel due to the difference in conditioning on covariates $z_k$. In practice, we estimate $var(x_j|z_k)$ from the residualized categorical data (described above) and we estimate $\sigma_x$ from the category probabilities implied by the fitted probit model in the CFA. We then use the resulting estimate of $cov(f_t, x_j|z_k)$ in place of the estimated loadings $\hat{\lambda}_{j,t}$ for categorical items for the purpose of estimating factor scoring coefficients $A$. Note however that this transformation is only for factor scoring, and does not affect the loadings reported for the CFA.

#### Transformed residual variances

Consistent with the loadings $\hat{\lambda}_{j,t}$, the residual variances $\widehat{\Psi}^{-1}$ present in both factor score estimator equations are estimated in the CFA assuming a probit link for categorical variables. For the purpose of factor score estimation we similarly transform these values to be consistent with the expected attenuation of signal on the transformed and residualized categorical variables.

Specifically, for the CFA $\widehat{\Psi}^{-1}$ is a diagonal matrix whose elements are the residual variance of each (latent) item conditional on the factors and covariates. Categorical items are standardized such that $var(x_j^*|z_k) = 1$, therefore the residual is equal to 1 minus the variance explained by the factors $R^2_{j^*,F}$, where $j^*$ denotes that the variance explained is for the latent continuous item $x_j^*$.

$$\psi_{jj} = 1 - R^2_{j^*,F}$$

As for the loadings, $R^2_{j^*,F}$ is a squared polyserial correlation. There we can approximate (22)

$$\psi_{jj} \approx 1 - (R^2_{j,F} \times \frac{\sigma_x}{\left[\sum_{c \epsilon C} h(c)\right]^2})$$

The corresponding residual variance of the observed categorical items conditional on the covariates and the factors (i.e., the desired value for factor scoring with the observed data) can be expressed as

$$\psi_{jj,obs} \approx var(x_j|z_k) \times (1 - R^2_{j,F})$$

As before, this approximation is inexact due to the transformation and residualization of the categorical items as described above, but we anticipate this approximation will perform adequately, especially when covariate effects are small. Rearrangement and substitution with the approximation for $\psi_{jj}$ leads to

$$\psi_{jj,obs} \approx var(x_j|z_k) \times (1 - [1 - \psi_{jj}] \frac{[\sum_{c \epsilon C} h(c)]^2}{\sigma_x^2})$$

where $\psi_{jj}$ is the estimated residual variance in the CFA and the remaining terms are calculated as in the transformation of the loadings. As with the loadings, these estimated residual variances for the observed categorical items are then substituted into $\widehat{\Psi}^{-1}$ for the estimation of factor scoring coefficients $A$.

#### Estimating factor scores with missingness

For individuals with missing data some elements of $x_i$ are unobserved. This prevents calculation of the complete factor scores

$$\hat{f}_{i,t} = \sum_j a_{j,t} x_{i,j}$$

Given the high rates of missingness in UK Biobank, there is obvious interest in being able to estimate these factor scores for individuals with missing data. Thankfully the factor score estimators provides a natural way to use the same estimation framework when some items are unobserved.

Recall that the Bartlett estimator derives from WLS of the items in the factor model. Then if $M$ is a set of unobserved items then we could chose to optimize for

$$\sum_{j \notin M} \frac{\epsilon_{ij}^2}{\Psi_j} = (X_{-M} - F\Lambda'_{-M})'\Psi^{-1}{}_{-M}(X_{-M} - F\Lambda'_{-M})$$

where $X_{-M}$, $\Lambda'_{-M}$, and $\Psi_{-M}$ denote the observed data, loadings, and residual variance matrices omitting the rows and columns corresponding to the unobserved items in $M$. This is equivalent to giving zero weight to the amount of residual error in unobserved items. Conceptually, this is the same treatment given to items not present in the CFA that might also reflect the fitted factors. Following the same derivations for the Bartlett estimator, the resulting factor scoring coefficients would be estimated as

$$A_{-M} = \widehat{\Psi}_{-M}^{-1}\widehat{\Lambda}_{-M}(\widehat{\Lambda'}_{-M}\widehat{\Psi}_{-M}^{-1}\widehat{\Lambda}_{-M})^{-1}$$

Thus factor scoring coefficients, and the resulting estimated factor scores $\hat{F} = X_{-M}A_{-M}$, can be calculated for each individual based on their set of available observed items. The only exception is where $\widehat{\Lambda'}_{-M}\widehat{\Psi}_{-M}^{-1}\widehat{\Lambda}_{-M}$ is singular, which will occur when one or more factor have no observed items with non-zero items (i.e., one or more columns of $\widehat{\Lambda}_{-M}$ contains all zeros); in that case we drop the individual from the analysis.

The same argument similarly applies to the Thomson-Thurstone factor scores intended for use as independent variables. Recalling that the Thomson-Thurstone estimator is based on the idea of regressing the "true" factor scores on the available items, subsetting to observed items gives

$$\widehat{F_{-M}} = X_{-M}([X'_{-M}X_{-M}]^{-1}X'_{-M}F) = X_{-M}\hat{\Sigma}_{-\mathrm{M}}^{-1}\widehat{\Lambda}_{-\mathrm{M}} = XA_{TT,-M}$$

Thus as with the Bartlett scores, we can compute Thomson-Thurstone scores for each individual based on the subset of observed items. Individuals missing all items for one or more factors are similarly omitted.

Based on this solution for factor scoring in the presence of missingness, we may have a few concerns. First, in order to maintain constant covariance with other variables the factor scores will be highly heteroskedastic dependent on the missingness pattern. Second, covariance of other variables with the factor scores will not remain constant if those variables are related to item uniquenesses, potentially influencing regression results and their interpretation at different levels of missingness. Third, the fitted CFA may not have measurement invariance across different missingness levels, and thus the CFA may be misspecified for some missingness patterns.

#### Heteroskedasticity of Bartlett factor scores by missingness pattern

Because the set of missing variables $M$ varies between individuals, the estimated Bartlett factor scores used as dependent variables will be heteroskedastic as discussed above. When the factor score is used as a dependent variable, we can potentially account for that heteroskedasticity in downstream analyses by estimating the residual variance in the estimated factor score for a given individual as a function of the missingness pattern.

Specifically, we focus on the use case of GWAS of the estimated factor score as the example primary analysis of interest with the factor score as the dependent variable.

$$\hat{f}_{i,t} = \beta_{0,t} + \beta_{1,j,t} SNP_{i,j} + \sum_k \beta_{k,t} z_{i,k} + e_{i,t}$$

i.e., regression of the factor score on a SNP and accompanying GWAS covariates $z$. Heteroskedasticity will exist here if the residual variance differs between individuals.

$$var(e_{i,t}) = \sigma_{i,t}^2$$

One conventional solution to efficient estimation of regression in the presence of heteroskedasticity is to use weighted least squares (WLS) with weights for each observation proportional to residual variance. Specifically, WLS estimation of this GWAS model will be optimal if the model is correctly specified and we can compute weights equal to the inverse residual variance, $w_{i,t} = {}^{1}/_{var(e_{i,t})}$.

With this approach it becomes critically valuable that the Bartlett estimator provides unbiased estimates and maintains the covariance of the estimate with the "true" factor score. This ensures that for regression analyses using the factor score as a dependent variable the population coefficients $\beta$ will remain constant in expectation across groups of individuals regardless of their missingness pattern in estimating $\hat{f}_{i,t}$, as long as the SNP is associated with the factor and not the uniquenesses of the items in the individual factor.

To estimate the expected residual variance, we first note that we expect most SNPs to have small or null effects. Similarly, since the factor scores are constructed from items that have already been residualized on the standard GWAS covariates, we expect little to no covariate effect. Thus we can approximate

$$var(e_{i,t}) = var(\hat{f}_{i,t}|SNP_i, z_i) \approx var(\hat{f}_{i,t})$$

For an individual missing observations for a set of items $M$, the expected variance of the estimated factor score is

$$var(\hat{f}_{i,t}) = var(X_{-M}A_{-M,t}) = E[A'_{-M,t}X'_{-M}X_{-M}A_{-M,t}]$$

With observed data $X_{-M}$ (with transformation of categorical variables as described previously) and factor scoring matrix $A_{-M}$ computed from the Bartlett estimator for the corresponding missingness pattern. Treating the factor scoring coefficients as fixed,

$$var(\hat{f}_{i,t}) = A'_{-M,t}E[X'_{-M}X_{-M}]A_{-M,t} = A'_{-M,t}S_{-M}A_{-M,t}$$

where $S_{-M}$ is the covariance matrix of the observed items. Assuming that the covariance of the items is constant across the missingness patterns, we can compute $S_{-M}$ as the sample covariance matrix from the pairwise complete observations in $X$ (again, after residualization). Therefore we estimate the weights for WLS as

$$w_{i,t} = \frac{1}{A'_{-M,t}S_{-M}A_{-M,t}}$$

for each individual $i$ with missingness pattern $M$.

If these WLS weights are correct then the resulting regression will provide the best linear unbiased estimator of the SNP effects in the GWAS. To the extent that these $w_{i,t}$ are incorrect due to estimation error, model misspecification, or other issues, this WLS regression will behave like OLS in the presence of heteroskedasticity: coefficient estimates will remain asymptotically unbiased, but will not be efficiently estimated and their standard errors will tend to be underestimated, increasing type I error rates.

#### Missingness-based heteroskedasticity and bias for Thomson-Thurstone scores

The variation introduced by differences in missingness patterns is harder to resolve for the independent variable factor scores from the Thomson-Thurstone estimator. First, in order to use WLS as a control for differential information due to missingness when the factor score is an independent variable rather than the dependent variable would require estimation of $var(y_i|\hat{f}_{i,t}, z_i)$, the residual variance of the observed phenotype of interest $y$ conditional on the factor score and covariates. This quantity depends on the true effect of the factor score on $y$, and unlike the GWAS case it is unlikely that we can assume that the effect of the factor – as well as the effects of the covariates – are small enough they could be ignored in estimating appropriate WLS weights.

Given the difficulty of estimating appropriate weights for WLS in the independent variable case, we instead address the expected heteroskedasticity from differential missingness in the items used to estimate the Thomson-Thurstone scores using sandwich (i.e., Huber-White (23)) standard errors. Briefly, for linear regression in the presence of heteroskedasticity

$$(X'X)^{-1}X'\widehat{\Omega}X(X'X)^{-1} \ ,$$

where $\widehat{\Omega}$ is a diagonal matrix of the estimated squared OLS residuals

$$\widehat{\Omega} = diag([y_i - \widehat{y}_i]^2) \ ,$$

is a consistent estimator of the covariance matrix for sampling error in the regression coefficients. Although this approach does not affect the estimation of the regression coefficients themselves, it does help improve the control of Type I error rates. Additional refinements of the sandwich estimator have been proposed (24), but we opt for the original estimator of White (23) due to its computational efficiency and the minimal impact of further adjustments at large sample sizes (25) like our current analyses in UK Biobank. We use these standard errors for the linear regression analyses (i.e., biomarkers and phecodes). On the other hand, although robust standard errors have been proposed for Cox regression (26–28), they often require auxiliary data or specifying additional likelihoods with modelling assumptions for the error distribution. Therefore for ease of implementation we choose not to apply a heteroskedasticity correction for the current mortality analyses (though see **Minimum correlation with complete data scores** below which limits the potential differential missingness in the mortality analysis).

In addition to incorrect standard errors, there is also a risk of biased parameter estimates in regression with the independent variable factor scores from the Thomson-Thurstone estimator. As previously noted (16), The Thomson-Thurstone estimator yields factor scores that maintain

$$E\left(y_i \middle| \hat{f}_{i,t,-M}, z_i\right) = \beta_0 + \beta_{1,t}\hat{f}_{i,t,-M} + \sum_k \beta_{k,t} z_{i,k}$$

with the same regression coefficients $\boldsymbol{\beta}$ for any given missingness pattern $M$ in estimation of the factor score. The OLS regression estimator

$$\widehat{\boldsymbol{\beta}} = (X'X)^{-1}X'y$$

however, will not reliably estimate this $\beta_{1,t}$ since under the assumption of homoskedasticity it evaluates the variance of the factor score – as well as its covariance with other covariates – across all individuals in $(X'X)^{-1}$ rather than conditional on missingness. Therefore estimates of $\beta_{1,t}$ will be biased if the factor score has heteroskedasticity and is correlated with other covariates in the model. While corrections for the general case of this "errors in variables" bias have been proposed (29,30), they generally rely on additional instrumental variables or estimating the covariance matrix of estimation error across the covariates, which are not easily available for our factor scores. For our current use cases, we expect the correlation between the factor score and the regression covariates to be generally small, given the factor scores are already estimated conditional on the exongenous covariates in the CFA model and we do not expect substantial correlation of the factor scores with the added covariates for assessment center, dilution factor for biomarkers, or date of baseline assessment for mortality analyses (indeed differences in the distribution of factor scores by e.g. assessment center would likely imply more general violations of factor invariance assumptions, affecting more than just the phenotypic factor score regressions, see **Notes on modeling and limitations**). This suggests that the bias from using OLS with the Thomson-Thurstone estimator of factor scores as independent variables should be somewhat limited. We therefore do not attempt further adjustments for bias in the independent factor score analyses.

#### Minimum correlation with complete data scores

Although we can estimate factor scores allowing for missingness, our confidence in those factor scores decreases as the level of missingness increases. For the Bartlett estimator, increased missingness means the factor score is estimated from fewer items, reducing the plausibility of assuming that the predictors of interest (e.g. GWAS SNPs) will be uncorrelated with the sampling variation in the factor score estimate (e.g. that $E[Z'\epsilon] = 0$). For the Thomson-Thurstone estimator, increasing variability in the accuracy of the factor score between individuals due to differential missingness will also exacerbate the regression attenuation caused by the error in the independent variables. For this reason, we choose to limit the impact of missingness on our analyses of the factor scores by restricting analysis to individuals with sufficiently informative factor scores.

Specifically, for each individual we evaluate the expected correlation between the factor score computed with their observed items and their corresponding estimated factor score if all items had been observed (i.e. complete data no missingness).

$$E\left[cor\left(\hat{f}_{i,t}, \hat{f}_{i,t,complete}\right)\right] = E\left[\frac{A'_{-M,t}X'_{-M}XA_t}{\sqrt{A'_{-M,t}X'_{-M}X_{-M}A_{-M,t} \times A'_tX'XA_t}}\right]$$

$$= \frac{A'_{-M,t}S_{-M,\cdot}A_t}{\sqrt{A'_{-M,t}S_{-M,-M}A_{-M,t} \times A'_tSA_t}}$$

where $A_t$ and $A'_{-M,t}$ are the vectors of factor scoring coefficients for factor $t$ with all items and with items observed in missingness pattern $M$, respectively, and $S$ is the sample covariance matrix for the residualized items, with subscripts denoting subsetting for missingness $M$ on the rows or columns, respectively. Note this expectation assumes that the distribution of the items is independent of the missingness pattern (e.g. that the expectation of $X'_{-M}X$ remains the sample covariance $S$ regardless of $M$), the same assumption made for WLS weights for the Bartlett estimator. This assumption is likely violated in practice, but we only rely on this assumption here to derive this approximate metric to use for filtering individuals for inclusion in analysis, and do not make any inference on the estimate of $E\left[cor\left(\hat{f}_{i,t}, \hat{f}_{i,t,complete}\right)\right]$.

Based on this metric, we exclude individuals with $E\left[cor\left(\hat{f}_{i,t}, \hat{f}_{i,t,complete}\right)\right]^2 < 0.8$ from analyses with factor scores for factor $t$. This exclusion was computed and applied for both independent- and dependent-variable use of the factor scores. We chose this threshold based on examination of the distribution of $E\left[cor\left(\hat{f}_{i,t}, \hat{f}_{i,t,complete}\right)\right]$ across individuals for each factor (**Figure S10**), with consideration of how this distribution corresponded to structured missingness for top items and the potential impact of that missingness on item interpretability. We observed this threshold to be more universally liberal in the independent than dependent factor scores, thus we also chose to restrict the sample in independent factor score analyses to only those individuals included in the genetic analyses to allow for better concordance and comparability in samples across phenotypic and genetic analyses.

#### Validation of factor scoring methods in the core data group

To validate our factor-score-generating methodologies, we compared scores from our methods to those generated using a maximum-likelihood-based (ML) method in lavaan for the core data group. Factor scoring was performed using the ML option in lavaan for all 10 multiple imputations of the core dataset. Though we chose to compare our scoring methods to the ML method in lavaan, the latter cannot be considered the “gold standard,” as numerous factor-scoring methods exist, with each simply relying on and/or prioritizing different assumptions. However, this comparison performed to provide more confidence in our chosen method, which sought to mimic ML estimation in spirit.

To test for phenotypic concordance across methods, we obtained Pearson correlation coefficients between factor scores generated using our method and the mean of those obtained in lavaan across all 10 imputations. These correlation estimates did not weight individuals based on the expected precision of their factor scores, but were restricted to only individuals meeting our analytic inclusion thresholds. Phenotypic correlations were moderate to very high for the dependent-variable (range: 0.531-0.994, mean=0.842(0.137)) and independent-variable (range: 0.540-0.993, mean=0.843[0.132]) formulations. Concordance across dependent- and independent-variable formulations was excellent (range: 0.967-1.000, mean=0.992[0.009]).

We performed GWAS for each factor for the scores generated with lavaan and with our dependent-variable factor scores. GWAS of our scores were conducted using weighted least squares (WLS) regression and appropriate covariates as described in **Methods**. GWAS of the lavaan-generated scores were similarly conducted with WLS and estimated inverse-variance weights $w_{i,t} = 1/var(\hat{f}_{i,t})$. To estimate the variance of lavaan-generated factor scores, we note that under the law of total variance we can decompose

$$Var(\hat{f}_{i,t}) = E[Var(\hat{f}_{i,t}|M_i)] + Var[E(\hat{f}_{i,t}|M_i)]$$

with the individuals missingness $M_i$. We then estimate

$$E[Var(\hat{f}_{i,t}|M_i)] = Var(\hat{f}^{(r)}{}_{i,t})/R$$

as the standard error of the mean across the $R = 10$ imputation replicates $\hat{f}^{(r)}{}_{i,t}$ for individual $i$, and approximate $Var[E(\hat{f}_{i,t}|M_i)]$ by the observed variance of the lavaan-generated factor scores across individuals. These weights thus serve as a proxy for imputation quality and missingness proportion across participants analogous to the WLS weights used for the dependent variable factor scores.

For each factor the GWAS results from the two methods were compared using LDSC. Heritability estimates from S-LDSC for each factor were generally concordant across methods (range absolute difference: 0.0002-0.079, mean=0.015(0.019)), with no evidence of systematic bias. The genetic correlation between methods for each factor was moderate to very high (range: 0.577-1.072, mean=0.976(0.090); **Fig S14**).

Lastly, we also compare our use of the Bartlett estimator to the generalized least squares (GLS) approach proposed by Bentler and Yuan (31). Similar to the WLS approach of the Bartlett estimator, the GLS approach weights using the full covariance matrix of the items $\Sigma$ rather than the estimated residual variances $\hat{\Psi}$.

$$A_{GLS} = \Sigma^{-1}\Lambda(\Lambda'\Sigma^{-1}\Lambda)^{-1}$$

We find that the GLS estimator using the sample covariance matrix for $\Sigma$ yields highly similar results to our modified Bartlett estimator despite our modifications to $\hat{\Psi}$ for the Bartlett estimator (results not shown). Along with the comparison to ML estimation, this provides additional reassuring evidence that our chosen factor score estimators are at least somewhat robust to our choice of weighting modifications.

### Notes on modeling and limitations

In interpreting the results of this paper, it is important to keep in mind that these factors are not “real” and do not exist as distinct, measurable entities. Instead, they are simply statistical tools that we use to understand relationships between observed variables in this particular cohort and to facilitate downstream analyses by modelling items' covariance structure with a reduced rank. No prior factor analysis, to our knowledge, has modeled such a large array of variables, across multiple assessments, and spanning multiple data types. Given the extensive adaptations made to traditional factor analysis methodology to use FA as a principled dimensionality reduction technique, we wish to outline "lessons learned" during the course of the analysis, to inform future studies and also highlight potential limitations. In the following sections, we touch on a host of analytic decisions and assumptions made throughout this project and provide concrete examples of the ways in which they may impact results and limit generalizability.

#### Impact of input items on factor structure

Factor analysis, as a psychometric technique, is typically used to model inter-item structure within a single questionnaire (14,32–34). Expanding this technique to cover many different questionnaires and assessments types necessitates that the differential *number* of items covering a particular construct will influence the outcome of the factor model. For example, roughly a quarter of all factors (e.g., Factors 5, 13, 17, 19, 24, 26, 27, 32, and 36) contained items derived primarily from a single questionnaire or assessment. Moreover, the top 3 factors identified by the EFA all contained items broadly related to anxiety and depression. This is unsurprising, since of 730 items used in the EFA, 106 were from the mental health questionnaires of either the initial touchscreen (40 items) or online follow-up (66 items). Results would likely look different if other questionnaires and assessments were included (see **Tables S2** and **S5** for the distribution of items across categories in the final model and core data group, respectively). One questionnaire which was almost entirely excluded from modeling due to low N, Diet by 24-hour recall, contained 317 individual items relating to intake of certain foods over a 24-hour period. If this assessment had been included, we may have observed one or more factors more directly reflecting diet. This is also a hazard of trying to generalize our factor solution beyond the UK Biobank: beyond potential differences in sample makeup, the makeup of the items themselves would greatly influence resultant factor structure.

#### Impact of questionnaire structure

Surveys within the UK Biobank contained numerous questions for which a person may select either multiple responses (“categorical-multiple”) or only a single response (“categorical-single”). For example, a person may be asked to select as many vascular disorders as they have been previously diagnosed with, or to select only the type of milk that they most commonly consume. Typically these questions also include an option for “None of the above”. This is a common practice in surveys, and many in the lay public are familiar with such response patterns.

These types of questions pose an analytic problem for methods that rely on correlations across items, such as factor analysis, due to the constraints imposed by the survey question itself. For example, for both categorical-multiple and categorical-single question types, an answer of “None of the above” will necessarily be anticorrelated with dummy variables created for the other responses, resulting in a correlation structure that is entirely dependent upon what other options were included in the question. We observe this induced correlation structure in the results of our initial EFA, where items indicating a response of “None of the above” to a question were consistently among the top-loading items for factors. Factor 13, for example began with two “None of the above” responses to categorical-multiple questions involving use of supplements (e.g., “Vitamin and mineral supplements: None of the above” and “Mineral and other dietary supplements: None of the above”). The Factor consisted mostly of other responses from those two questions. When moving into an EFA framework, the “None of the above” items almost universally caused issues with model fit in the form of Heywood cases, and we therefore removed all of them (see **Differences between EFA and CFA** and **Table S6**).

Categorical-single questions similarly force an anticorrelation across all possible responses. Returning to the example of milk type discussed above, a participant who most commonly drinks skimmed milk will necessarily not most commonly drink full cream or soy milk. Notably it’s quite possible that for example favoring skimmed milk is similar to favoring semi-skimmed milk, in terms of underlying factors affecting milk preference or their correlation with would have traits (both diet and non-diet). Nevertheless the forced choice structure of the categorical-single question will prevent ever observing a positive correlation between skimmed and semi-skimmed milk preference.

Aside from questionnaire structure, this issue also arises from exclusionary diagnoses and medications. For example, individuals are commonly prescribed just one medication from a single class of medications. In these cases, though a group of individuals may have the same disease and associated conditions, each individual could be taking different medications. Therefore, though the disease and its associated conditions would independently be correlated with each of the medications, the medications themselves would be anticorrelated. Within our final CFA model, categorical-single items and medication codes within the same general group (e.g., the ACE inhibitors ramipril, perindopril, and lisinopril) consistently demonstrated the poorest pairwise residual misfit. Attempts to model the residual covariance between these items resulted in problems with the inversion of the information matrix (i.e., a nonpositive definite correlation matrix that then prevents inversion of information matrix) and would have presented an additional computational challenge in the calculation of latent factor scores for individuals. Alternatively, composite items could be created to combine these reported items that are believed to be interchangeable into a single measure for e.g. taking an ACE inhibitor, but building such an item would instead require imposing assumptions on the relationship between the component items and how they each relate to other items in the factor analysis. Given the relatively low prevalence of such items in the overall factor model, we simply included these items "as is" in the analyses.

#### Extreme outliers on continuous variables

Though the primary Neale Lab UKB mega-GWAS results for continuous variables were reported for inverse-rank normal transformations, we used raw data values to facilitate easier comparison between the core and full EUR data groups. However, when moving from the EFA to the CFA framework, we noticed that the top modification indices for the model, which are meant to suggest alterations which would improve model fit, were dominated by continuous items with extreme outliers. This suggests that these extreme outliers (i.e., >20 standard deviations from the mean) were at least in part driving some of the correlation structure. To reduce this influence, we subsequently dropped from the core data group any individuals with a value >20 standard deviations from the mean on any continuous variable (N=65). Depending on whether these outliers reflect true values or data errors, this choice to stabilize the model risks reducing generalizability to individuals with truly extreme values. On the other hand it is unlikely that the phenotypic structure for such individuals would be well modeled regardless of their inclusion. We also note that these outliers still influence our current results, since excluding these outliers for CFA does not eliminate their impact on the structure selected from the EFA, and we chose not to rerun the EFA excluding these 65 individuals.

#### Modeling of non-continuous items

The items in the UK Biobank were derived from many different survey and assessment types, from self-report to verbal interview to medical diagnosis to biometric measurement. UKB therefore contained many different variable types, including continuous, binary, and ordinal, which is expected for any sort of large-scale deep-phenotyping biobank. Within the EFA, to arrive at the initial factor structure, we chose to treat all variables as continuous, consistent with the use of linear regression for GWAS in the Neale Lab UKB Round 2 mega-GWAS. However, treatment of ordered variables as continuous will necessarily misestimate their correlation. In the CFA, we therefore treated all variables as the appropriate data type: 88 as continuous, 346 as binary, and 130 as ordinal, and we considered the ordered variables as thresholds of a continuous liability distribution. Such a conversion seemed appropriate for use in these cases, as diagnoses are often conceptualized as artificial symptom thresholds imposed upon an underlying liability distribution.

#### Forced orthogonality between factors

For modeling purposes, with an eye towards computational scale and downstream analyses, we forced all factors to be orthogonal in both the EFA and CFA. Notably, though the *latent* factors were specified as orthogonal, the *observed* factor scores (generated in our case using extensions of the Bartlett and Thomson-Thurstone methods) were not necessarily orthogonal. Nonetheless, the highest pairwise correlation between factor scores was 0.176 (i.e., between F4 and F33; mean correlation=0.001[0.044]; see **Fig S13**).

Orthogonality likely does not reflect the “true” behavior of latent constructs, and oblique rotations are generally favored with factor analysis. Our choice of such an orthogonal rotation therefore has some important implications for interpretation of our factors. Specifically, each factor must be viewed as representing the covariance structure of the items within it, *once accounting for covariance modeled by the other factors*.

A useful illustration can be found within the four factors most directly related to cardiometabolic disease, Factors 7, 12, 16, and 28. These factors can be broadly characterized as containing items related to BMI and adiposity (Factor 7), hypertension (Factor 12), coronary artery disease (Factor 16), and diabetes (Factor 28). In the “real world,” the variance captured by each of these factors would likely be related; however, within our orthogonal model, we have explicitly required them to be unrelated. As such, Factor 28 could most accurately be interpreted as representing the remaining covariance of the items within it (e.g., mostly reflective of a diabetes diagnosis) once accounting for the variance explained by the other related (and unrelated) factors. The impacts of this orthogonalization can be demonstrated by comparing the GWAS of Factor 28 to a prior GWAS of type 2 diabetes ($r_g$=0.68[0.02]; (35)): the factor has higher genetic overlap with cholesterol measures (e.g., total cholesterol $r_g$=0.29 vs. 0.04; (36)) but lower overlap with BMI ($r_g$=0.23 vs. 0.49; (37)) and an inverse correlation with blood pressure ($r_g$=-0.18 vs. 0.20; (38)), reflective of its inclusion of high cholesterol in its factor definition and its independence of Factors 7 and 12 described above (**Fig S15**). It is therefore critical to not assign meaning to the latent constructs identified by these analyses, or to their underlying genetic etiology, beyond seeing them as useful tools for interrogating potentially relevant axes of phenotypic variation across individuals.

#### Selection of "nuisance" covariates

Within both the EFA and CFA, we chose to residualize observed variables for all covariates used in the Neale Lab UKB Round 2 mega-GWAS: the first 20 genetic PCs, age, chromosomal sex, age$^2$, age-x-chromosomal sex, and age$^2$-x-chromosomal sex. We purposely selected these "nuisance" covariates to be consistent with the prior work of our collaborators, and also to avoid the identification of factors driven by these covariates. Put another way, we wanted to identify consistent axes of variation across the entire EUR subset of the UKB, regardless of chromosomal sex, age, or [EUR] ancestry. One could argue for the inclusion of additional "nuisance" regressors, such as assessment center or measures of socioeconomic status, depending on the intentions and goals of the analysis. In our own analysis, at least one factor, Factor 33, seems to recapitulate some aspects of regional clustering via assessment center. Items within that factor include current home and place-of-birth geographic coordinates, variables related to accommodation and heating types, and certain likely cultural food choices, such as preference for ground vs.

instant coffee and weekly intake of both champagne and white wine. Scores on this factor were generally associated with distance from London, and were highest among those at assessment centers in the immediate area.

#### Assumptions of factor invariance

Factor analysis relies on the assumption that the measurement model, or the relationship between the observed and latent variables, is equivalent across subgroups. These subgroups may be split by demographics (e.g., gender or assessment center) or, in the special case of our analyses, patterns of response. This assumption is unlikely to hold across all subgroups, and a number of methods exist to test for such differences (39). The presence of a reasonably assessment-center-specific factor, described in the previous section, as well as differences in demographics between the core and full EUR groups (see **Characteristics of final core data group**) provide further evidence that this assumption is violated in practice. Additionally, our calculation of factor scores in the presence of missingness explicitly assumes that the covariance structure is the same across individuals regardless of which observed items were actually measured. Caution must thus be exercised in generalizing the model across subgroups. Conversely, a particularly fruitful avenue of future research may be to explicitly model and test for differences across subgroups.

#### Impact of structured missingness

A major goal of this project was to fairly comprehensively model the phenotypic landscape captured by UKB; as such, we sought to include as many assessments as possible to uncover relationships between variables not grouped a priori. However, this decision introduced the issue of structured missingness across assessments, as not all individuals were given or responded to every assessment. To address this, we utilized a multi-stage approach in which the factor model was constructed based on a subsample of individuals with high assessment-level completeness, and then we scored the full EUR sample based on the parameters estimated within that subgroup. As mentioned in the prior section, this approach relies on the assumption of measurement invariance across the core and full subgroups.

Beyond special considerations given to the factor model in the context of structured missingness, problems arose when attempting to estimate factors scores in individuals for which the majority of variance in the latent score was missing. Though our factor scoring method is able to estimate scores in the context of item-level missingness, relying on expected patterns of covariance between the missing and nonmissing items, it cannot "recover" variance when the majority of items within a factor are missing, as in the case of assessment-level missingness. For example, Factor 24 is based primarily on items from an empirical eye assessment which was introduced later in the UKB battery (i.e., 14 of 17 factor items are from that one assessment). As such, individuals who are missing that assessment do not have enough measured indicators to reasonably estimate a value for that latent factor. To account for this, calculated for each missingness pattern the amount of variance explained by the items available versus a hypothetical Bartlett factor score with no missingness. For inclusion in further analyses, we required that individuals have nonmisssing items with the ability to explain at least 80% of variance in that hypothetical factor score. This thresholding severely reduced our sample sizes for a number of factors (e.g., N=75,226 for Factor 24; **Fig S10**) but was necessary to ensure some reasonable degree of comparison across individuals with different missingness patterns. Even with this restriction our factor score regression results may be biased, especially for analyses with the independent variable factor scores, depending on whether the estimation error in the factor scores as a function of the different missingness patterns is correlated with the other terms in the regression models.

Structured missingness thus has the ability to impact both the estimation of the factor model and individual-level latent factor scores, and extensive consideration must be given to how to reduce its impact if it is necessary to the research question at hand.

### Tables S2, S5, and S6

#### Table S2. Items included in final factor model by UKB category.

| Primary | Secondary | Tertiary | Quaternary | N |
|---|---|---|---|---|
| Online follow-up | Work environment | Employment history | | 55 |
| UK Biobank Assessment Centre | Touchscreen | Psychosocial factors | Mental health | 33 |
| UK Biobank Assessment Centre | Verbal interview | Medications | | 29 |
| UK Biobank Assessment Centre | Touchscreen | Lifestyle and environment | Diet | 29 |
| Health-related outcomes | Hospital in-patient | Diagnoses | Summary Information (diagnoses) | 27 |
| UK Biobank Assessment Centre | Touchscreen | Lifestyle and environment | Physical activity | 20 |
| Online follow-up | Mental health | Traumatic events | | 18 |
| UK Biobank Assessment Centre | Touchscreen | Health and medical history | Medication | 15 |
| UK Biobank Assessment Centre | Verbal interview | Medical conditions | | 12 |
| UK Biobank Assessment Centre | Physical measures | Eye measures | Autorefraction | 12 |
| UK Biobank Assessment Centre | Touchscreen | Sociodemographics | Household | 12 |
| Online follow-up | Mental health | Depression | | 11 |
| UK Biobank Assessment Centre | Touchscreen | Health and medical history | Medical conditions | 11 |
| Biological samples | Assay results | Blood assays | Blood count | 10 |
| UK Biobank Assessment Centre | Touchscreen | Family history | | 9 |
| Additional exposures | Local environment | Residential air pollution | | 9 |
| Online follow-up | Mental health | Anxiety | | 9 |
| UK Biobank Assessment Centre | Touchscreen | Health and medical history | Eyesight | 8 |
| UK Biobank Assessment Centre | Touchscreen | Psychosocial factors | Social support | 7 |
| UK Biobank Assessment Centre | Touchscreen | Health and medical history | Pain | 7 |
| UK Biobank Assessment Centre | Touchscreen | Health and medical history | Hearing | 7 |
| UK Biobank Assessment Centre | Touchscreen | Lifestyle and environment | Sleep | 7 |
| Online follow-up | Work environment | Medical information | | 7 |
| UK Biobank Assessment Centre | Physical measures | Hearing test | | 6 |
| UK Biobank Assessment Centre | Touchscreen | Sociodemographics | Education | 6 |
| UK Biobank Assessment Centre | Touchscreen | Lifestyle and environment | Electronic device use | 6 |
| Online follow-up | Mental health | Mental distress | | 6 |
| UK Biobank Assessment Centre | Physical measures | Eye measures | Intraocular pressure | 6 |
| UK Biobank Assessment Centre | Touchscreen | Lifestyle and environment | Sun exposure | 6 |
| UK Biobank Assessment Centre | Touchscreen | Lifestyle and environment | Alcohol | 6 |
| UK Biobank Assessment Centre | Physical measures | Bone-densitometry of heel | | 6 |
| Online follow-up | Mental health | Alcohol use | | 5 |
| UK Biobank Assessment Centre | Physical measures | Anthropometry | Body size measures | 5 |
| UK Biobank Assessment Centre | Recruitment | Reception | | 4 |
| UK Biobank Assessment Centre | Touchscreen | Early life factors | | 4 |

| Primary | Secondary | Tertiary | Quaternary | N |
|---|---|---|---|---|
| UK Biobank Assessment Centre | Touchscreen | Sociodemographics | Employment | 4 |
| UK Biobank Assessment Centre | Touchscreen | Health and medical history | General health | 4 |
| UK Biobank Assessment Centre | Physical measures | Blood pressure | | 3 |
| UK Biobank Assessment Centre | Touchscreen | Lifestyle and environment | Smoking | 3 |
| Online follow-up | Mental health | Self-harm behaviours | | 3 |
| UK Biobank Assessment Centre | Touchscreen | Lifestyle and environment | Sexual factors | 3 |
| Biological samples | Assay results | Urine assays | | 3 |
| UK Biobank Assessment Centre | Physical measures | Arterial stiffness | | 3 |
| UK Biobank Assessment Centre | Cognitive function | Prospective memory | | 3 |
| UK Biobank Assessment Centre | Physical measures | Spirometry | | 3 |
| Online follow-up | Mental health | Happiness and subjective well-being | | 3 |
| UK Biobank Assessment Centre | Touchscreen | Health and medical history | Mouth | 3 |
| UK Biobank Assessment Centre | Physical measures | Eye measures | Visual acuity | 2 |
| UK Biobank Assessment Centre | Cognitive function | Reaction time | | 2 |
| Online follow-up | Mental health | Addictions | | 2 |
| UK Biobank Assessment Centre | Verbal interview | Early life factors | | 2 |
| UK Biobank Assessment Centre | Cognitive function | Pairs matching | | 2 |
| UK Biobank Assessment Centre | Physical measures | Hand grip strength | | 2 |
| Online follow-up | Mental health | Unusual and psychotic experiences | | 2 |
| Health-related outcomes | Hospital in-patient | Admission and discharge | Summary Information (admission and discharge) | 2 |
| Online follow-up | Mental health | Mania | | 2 |
| UK Biobank Assessment Centre | Cognitive function | Fluid intelligence | | 2 |
| Additional exposures | Local environment | Home locations | | 2 |
| UK Biobank Assessment Centre | Touchscreen | Health and medical history | Breathing | 2 |
| Additional exposures | Local environment | Residential noise pollution | | 1 |
| Online follow-up | Mental health | Cannabis use | | 1 |
| Online follow-up | Diet by 24-hour recall | Diet questionnaire performance | | 1 |
| UK Biobank Assessment Centre | Touchscreen | Health and medical history | Chest pain | 1 |
| UK Biobank Assessment Centre | Physical measures | Eye measures | Eye surgery/complications | 1 |
| Population characteristics | Baseline characteristics | | | 1 |
| UK Biobank Assessment Centre | Physical measures | Anthropometry | Impedance measures | 1 |
| UK Biobank Assessment Centre | Touchscreen | Sociodemographics | Other sociodemographic factors | 1 |
| UK Biobank Assessment Centre | Touchscreen | Health and medical history | Claudication and peripheral artery disease | 1 |
| Health-related outcomes | Hospital in-patient | Psychiatric | Summary Information (psychiatric) | 1 |
| UK Biobank Assessment Centre | Touchscreen | Health and medical history | Cancer screening | 1 |
| UK Biobank Assessment Centre | Verbal interview | Operations | | 1 |
| Online follow-up | Work environment | | | 1 |

#### Table S5. Items included in core data group by UKB category.

| Primary | Secondary | Tertiary | Quaternary | N |
|---|---|---|---|---|
| Online follow-up | Work environment | Employment history | | 159 |
| Health-related outcomes | Hospital in-patient | Diagnoses | Summary Information (diagnoses) | 50 |
| UK Biobank Assessment Centre | Touchscreen | Lifestyle and environment | Diet | 49 |
| UK Biobank Assessment Centre | Touchscreen | Family history | | 44 |
| UK Biobank Assessment Centre | Touchscreen | Psychosocial factors | Mental health | 41 |
| UK Biobank Assessment Centre | Verbal interview | Medications | | 35 |
| UK Biobank Assessment Centre | Physical measures | Anthropometry | Impedance measures | 32 |
| Biological samples | Assay results | Blood assays | Blood count | 31 |
| UK Biobank Assessment Centre | Physical measures | Eye measures | Autorefraction | 28 |
| UK Biobank Assessment Centre | Verbal interview | Medical conditions | | 28 |
| UK Biobank Assessment Centre | Touchscreen | Health and medical history | Medication | 23 |
| UK Biobank Assessment Centre | Touchscreen | Sociodemographics | Household | 22 |
| UK Biobank Assessment Centre | Touchscreen | Lifestyle and environment | Physical activity | 20 |
| Online follow-up | Mental health | Traumatic events | | 19 |
| UK Biobank Assessment Centre | Touchscreen | Health and medical history | Medical conditions | 17 |
| Additional exposures | Local environment | Residential air pollution | | 17 |
| UK Biobank Assessment Centre | Touchscreen | Lifestyle and environment | Sun exposure | 13 |
| UK Biobank Assessment Centre | Touchscreen | Health and medical history | Eyesight | 11 |
| Online follow-up | Mental health | Depression | | 11 |
| UK Biobank Assessment Centre | Touchscreen | Health and medical history | Hearing | 11 |
| Online follow-up | Work environment | Medical information | | 10 |
| UK Biobank Assessment Centre | Physical measures | Bone-densitometry of heel | | 10 |
| UK Biobank Assessment Centre | Touchscreen | Lifestyle and environment | Smoking | 9 |
| Online follow-up | Mental health | Anxiety | | 9 |
| UK Biobank Assessment Centre | Physical measures | Hearing test | | 9 |
| UK Biobank Assessment Centre | Cognitive function | Fluid intelligence | | 8 |
| UK Biobank Assessment Centre | Touchscreen | Early life factors | | 8 |
| Online follow-up | Mental health | Alcohol use | | 8 |
| UK Biobank Assessment Centre | Touchscreen | Health and medical history | Pain | 8 |
| UK Biobank Assessment Centre | Physical measures | Eye measures | Intraocular pressure | 8 |
| UK Biobank Assessment Centre | Touchscreen | Psychosocial factors | Social support | 8 |
| UK Biobank Assessment Centre | Touchscreen | Lifestyle and environment | Alcohol | 8 |
| UK Biobank Assessment Centre | Touchscreen | Lifestyle and environment | Electronic device use | 8 |
| UK Biobank Assessment Centre | Touchscreen | Sociodemographics | Education | 7 |
| UK Biobank Assessment Centre | Touchscreen | Lifestyle and environment | Sleep | 7 |
| UK Biobank Assessment Centre | Touchscreen | Health and medical history | Mouth | 7 |
| Online follow-up | Mental health | Mental distress | | 7 |
| UK Biobank Assessment Centre | Physical measures | Anthropometry | Body size measures | 6 |
| UK Biobank Assessment Centre | Physical measures | Spirometry | | 6 |
| UK Biobank Assessment Centre | Touchscreen | Sociodemographics | Employment | 6 |
| UK Biobank Assessment Centre | Recruitment | Reception | | 6 |

| Primary | Secondary | Tertiary | Quaternary | N |
|---|---|---|---|---|
| UK Biobank Assessment Centre | Cognitive function | Prospective memory | | 6 |
| Additional exposures | Local environment | Residential noise pollution | | 5 |
| UK Biobank Assessment Centre | Physical measures | Arterial stiffness | | 4 |
| UK Biobank Assessment Centre | Touchscreen | Health and medical history | General health | 4 |
| UK Biobank Assessment Centre | Touchscreen | Sociodemographics | Other sociodemographic factors | 4 |
| Online follow-up | Mental health | Happiness and subjective well-being | | 3 |
| Online follow-up | Mental health | Self-harm behaviours | | 3 |
| Biological samples | Assay results | Urine assays | | 3 |
| UK Biobank Assessment Centre | Touchscreen | Lifestyle and environment | Sexual factors | 3 |
| UK Biobank Assessment Centre | Cognitive function | Pairs matching | | 3 |
| Health-related outcomes | Hospital in-patient | Admission and discharge | Summary Information (admission and discharge) | 3 |
| UK Biobank Assessment Centre | Cognitive function | Reaction time | | 3 |
| UK Biobank Assessment Centre | Physical measures | Blood pressure | | 3 |
| Online follow-up | Mental health | Unusual and psychotic experiences | | 2 |
| Additional exposures | Local environment | Home locations | | 2 |
| UK Biobank Assessment Centre | Touchscreen | Health and medical history | Breathing | 2 |
| Online follow-up | Mental health | Addictions | | 2 |
| UK Biobank Assessment Centre | Physical measures | Eye measures | Visual acuity | 2 |
| UK Biobank Assessment Centre | Verbal interview | Early life factors | | 2 |
| Online follow-up | Mental health | Mania | | 2 |
| UK Biobank Assessment Centre | Physical measures | Hand grip strength | | 2 |
| UK Biobank Assessment Centre | Verbal interview | Operations | | 1 |
| UK Biobank Assessment Centre | Physical measures | Eye measures | Eye surgery/complications | 1 |
| Population characteristics | Baseline characteristics | | | 1 |
| Online follow-up | Work environment | | | 1 |
| Online follow-up | Mental health | Cannabis use | | 1 |
| UK Biobank Assessment Centre | Touchscreen | Health and medical history | Cancer screening | 1 |
| Health-related outcomes | Hospital in-patient | Psychiatric | Summary Information (psychiatric) | 1 |
| Online follow-up | Diet by 24-hour recall | | | 1 |
| UK Biobank Assessment Centre | Touchscreen | Health and medical history | Chest pain | 1 |
| UK Biobank Assessment Centre | Touchscreen | Health and medical history | Claudication and peripheral artery disease | 1 |
| Online follow-up | Diet by 24-hour recall | Diet questionnaire performance | | 1 |

**Table S6. Reasons for item exclusion from EFA to final model.**

| Item | Item Name | Reason for Removal | Removal Step |
|---|---|---|---|
| X1249 | Past tobacco smoking | pairwise colinearity >0.99 | after initial EFA fit |
| X134 | Number of self-reported cancers | minimum cell size <25 | pre-fit |
| X1369 | Beef intake | minimum cell size <25 | pre-fit |
| X137 | Number of treatments/medications taken | likely multicolinearity | after initial EFA fit |
| X1379 | Lamb/mutton intake | minimum cell size <25 | pre-fit |
| X1389 | Pork intake | minimum cell size <25 | pre-fit |
| X1418_2 | Milk type used: Semi-skimmed | pairwise colinearity >0.99 | after initial EFA fit |
| X1428_3 | Spread type: Other type of spread/margarine | pairwise colinearity >0.99 | after initial EFA fit |
| X1448_3 | Bread type: Wholemeal or wholegrain | pairwise colinearity (though under initial threshold) | subsequent iteration |
| X1707_2 | Handedness (chirality/laterality): Left-handed | inestimable correlation due to pairwise colinearity | pre-fit |
| X1717 | Skin colour | minimum cell size <25 | pre-fit |
| X20002_1220 | Non-cancer illness code, self-reported: diabetes | pairwise colinearity >0.99 | after initial EFA fit |
| X20003_1140884600 | Treatment/medication code: metformin | likely multicolinearity | subsequent iteration |
| X20116_0 | Smoking status: Never | inestimable correlation due to pairwise colinearity | pre-fit |
| X20116_1 | Smoking status: Previous | pairwise colinearity >0.99 | after initial EFA fit |
| X20126_0 | Bipolar and major depression status: No Bipolar or Depression | likely multicolinearity | subsequent iteration |
| X20126_3 | Bipolar and major depression status: Probable Recurrent major depression (severe) | likely multicolinearity | after initial EFA fit |
| X20126_4 | Bipolar and major depression status: Probable Recurrent major depression (moderate) | likely multicolinearity | after initial EFA fit |
| X20126_5 | Bipolar and major depression status: Single Probable major depression episode | likely multicolinearity | subsequent iteration |
| X20405_1 | Ever had known person concerned about, or recommend reduction of, alcohol consumption: Yes, but not in the last year | redundant item | |
| X20411_1 | Ever been injured or injured someone else through drinking alcohol: Yes, but not in the last year | redundant item | |
| X20524 | Sexual interference by partner or ex-partner without consent as an adult | cell count <10 when split by sex | after initial EFA fit |
| X22506_114 | Tobacco smoking: Never smoked | problemmatic smoking cluster | after initial EFA fit |
| X22617_4217 | Job SOC coding: Typists | cell count <10 when split by sex | after initial EFA fit |
| X24007 | Particulate matter air pollution (pm2.5) absorbance; 2010 | inestimable pairwise polyserial correlation due to low cell counts | after initial EFA fit |

| Item | Item Name | Reason for Removal | Removal Step |
|---|---|---|---|
| X24009 | Traffic intensity on the nearest road | problemmatic traffic cluster | after initial EFA fit |
| X24012 | Inverse distance to the nearest major road | problemmatic traffic cluster | after initial EFA fit |
| X24014 | Close to major road | problemmatic traffic cluster | |
| X24015 | Sum of road length of major roads within 100m | problemmatic traffic cluster | after initial EFA fit |
| X41248_1000 | Destinations on discharge from hospital (recoded): Usual Place of residence | pairwise colinearity (though under initial threshold) | after initial EFA fit |
| X6138_100 | Qualifications: None of the above | "None of the above" | after initial EFA fit |
| X6139_100 | Gas or solid-fuel cooking/heating: None of the above | "None of the above" | after initial EFA fit |
| X6140_100 | Heating type(s) in home: None of the above | "None of the above" | after initial EFA fit |
| X6142_4 | Current employment status: Unable to work because of sickness or disability | likely multicolinearity | subsequent iteration |
| X6144_5 | Never eat eggs, dairy, wheat, sugar: I eat all of the above | inestimable correlation due to pairwise colinearity | pre-fit |
| X6145_100 | Illness, injury, bereavement, stress in last 2 years: None of the above | "None of the above" | after initial EFA fit |
| X6146_100 | Attendance/disability/mobility allowance: None of the above | "None of the above" | after initial EFA fit |
| X6146_2 | Attendance/disability/mobility allowance: Disability living allowance | likely multicolinearity | subsequent iteration |
| X6146_3 | Attendance/disability/mobility allowance: Blue badge | likely multicolinearity | subsequent iteration |
| X6148_100 | Eye problems/disorders: None of the above | "None of the above" | after initial EFA fit |
| X6148_2 | Eye problems/disorders: Glaucoma | pairwise colinearity >0.99 | after initial EFA fit |
| X6148_4 | Eye problems/disorders: Cataract | pairwise colinearity (though under initial threshold) | after initial EFA fit |
| X6149_100 | Mouth/teeth dental problems: None of the above | "None of the above" | after initial EFA fit |
| X6150_100 | Vascular/heart problems diagnosed by doctor: None of the above | inestimable correlation due to pairwise colinearity | pre-fit |
| X6152_100 | Blood clot, DVT, bronchitis, emphysema, asthma, rhinitis, eczema, allergy diagnosed by doctor: None of the above | "None of the above" | after initial EFA fit |
| X6154_1 | Medication for pain relief, constipation, heartburn: Aspirin | pairwise colinearity >0.99 | after initial EFA fit |
| X6154_100 | Medication for pain relief, constipation, heartburn: None of the above | "None of the above" | after initial EFA fit |
| X6154_2 | Medication for pain relief, constipation, heartburn: Ibuprofen (e.g. Nurofen) | pairwise colinearity >0.99 | after initial EFA fit |
| X6154_3 | Medication for pain relief, constipation, heartburn: Paracetamol | pairwise colinearity >0.99 | after initial EFA fit |

| Item | Item Name | Reason for Removal | Removal Step |
|---|---|---|---|
| X6154_4 | Medication for pain relief, constipation, heartburn: Ranitidine (e.g. Zantac) | pairwise colinearity >0.99 | after initial EFA fit |
| X6154_5 | Medication for pain relief, constipation, heartburn: Omeprazole (e.g. Zanprol) | pairwise colinearity >0.99 | after initial EFA fit |
| X6155_100 | Vitamin and mineral supplements: None of the above | "None of the above" | after initial EFA fit |
| X6159_100 | Pain type(s) experienced in last month: None of the above | "None of the above" | after initial EFA fit |
| X6160_100 | Leisure/social activities: None of the above | "None of the above" | after initial EFA fit |
| X6179_100 | Mineral and other dietary supplements: None of the above | "None of the above" | after initial EFA fit |
| X680_2 | Own or rent accommodation lived in: Own with a mortgage | pairwise colinearity >0.99 | after initial EFA fit |
| X680_3 | Own or rent accommodation lived in: Rent - from local authority, local council, housing association | likely multicolinearity | subsequent iteration |
| X680_4 | Own or rent accommodation lived in: Rent - from private landlord or letting agency | likely multicolinearity | after initial EFA fit |
| X709 | Number in household | likely multicollinearity | after initial EFA fit |

### Figures S1 to S15

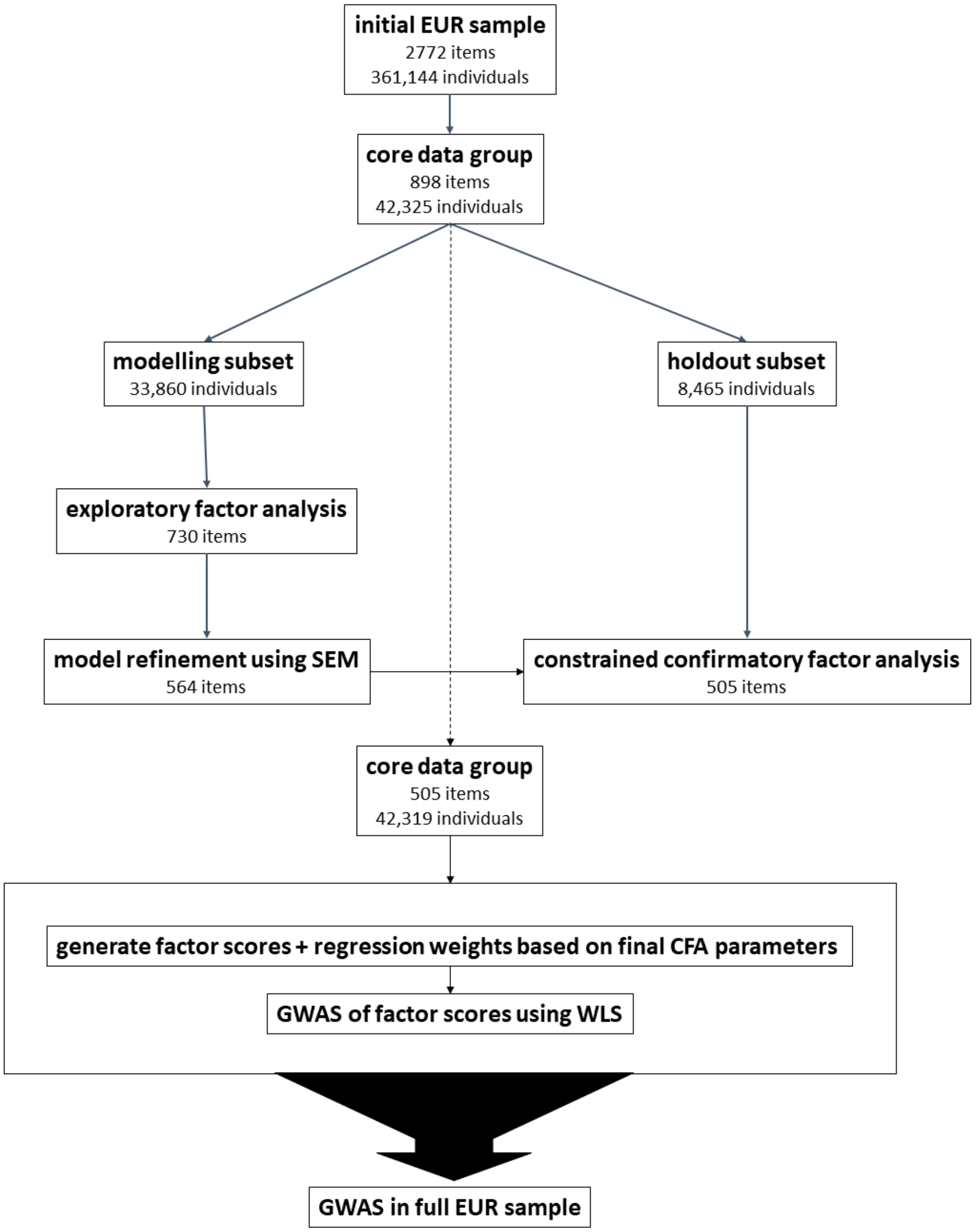


**Fig. S1. Schematic of overall analytic plan.** Displays the outline of analyses performed in the study as well as number of phenotypes and participants at each step.

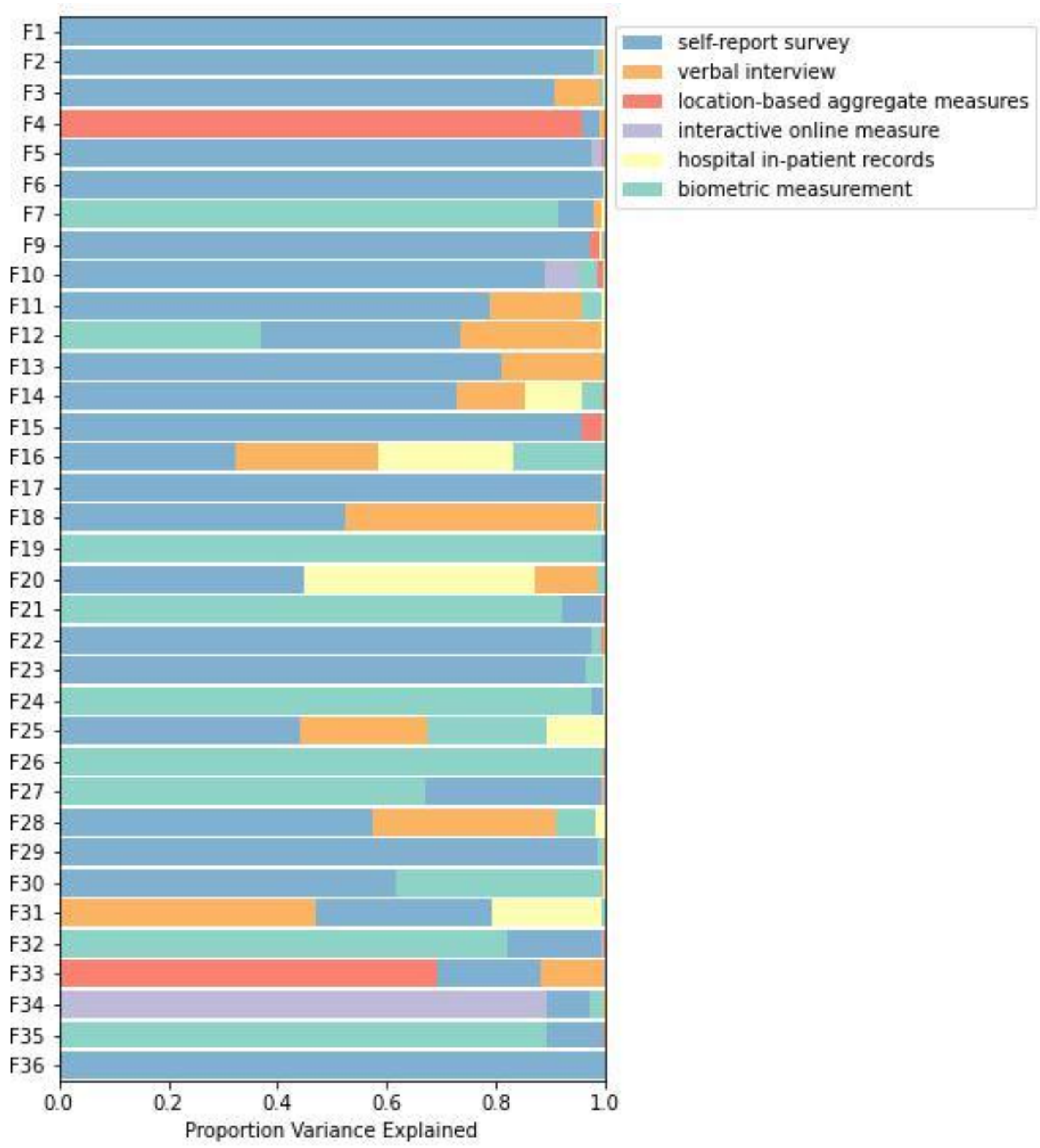


**Fig. S2. Representation of item types across factors.** Horizontal bars represent proportion variance explained in a given factor score by each of 6 major data types in UKB, estimated using hierarchical partitioning. To the left, factors are numbered in order of variance extraction in the exploratory factor analysis.

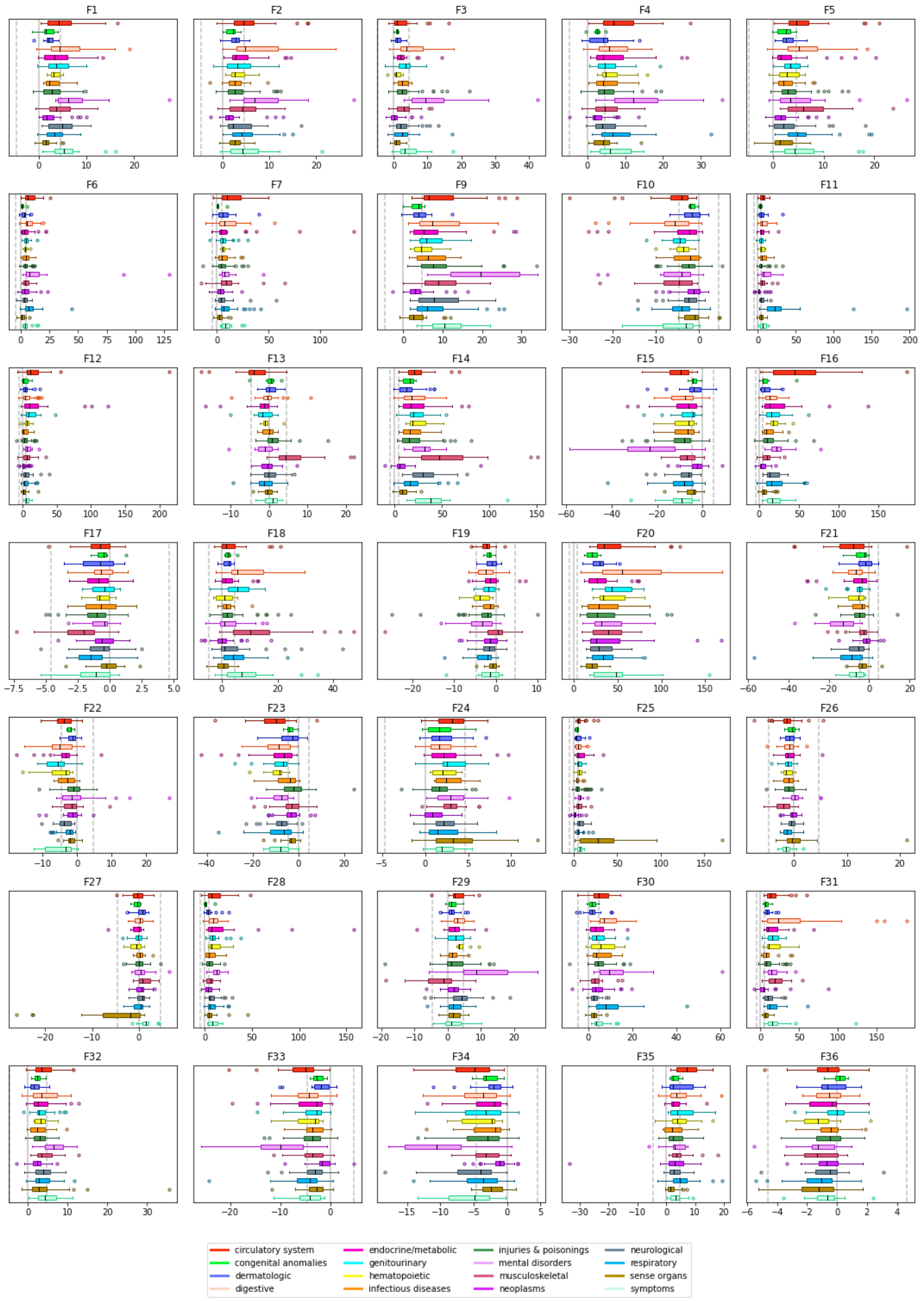


**Fig. S3. Phecode associations by factor.** Box-and-whisker plots are shown for associations with 403 derived medical phecodes grouped by category. Boxes represent the middle quartiles, with whiskers extending to 1.5x the interquartile range. Median values per category are indicated by individual black lines inside the boxes. The dotted grey lines represents the critical test statistics for significance at p<0.05 once correcting for comparisons across all 403 phecodes.

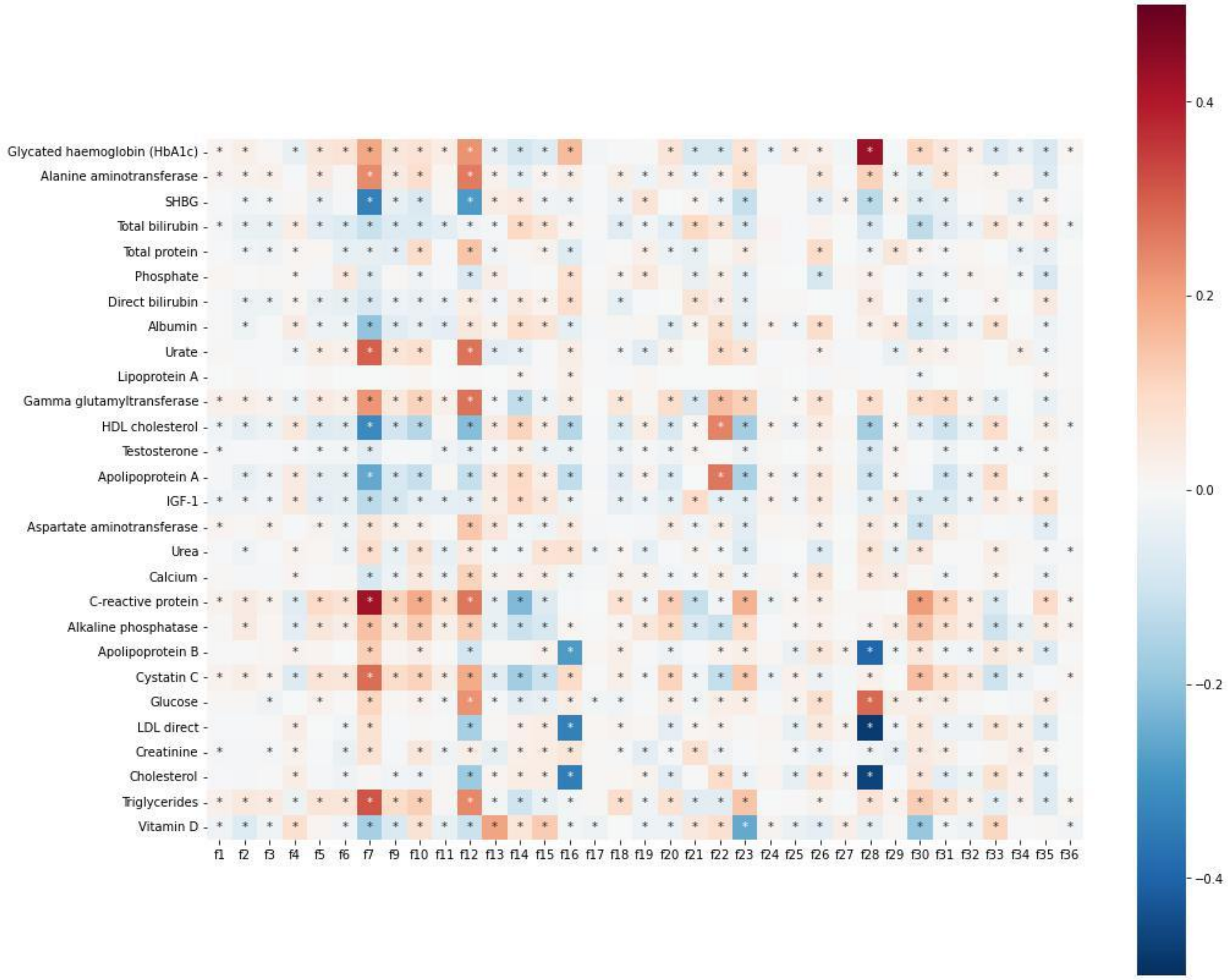


**Fig. S4. Biomarker associations by factor.** Phenotypic associations between factors and 28 biomarkers assayed in UKB. Colors represent the magnitude and direction of correlation, and asterisks (*) indicate which associations remain significant after correction for multiple testing (i.e., p<0.05 / (28 biomarkers x 35 factors)).

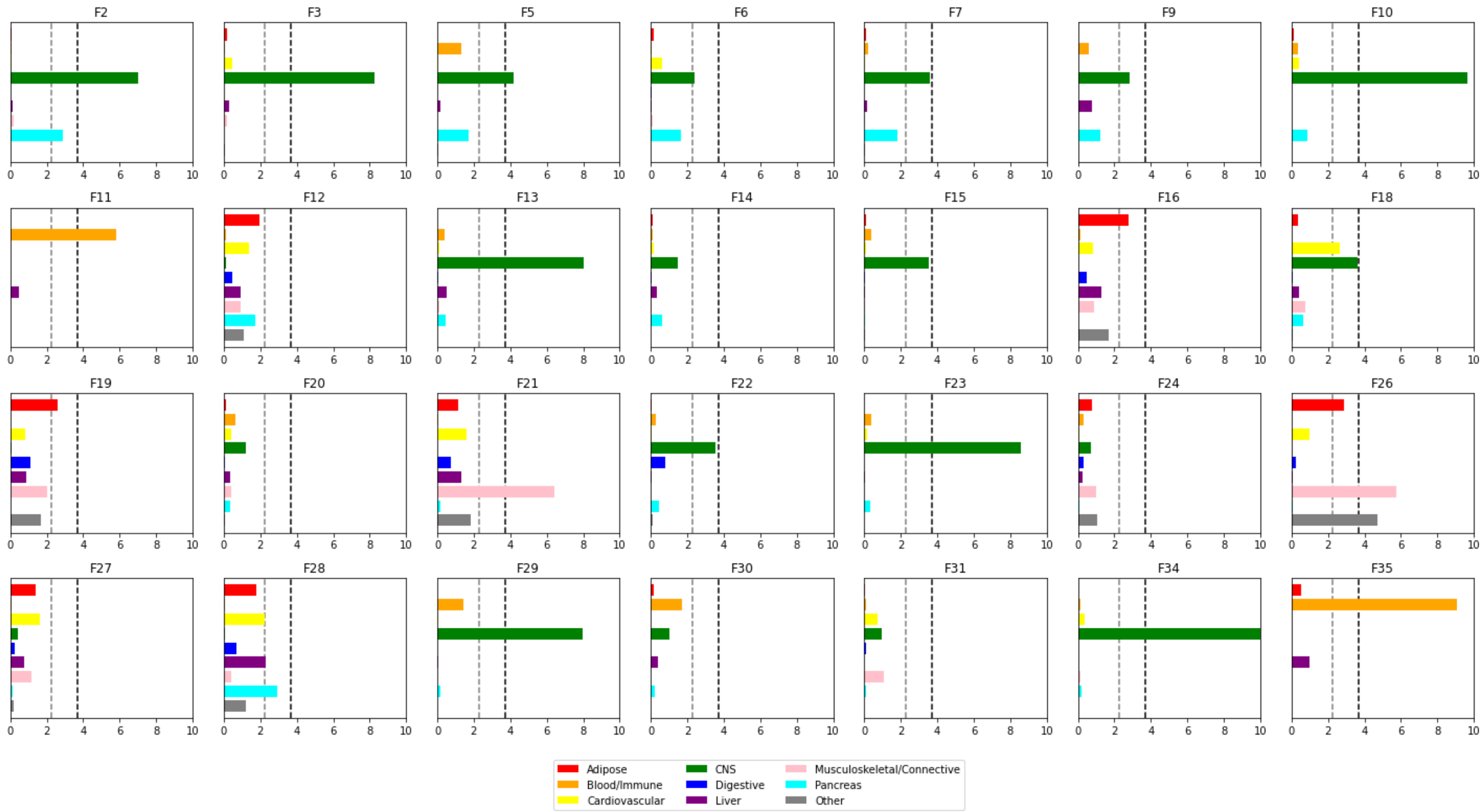


**Fig. S5. Heritability enrichment by cell type group.** Consistent with prior guidelines, only the 28 factors with $h^2$ z>7 were included in these analyses. -log10(p-value) is shown on the x-axis. The light grey dashed line represents the threshold for FDR-corrected significance at 0.05, while the black dashed line represents Bonferroni corrected threshold for 0.05 / (28 factors x 9 cell type groups).

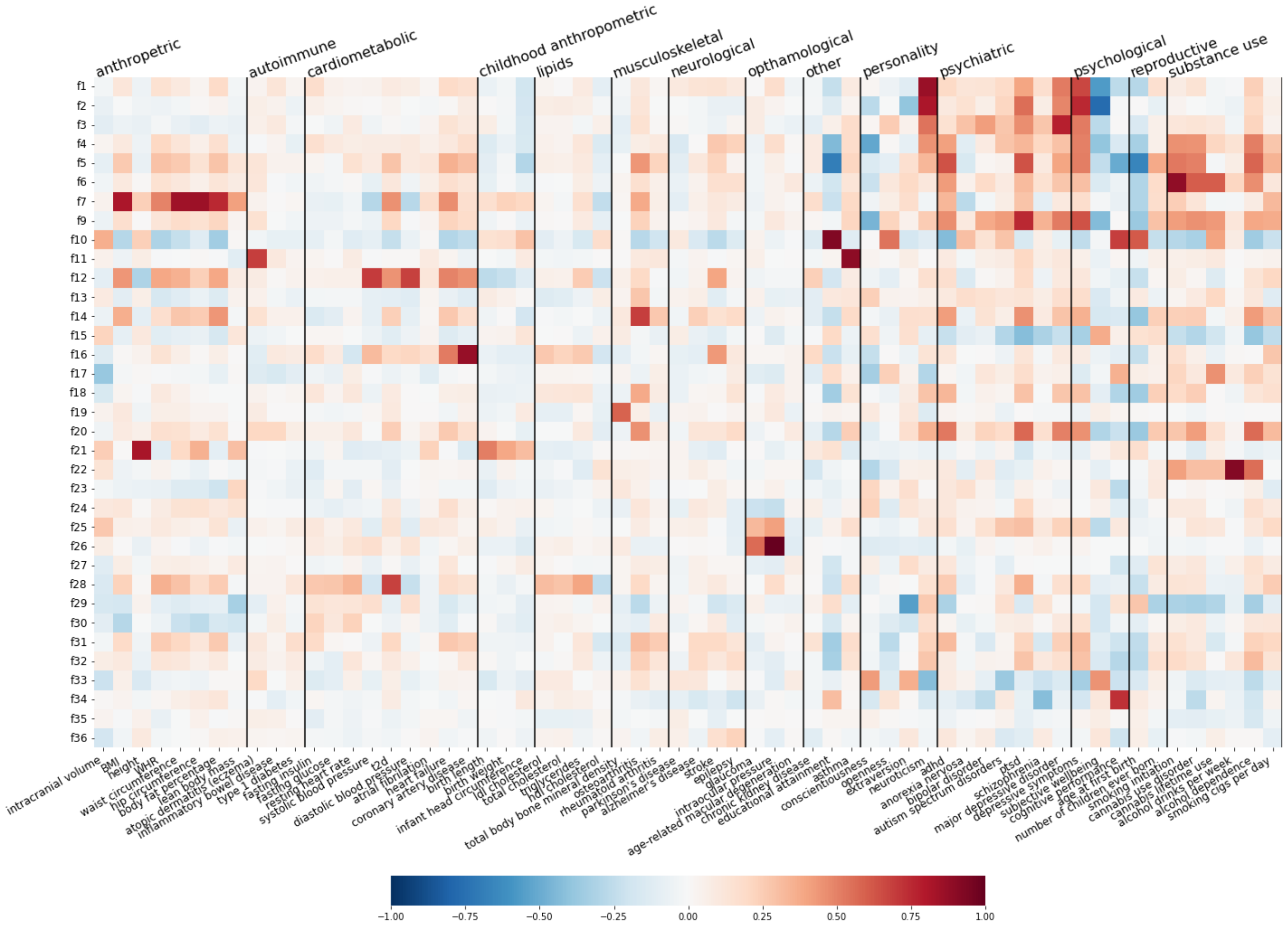


**Fig. S6. Genetic correlations of factors with outside traits.** The heatmap shows the estimated $r_g$ between our 35 factors and 62 selected outside summary statistics. Outside traits are grouped by general category. Color represents the magnitude and direction of genetic correlation.

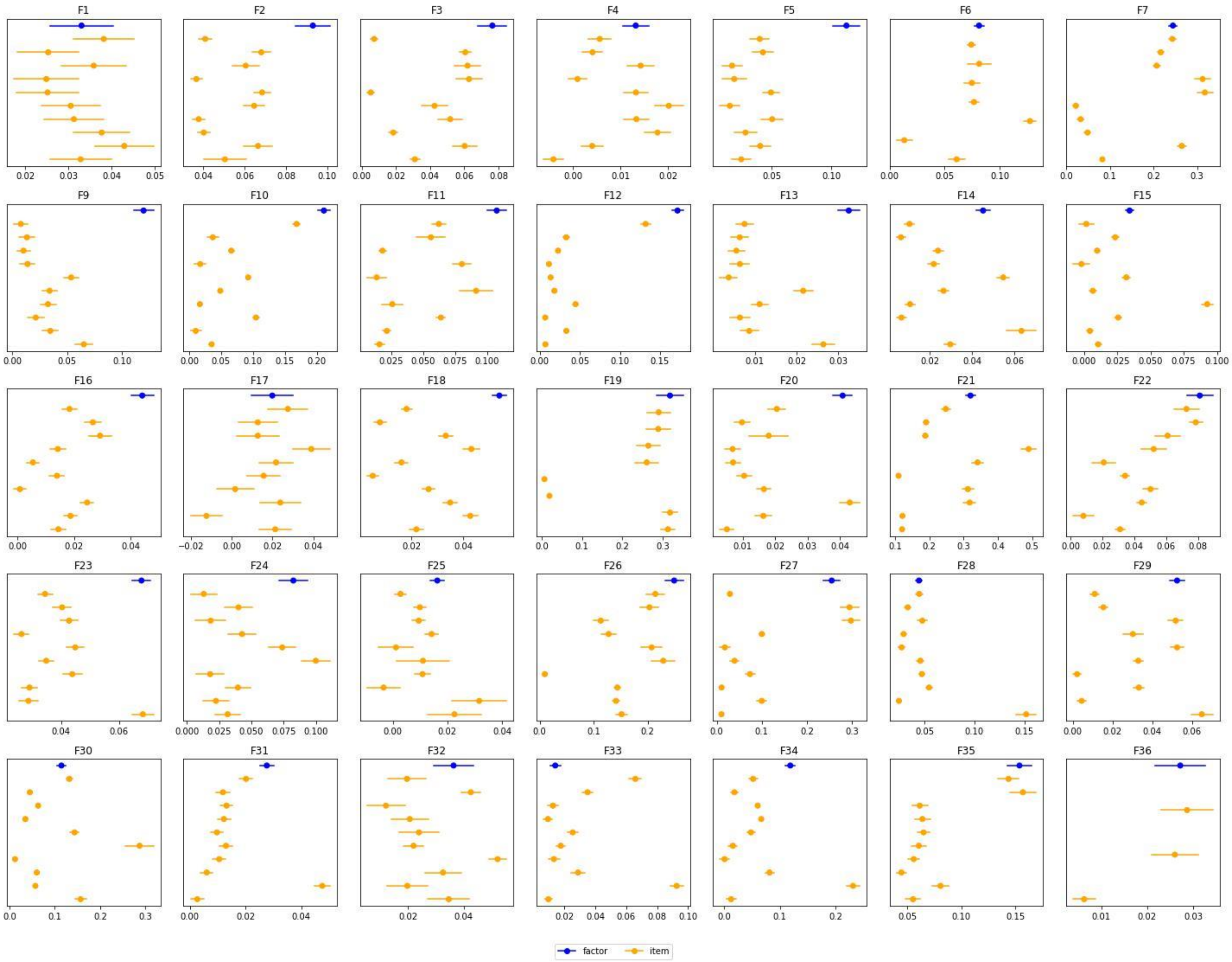


**Fig. S7. Heritability for factors versus top items.** Forest plots showing point estimates of heritability for factors (in blue) versus their top 10 items by loading (in orange). Error bars represent standard error.

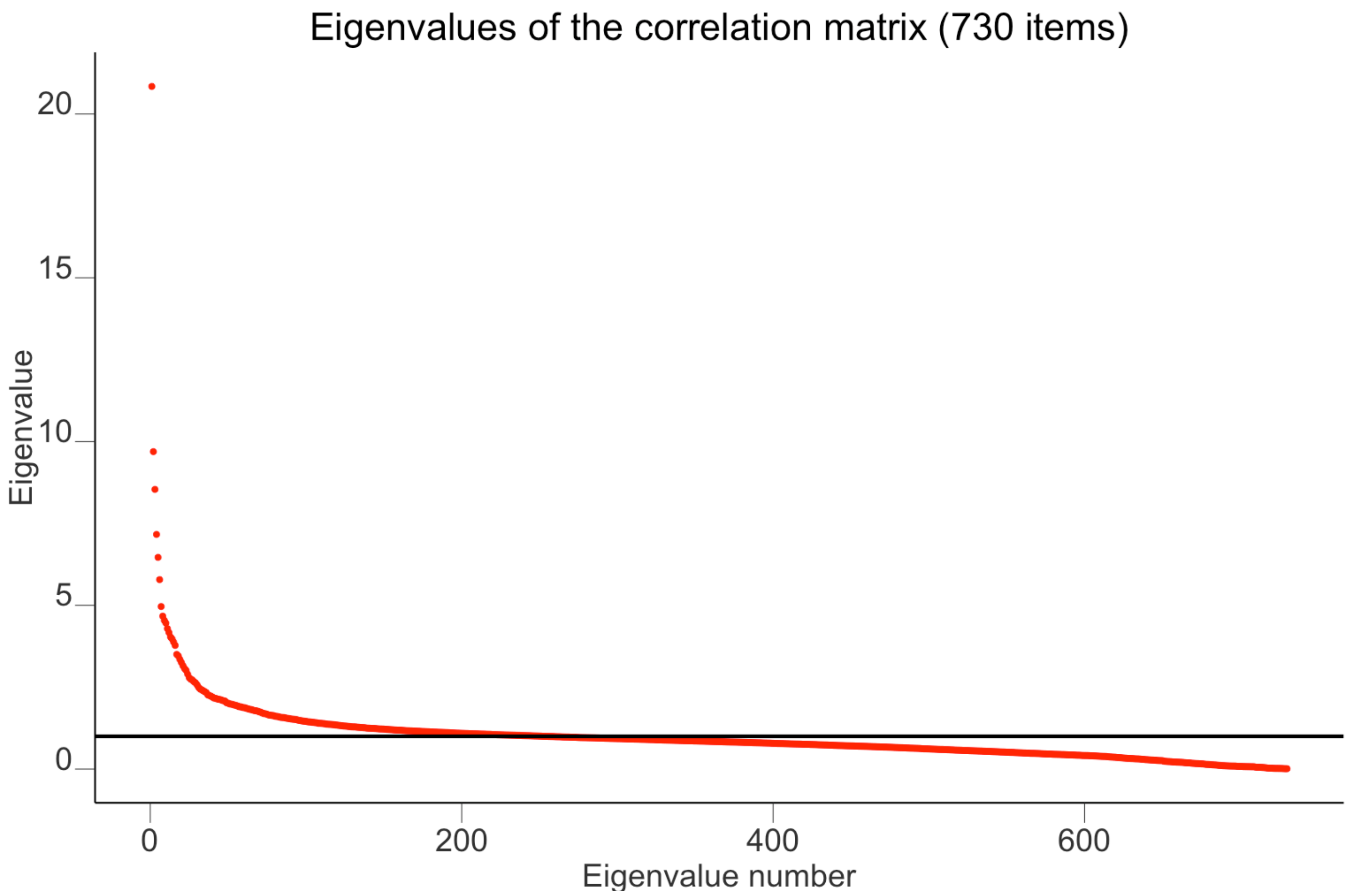


**Fig. S8. Scree plot of eigenvalues of the correlation matrix used for exploratory factor analysis.** The red dots show the 730 eigenvalues, and the horizontal dashed line corresponds to a value of 1.

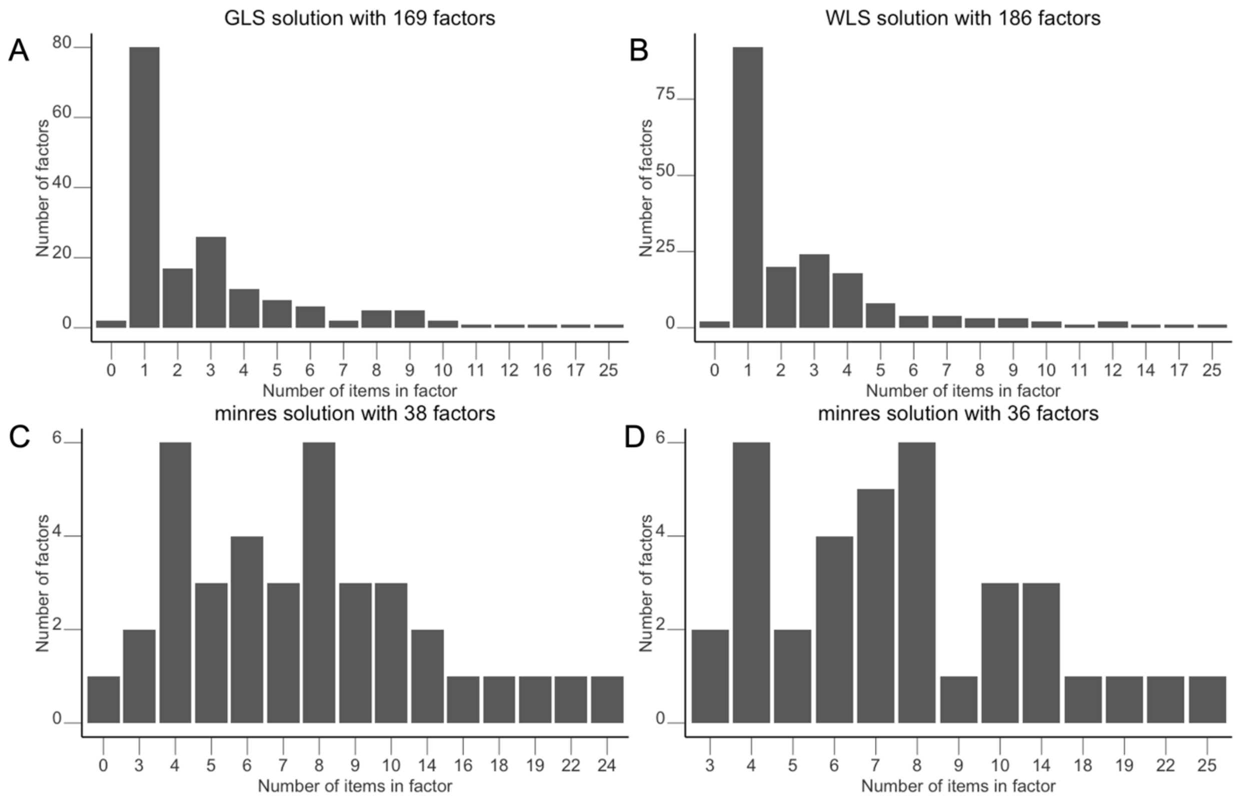


**Fig. S9. Distribution of number of items in each factor for different factor models.** **A-C:** GLS, WLS and MINRES methods with maximum number of factors (no ultra-Heywood cases). **D:** final EFA solution of 36 factors using the MINRES method.

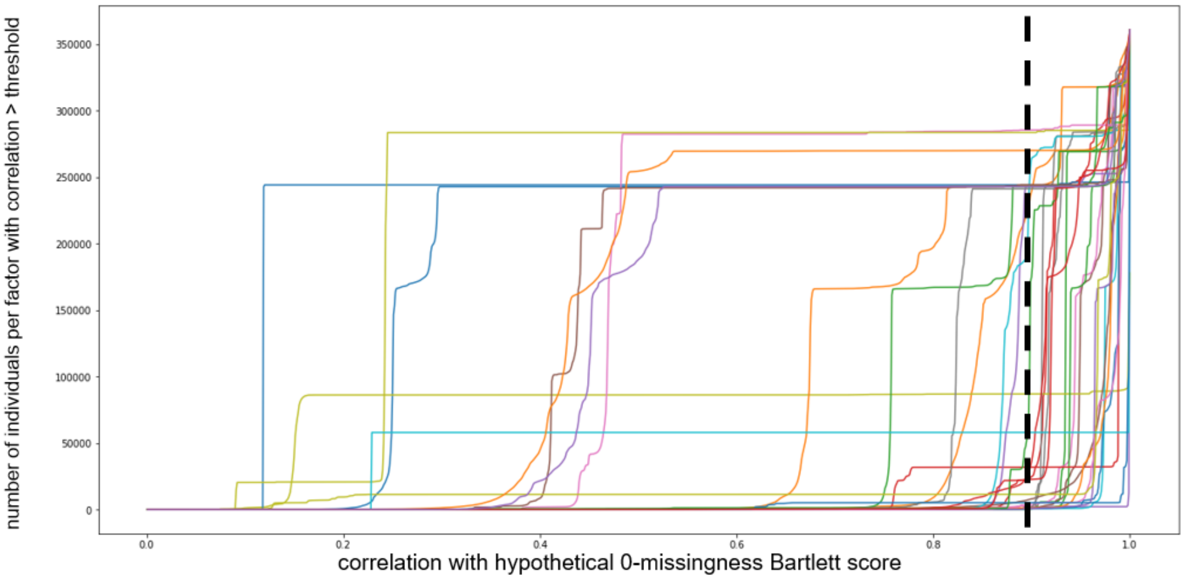


**Fig. S10. Factor score thresholding to account for structured missingness.** X-axis shows the correlation, for a given missingness pattern in the data, with a hypothetical 0-missingness Bartlett score. The y-axis shows the cumulative number of individuals per factor with a correlation value above that threshold. Major "jumps" in the data indicate the influence of structured missingness. The dashed black line represents our chosen threshold for inclusion, with an $r^2>0.80$.

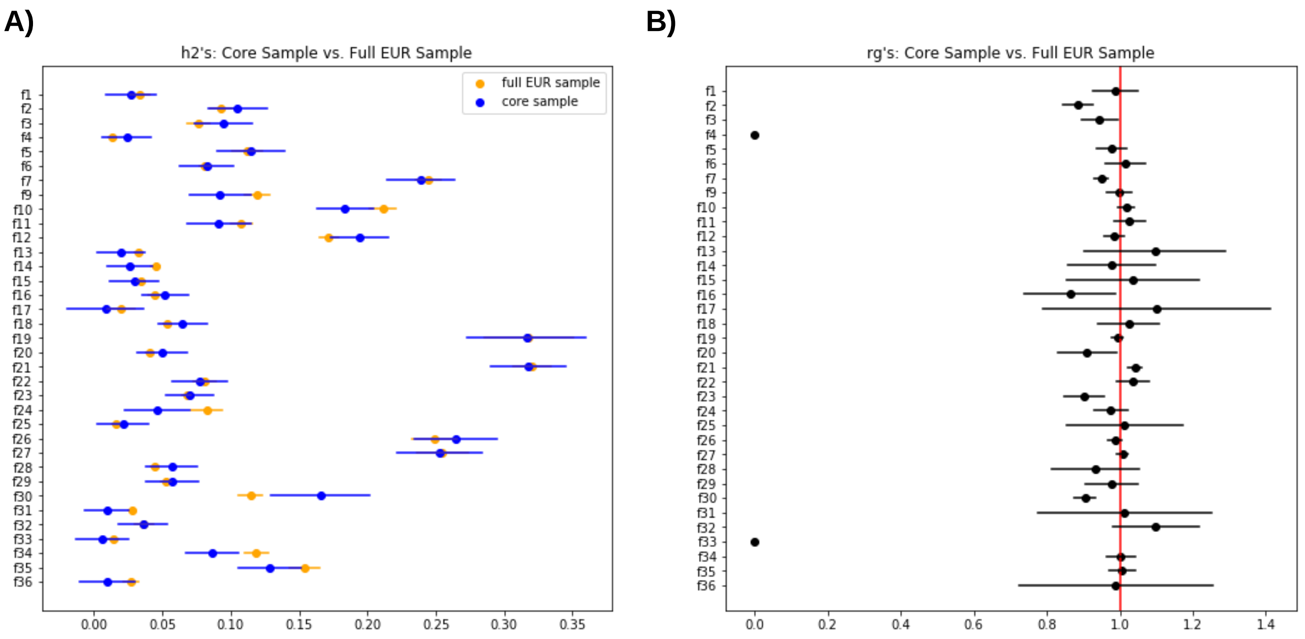


**Fig. S11. Differences in genetic architecture between core and full-European-ancestry samples.** Panel **A** shows the estimated heritability for each factor in the core sample (in blue) versus the full EUR-ancestry sample (in orange). Panel **B** shows the point estimate of the genetic correlation between both samples. Genetic correlations that could not be estimated (e.g., due to lack of heritability in one of the samples) are shown as $r_g$=0. Error bars for both panels represent standard errors.

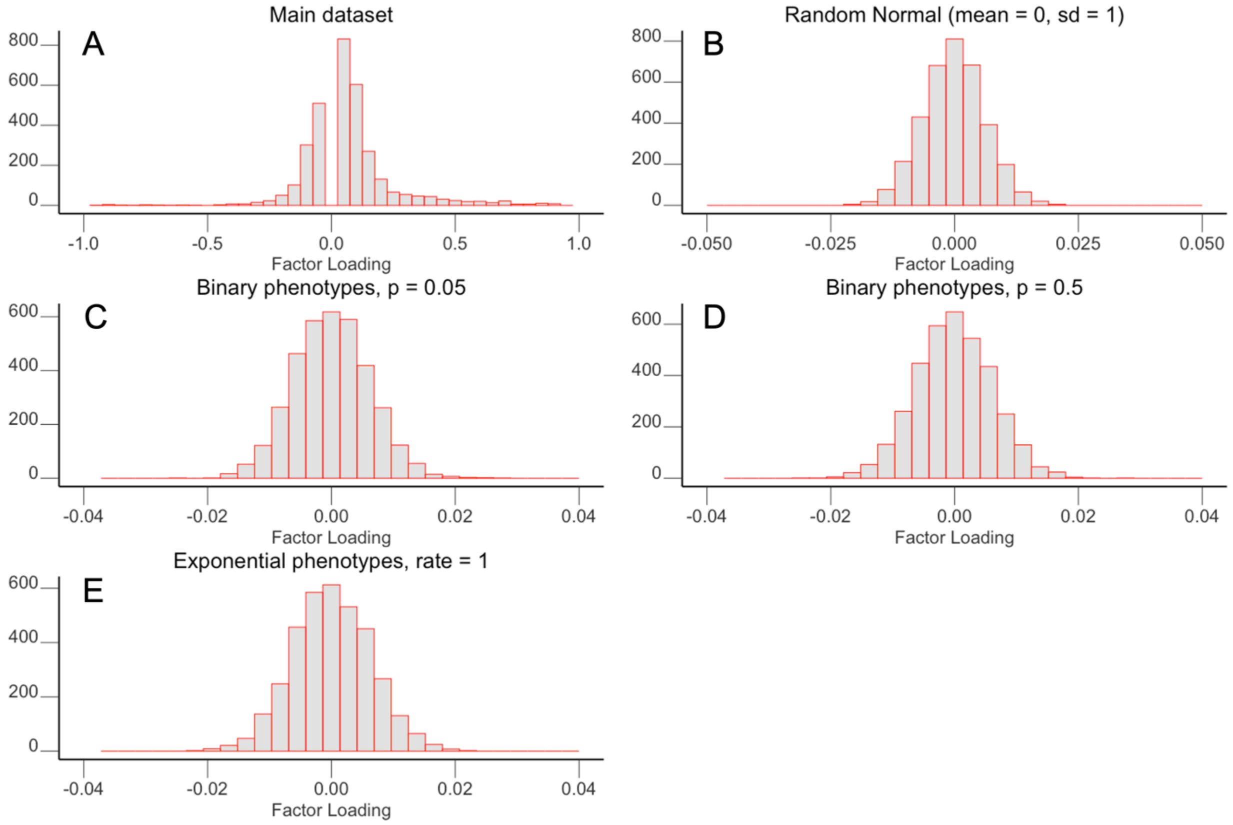


**Fig. S12. Comparison of factor loading distributions of the MINRES-36 model for randomly generated items and real data. A)** main dataset (730 items). Loadings between -0.05 and 0.05 were excluded to show the full range more clearly. **B)** Loadings for 100 random normally distributed traits. **C)** and **D)** Loadings for 100 binary traits (binomial distribution with p = 0.05 and p = 0.5, respectively). **E)** Loadings for 100 random exponential traits (rate = 1).

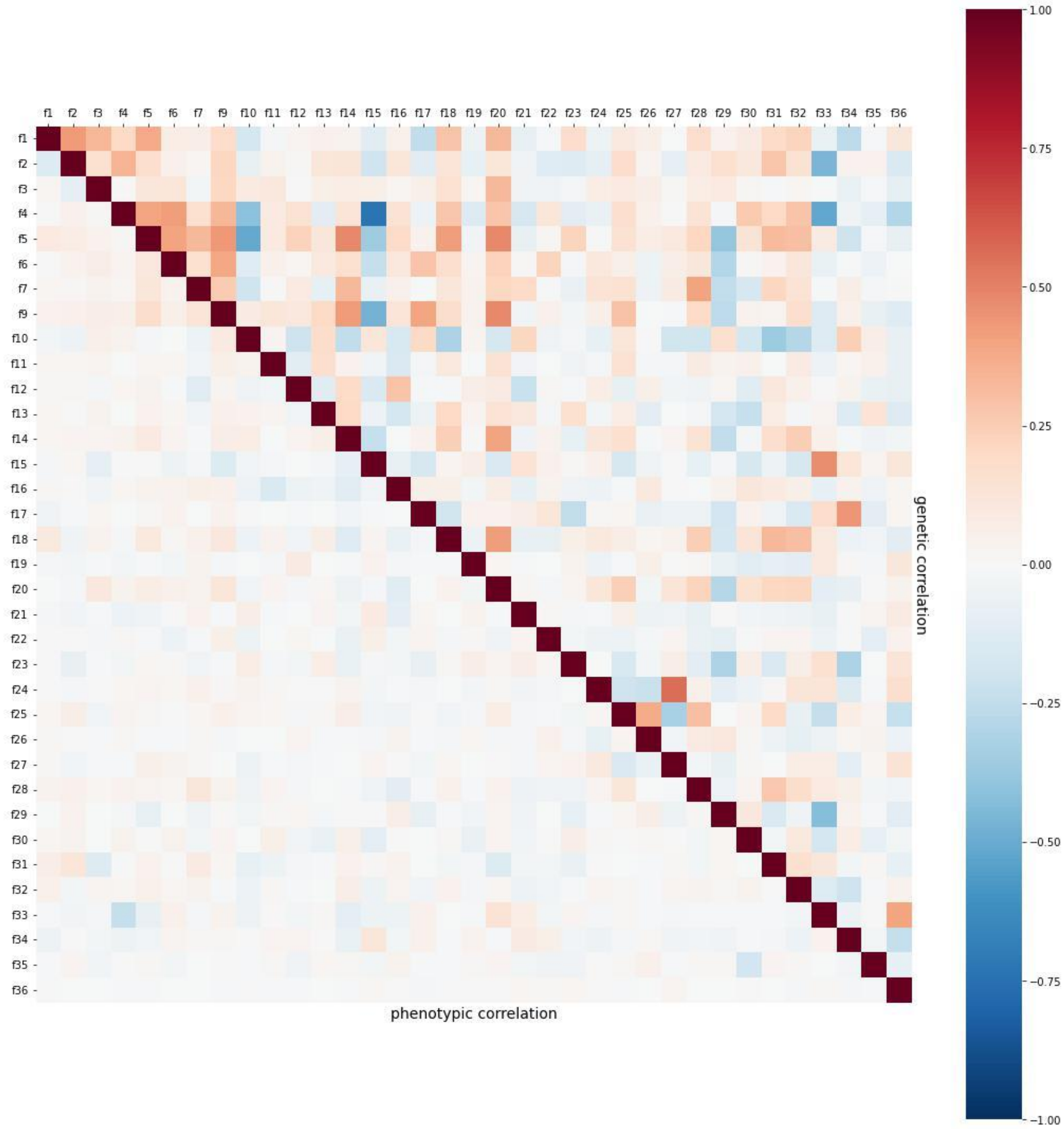


**Fig. S13. Phenotypic and genetic correlations across factors.** Phenotypic correlations between factors are shown in the lower triangle, and genetic correlations are shown in the upper triangle. Color indicates the magnitude and direction of correlation.

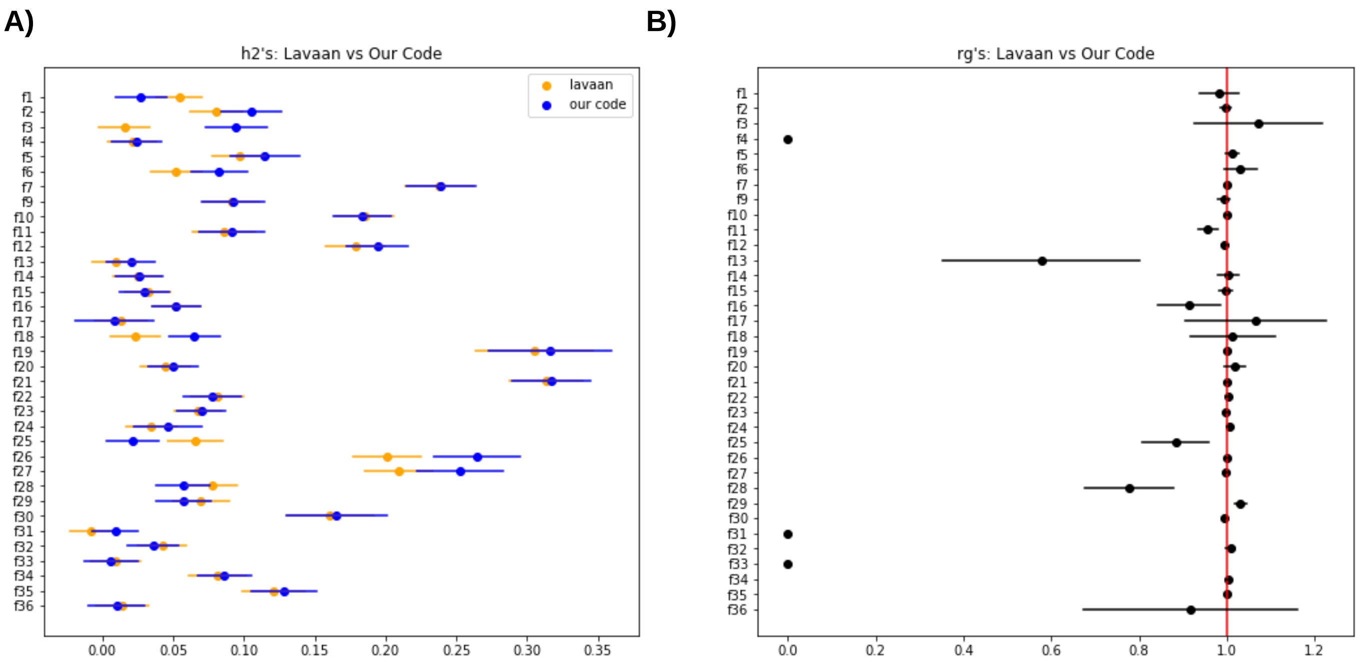


**Fig. S14. Differences in genetic architecture between factor scores estimated using lavPredict and our adapted Bartlett scoring methodology.** Panel **A** shows the estimated heritability for each factor when factor scores were generated using our adapted Bartlett scoring methodology (in blue) versus using the default estimator in lavaan (in orange), both in the core sample. Panel **B** shows the point estimate of the genetic correlation between both methods in the core sample. Genetic correlations that could not be estimated (e.g., due to lack of heritability for at least one of the methods) are shown as $r_g$=0. Error bars for both panels represent standard errors.

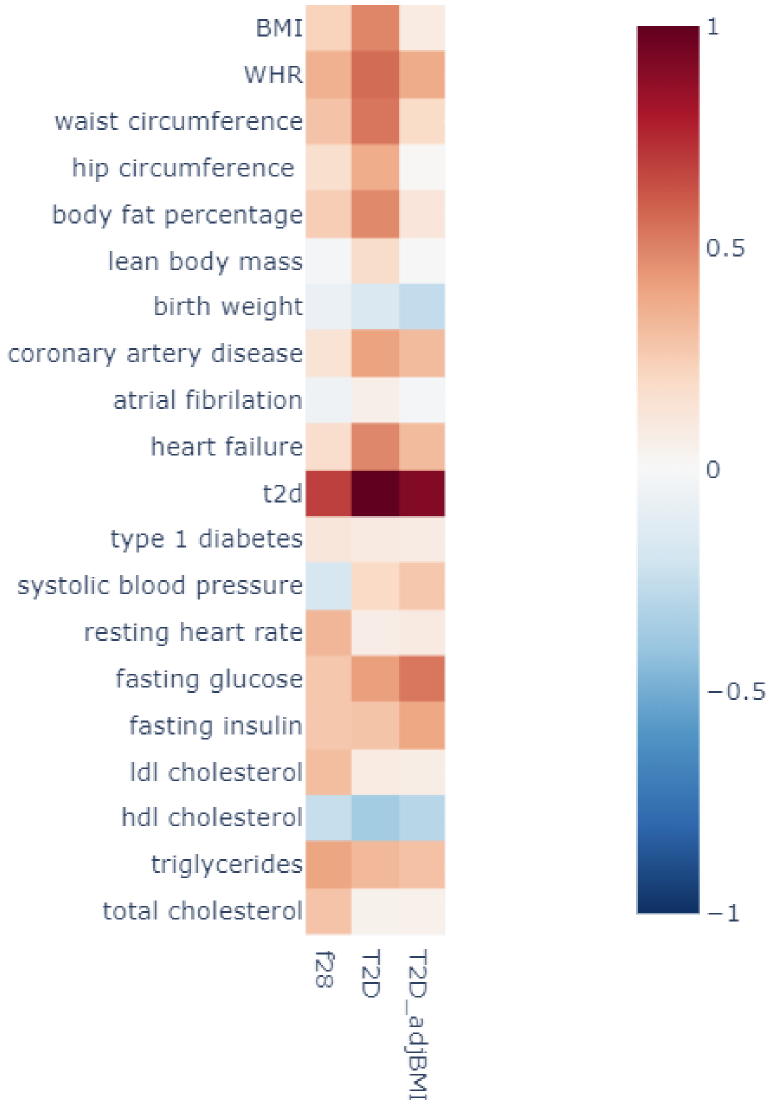


**Fig. S15. Demonstration of the impact of orthogonalization on genetic architecture of Factor 28 versus and outside GWAS of Type 2 Diabetes.** The heatmap shows the estimated $r_g$ between 20 selected outside cardiometabolic summary statistics and 1) our Factor 28, 2) an outside GWAS of Type 2 Diabetes, and 3) and outside GWAS of Type 2 Diabetes adjusted for BMI. Color represents the magnitude and direction of genetic correlation.
